# Saudi Arabian SARS-CoV-2 genomes implicate a mutant Nucleocapsid protein in modulating host interactions and increased viral load in COVID-19 patients

**DOI:** 10.1101/2021.05.06.21256706

**Authors:** Tobias Mourier, Muhammad Shuaib, Sharif Hala, Sara Mfarrej, Fadwa Alofi, Raeece Naeem, Afrah Alsomali, David Jorgensen, Amit Kumar Subudhi, Fathia Ben Rached, Qingtian Guan, Rahul P Salunke, Amanda Ooi, Luke Esau, Olga Douvropoulou, Raushan Nugmanova, Sadhasivam Perumal, Huoming Zhang, Issaac Rajan, Awad Al-Omari, Samer Salih, Abbas Shamsan, Abbas Al Mutair, Jumana Taha, Abdulaziz Alahmadi, Nashwa Khotani, Abdelrahman Alhamss, Ahmed Mahmoud, Khaled Alquthami, Abdullah Dageeg, Asim Khogeer, Anwar M. Hashem, Paula Moraga, Eric Volz, Naif Almontashiri, Arnab Pain

## Abstract

Monitoring SARS-CoV-2 spread and evolution through genome sequencing is essential in handling the COVID-19 pandemic. The availability of patient hospital records is crucial for linking the genomic sequence information to virus function during the course of infections. Here, we sequenced 892 SARS-CoV-2 genomes collected from patients in Saudi Arabia from March to August 2020. From the assembled sequences, we estimate the SARS-CoV-2 effective population size and infection rate and outline the epidemiological dynamics of import and transmission events during this period in Saudi Arabia. We show that two consecutive mutations (R203K/G204R) in the SARS-CoV-2 nucleocapsid (N) protein are associated with higher viral loads in COVID-19 patients. Our comparative biochemical analysis reveals that the mutant N protein displays enhanced viral RNA binding and differential interaction with key host proteins. We found hyper-phosphorylation of the adjacent serine site (S206) in the mutant N protein by mass-spectrometry analysis. Furthermore, analysis of the host cell transcriptome suggests that the mutant N protein results in dysregulated interferon response genes. We provide crucial information in linking the R203K/G204R mutations in the N protein as a major modulator of host-virus interactions and increased viral load and underline the potential of the nucleocapsid protein as a drug target during infection.

## Main

The emergence of novel severe acute respiratory syndrome coronavirus 2 (SARS-CoV-2), which causes the respiratory coronavirus infectious disease 2019 (COVID-19), resulted in a pandemic that has triggered an unparalleled public health emergency ^1,2^. The global spread of SARS-CoV-2 depended fundamentally on human mobility patterns. This is highly pertinent to a country like the Kingdom of Saudi Arabia, which as of 22nd February 2021 had a total of 374,691 cases and 6,457 deaths ^3^. The kingdom frequently experiences major population movements, particularly religious mass gatherings. For instance, during Umrah and Hajj roughly 9.5 million pilgrims visit two Islamic sites in Makkah and Madinah annually ^4,5^ and the Ministry of Health takes extraordinary public health measures to keep the pilgrims safe and major outbreaks have been by and large avoided in recent years. Further, an estimated 5 million Shiite Saudi nationals travel to Iran for pilgrimage, which became an early source of COVID-19 infections in the region ^5,6^. This movement has been reflected in the early phase of COVID-19 transmission within Saudi, as the first case was officially reported in Qatif (Eastern Region) on March 2^nd^, 2020 ^7^.

Genomic epidemiology of emerging viruses has proven to be a useful tool for outbreak investigation and tracking the pathogen’s progress ^8,9^. Currently, over half a million complete and high coverage genomes are accessible on GISAID^10,11^, which aids immensely in tracking the viral sequences globally ^12^. Novel SARS-CoV-2 variants are continuously arising and besides providing signals for epidemiological tracking, a subset of the resulting variants will have a functional impact on transmission and infection ^13-15^. It is therefore critical to monitor the genetic viral diversity throughout the pandemic.

In this study, we sequenced 892 SARS-CoV-2 genomes from nasopharyngeal swab samples of patients from the four main cities, Jeddah, Makkah, Madinah, and Riyadh, as well as a small number of patients from the Eastern region of Saudi Arabia (Figure 1, Table S1, Table S2). We analyzed the genomes to investigate the nucleotide changes and multiple mutation events that represent the first 6 months of the locally circulating pandemic lineages of the SARS-CoV-2 in Saudi Arabia and searched for potential association of polymorphic sites in the genome with available hospital records including severe disease and case fatality rates among the COVID-19 patients. We performed phylogenetic analysis to visualize the genetic diversity of SARS-CoV-2 and the nature of transmission lineages during March-August, 2020. We have presented a snapshot of the genomic variation landscapes of the SARS-CoV-2 lineages in our study population and linked specific set of mutation events in the N gene to viral loads in a diverse population of COVID-19 patients in Saudi Arabia (Figure S1). Finally, we experimentally show the functional impact of these mutations in the N protein on the virus’ interactions with the host.

**Figure 1.**
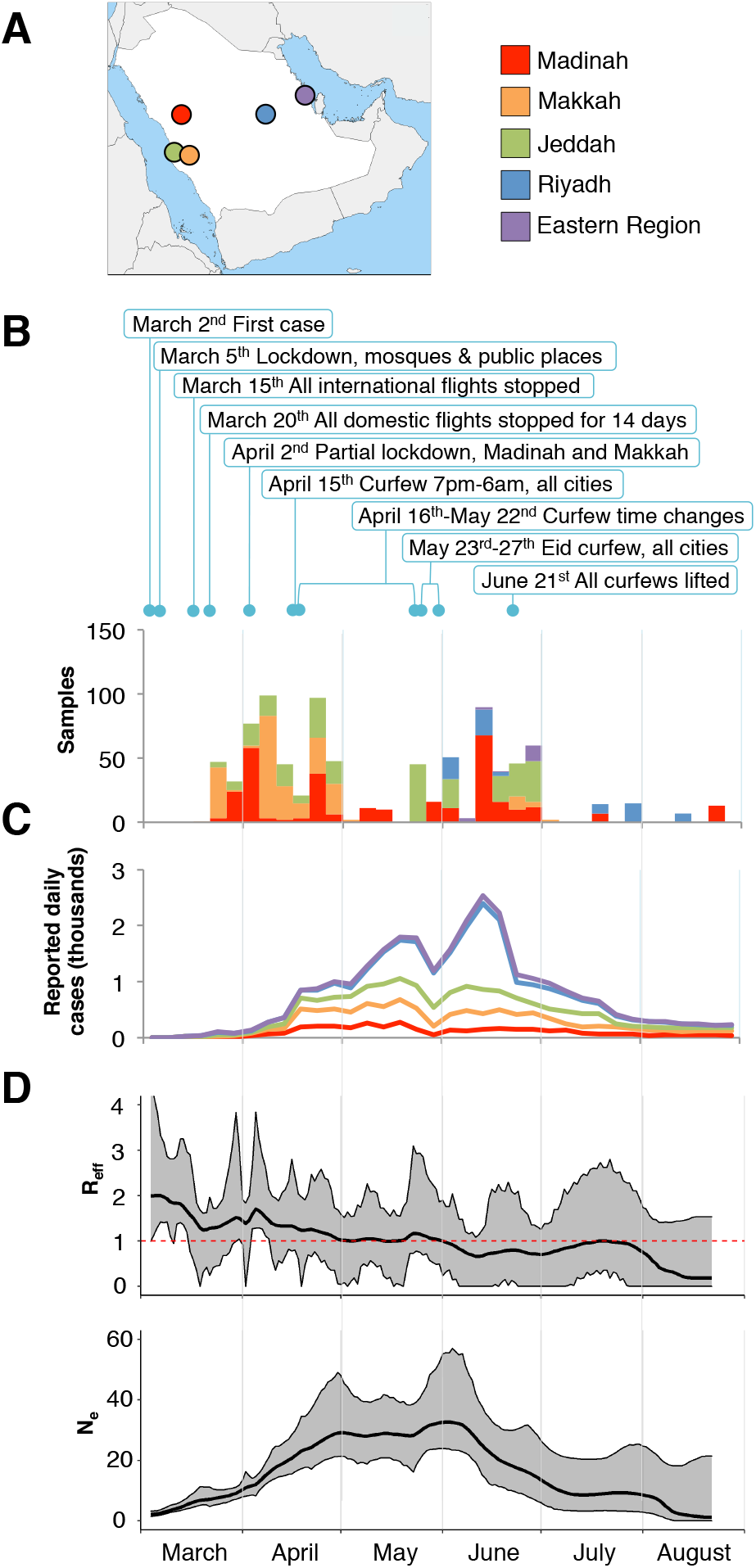
Sample overview and population genetics. A) Locations of the sampling cities within Saudi Arabia. B) Combined numbers of samples retrieved from the 4 cities and the Eastern region during the first six months of the pandemic. Cities are colored as in panel A. Months are shown at the bottom of the figure, and each month is divided into 5-days intervals. New daily cases for the city of Khobar is shown on the Eastern Region plot. Major restrictions imposed by the Ministry of Health and by Royal decrees are indicated above plots. C) Combined average numbers of new daily cases (Supplementary Information). D) Estimate of effective reproduction number [R_eff_] over time in Saudi Arabia (top) and the estimate of effective population size [Ne], the relative population size required to produce the diversity seen in the sample (bottom). The red horizontal red line represents an R of 1, the level required to sustain epidemic growth. Grey confidence areas denote the 95% credible intervals.

## Results

### SNP calling and phylodynamics of SARS-CoV-2 samples from Saudi Arabia

We sequenced and assembled SARS-CoV-2 genomes from 892 patient samples. This group includes 144 patients that were placed in quarantine and had either mild symptoms or were asymptomatic. The remaining patients were all hospitalized (Table S2). Data on comorbidities were available for 689 patients, and included diabetes, hypertension, Lung BA, Kidney CDK, Cardiovascular, and Cerebrovascular diseases. Patient outcome data was available for 850 samples, and 199 patients (23%) died during hospitalization.

From the 892 assembled viral genomes collected over a period of 6 months, we found a total of 836 single-nucleotide polymorphisms (SNPs) compared to the Wuhan SARS-CoV-2 reference (GenBank accession: NC_045512) (Figure S2). The observed numbers of SNPs relative to the Wuhan reference follow the numbers observed in global samples (Figure S3). We further detected 41 indels of which 26 reside in coding regions (Table S3). Most indels were specific to a single sample, and no identical indel was found in more than four samples. Compared with global SNP data, seven SNPs were found in higher frequencies (absolute difference > 0.1) in samples from Saudi Arabia (Figure S2). These include the Spike protein D614G (A23403G) and three consecutive SNPs causing the R203K and G204R changes in the nucleoprotein (G28881A, G28882A, and G28883C). Together with all sequences from Saudi Arabia available on GISAID on December 31^st^ 2020, the assembled sequences were used to construct the effective population size and growth rate estimates of SARS-CoV2 over the course of the first wave of the epidemic. The skygrowth model ^16^ (Figure 1D) shows a downward trend in the effective reproduction number (R) over time with the timely introduction and maintenance of effective non-pharmaceutical interventions by the Saudi Ministry of Health. Following the lifting of restrictions towards the end of June, the model estimates that R remained below or at 1 to the end of the period covered by the genetic data presented in this study. The effective population size (N_e_) represents the relative diversity of the sequences collected in Saudi Arabia over the course of the outbreak (Figure 1D). The model predicts a peak in viral diversity at the beginning of June. This is ahead of the peak number of cases reported nationally and is likely influenced by the earlier peak in reported cases in the three cities, which contribute the most viral sequences to this analysis (Madinah, Makkah and Jeddah).

A maximum-likelihood phylogenetic analysis revealed that samples from Saudi Arabia represent 5 major Nextstrain clades^12^, 19A-B and 20A-C (Figure 2A). This highlighted the clade 20A that all carried the Nucleocapsid (N) protein R203K/G204R mutations ^17^ with high incidences of ICU hospitalizations. These samples were predominantly coming from Jeddah. Through time-scaled phylogenies dates of importation events were then estimated for each clade. The majority of importations for all clades were inferred to have occurred early in the outbreak, primarily in March and early April (Figure 2B). Inferring importation events from a phylogenetic tree with estimated dating of nodes we see an early import from Asia followed by multiple imports from different continents (Figure 2C).

**Figure 2.**
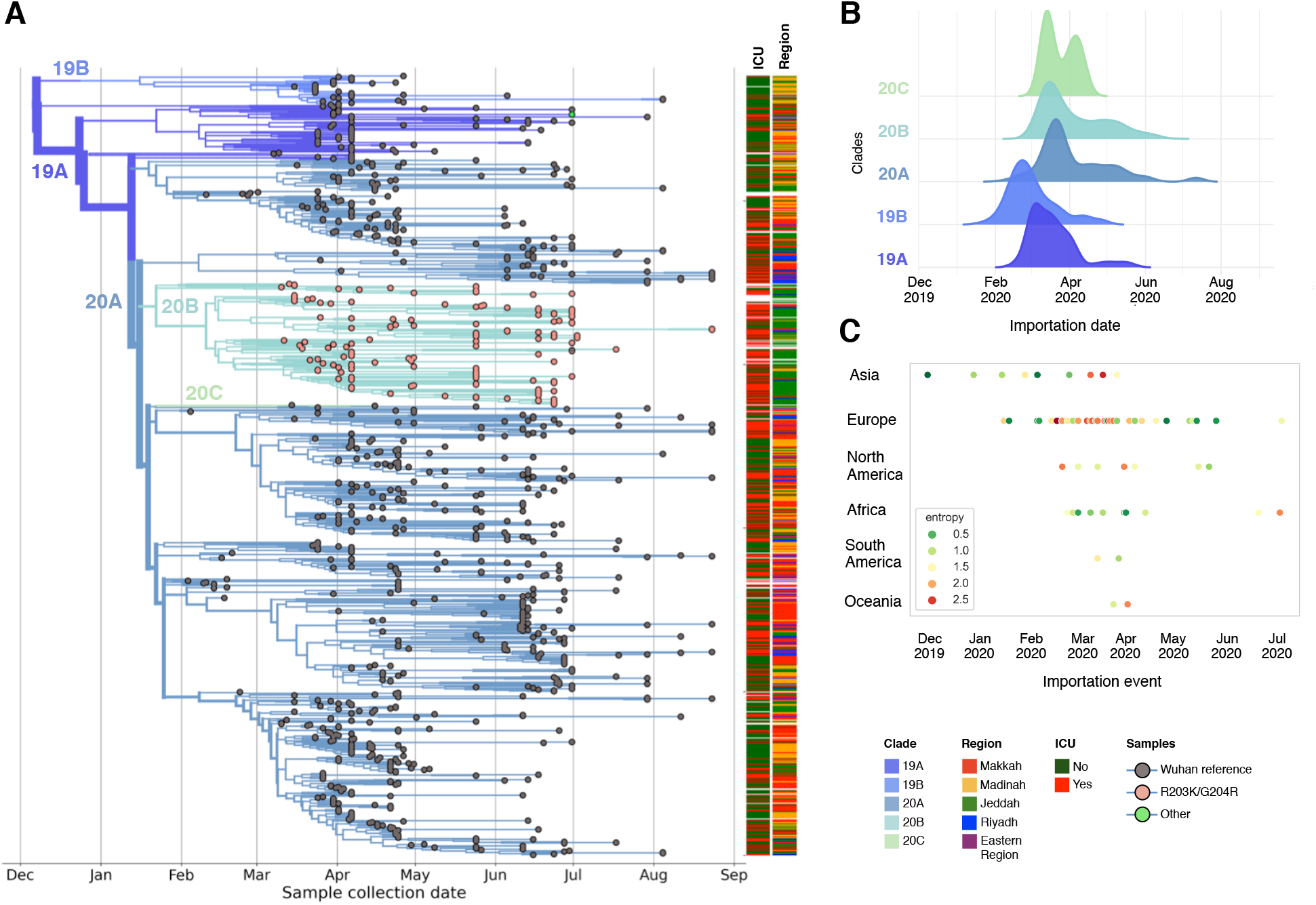
Phylodynamics of SARS-CoV-2 samples in Saudi Arabia. A) Global Time scaled phylogeny of 952 Saudi samples coloured by Nextstrain clades. Samples are shown as circles and coloured according to their genotype at genome positions 28,881-28,883. ICU status and sampling region are indicated on the right of the tree. B) Distributions of importation dates for the 5 Nextstrain (nextstrain.org) clades found in Saudi Arabia coloured by clade. C) Importation events estimated by traversing a phylogenetic tree to identify branches that resulted in transitions into Saudi Arabia from another country. Events are coloured by their normalised Shannon entropy, which measures the uncertainty inherent in the country of origin for a given importation event.

### Origin of R203K/G204R SNPs and importation into Saudi Arabia

A dated phylogeny of global samples showed that samples with the R203K/G204R SNPs are predominantly found in Nextstrain clades 20A, 20B and 20C, and do not form a monophyletic group (Figure S4). Furthermore, a few samples are further found in the early appearing 19A and 19B clades. However, due to the limited number of mutations separating SARS-CoV-2 genomes constructing a reliable and robust phylogeny is problematic^18^, and while different clades may be well supported, the exact relationship between clades is often less easily resolved. Although phylogenetic trees of SARS-CoV-2 genomes may appear to robustly reflect transmission events, collapsing branches with low support will typically result in extensive polytomies^19,20^. Additionally, the placement of individual virus genomes may be hampered by systematic errors, homoplasies, potential recombination, or co-infection of multiple virus strains^18,19,21-23^. It is therefore not clear if the phylogenetic distribution of samples with R203K/G204R SNPs reflects multiple independent origins of the SNPs, although it is evident that the R203K/G204R SNPs appeared early in the pandemic spread (Figure S4). Consistent with this, we find the earliest estimated importation events of R203K/G204R SNPs in late January 2020, most likely from Italy (Figure S5). This thus suggest a slightly earlier importation date than the estimate of importation events of clade 20B (Figure 2B). Within our sampling window we observe an apparent transient increase in the frequency of R203K/G204R SNPs (Figure 3A) in accordance with earlier observations ^17,24^. This peak is similarly observed in global data up until the fall of 2020, where the R203K/G204R SNPs once again increase along with the Spike protein Y501N mutation in the B1.1.17 lineage ^25^ (Figure 3A).

**Figure 3.**
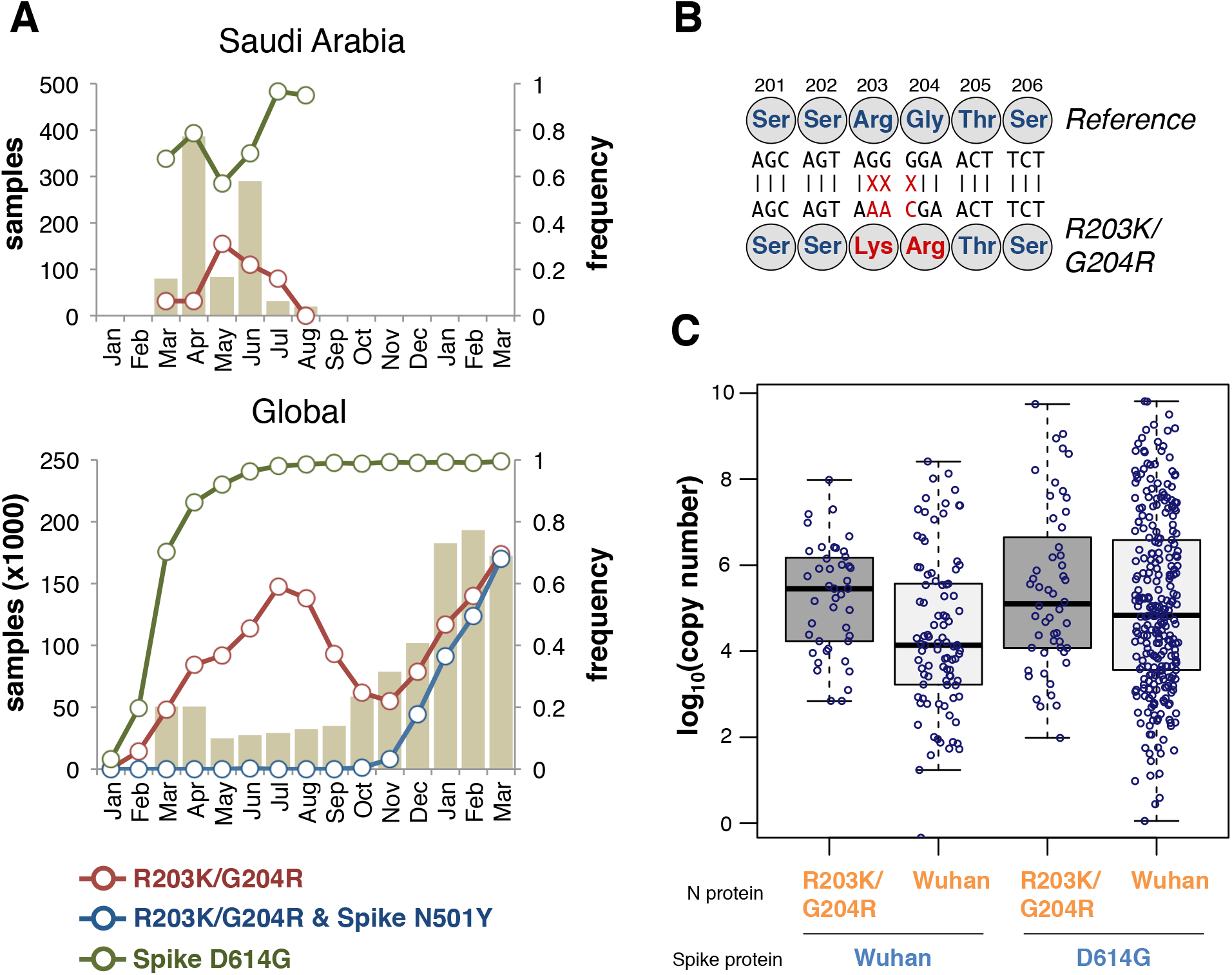
Higher viral loads in samples with R203K/G204R SNPs. A) Top: The numbers of samples from Saudi Arabia presented in this study are shown as bars by their sampling date (January 2020-March 2021). Bottom: Samples deposited in GISAID. On both plots, lines show the fraction of samples having the R203K/G204R SNPs (red line), having both the R203K/G204R SNPs and the Spike protein N501Y SNP (blue line), and having the Spike protein D614G SNP (green line). B) Overview of the three SNPs underlying the N protein R203K/G204R changes. Amino acid numbers in the N protein is shown above. C) Boxplot showing the distribution of virus copy number derived from Ct measurements. Ct values from the N1 primer pairs were normalized by RNase P primer pair values and converted to copy numbers from a standard curve. Only samples processed using the TaqPath™ kit (Thermofisher) were included (see Supplementary Information). Copy numbers are shown for four different haplotypes (as indicated below the plot) corresponding to virus genome positions 28,881-28,883 (orange text) and 23,403 (blue text). ‘Wuhan’ denotes the genotypes in the reference genome (NC_045512).

### A mutant form of the Nucleocapsid (N) protein associated with higher viral loads in COVID-19 patients in Saudi Arabia

A genome-wide association study between SARS-CoV-2 SNPs and patient mortality identified the three consecutive SNPs (G28881A, G28882A, G28883C) underlying the R203K/G204R mutations (Figure 3B, Figure S6). Of the 892 assembled genomes, 882 (98.9%) genomes either have the three reference alleles, GGG, or the three mutant alleles, AAC, at positions 28,881-28,883. This is similarly found in global samples deposited in GIASID in 2020, where 99.7% of samples with SNPs at positions 28,881-28,883 contain all three SNPs (Figure S7). In our samples, no other SNPs co-occur with the R203K/G204R SNPs (Figure S8). The frequency of the R203K/G204R SNPs is markedly higher in samples from Jeddah, where the observed frequency of 0.38 is more than 10-fold higher than the average of the other cities (Table S2). Within-host polymorphism has been observed for the R203K/G204R SNPs either resulting from co-infection of multiple strains or cross-sample contamination^21^. Co-infection of SARS-CoV-2 is demonstrated through observations of recombination between genetically distinct lineages^26^. To rule out cross-sample contamination, we investigated the levels of within-host polymorphisms in a range of SNP positions and found this more consistent with cases of co-infection among patients rather than contamination issues (Supplementary Information, Figure S9).

Using multivariable regression, we next evaluated the effect of the R203K/G204R SNPs on mortality, severity, and viral load in our COVID-19 patients samples for which limited amount of clinical meta-datasets were available. Disease severity was defined as deceased patients and patients admitted to ICU. For mortality and severity, we first fitted a linear model using R203K/G204R SNPs as a covariate. Then we fitted adjusted models by including gender, age, comorbidities, hospital and time. Additionally, the Spike protein D614G SNP that is associated with higher viral load ^13^ was included. Age and time were included using smoothing splines to allow for potential non-linear relationships ^27^. Using an unadjusted logistic regression, we observed a positive and statistically significant association between R203K/G204R SNPs and severity. Specifically, we found that the log-odds of severity increased by 1.16, 95% CI 0.70-1.64. In the adjusted model, the log-odds decreased to 0.66, 95% CI 0.01-1.32. That is, we found a borderline significant association between R203K/G204R SNPs and severity. The relationship between mortality and R203K/G204R SNPs was positive and statistically significant in the unadjusted model with log-odds equal to 1.39, 95% 0.99-1.79. We also found a positive and statistically significant relationship adjusting for D614G SNP, gender, age, comorbidities, and hospital (log-odds = 0.62, 95% 0.13-1.10). However, after adjusting for time as a variable, there was no longer any association between R203K/G204R SNPs and mortality (log-odds: 0.25, 95% CI -0.30-0.80). The models thus suggest a temporal component in our observations, and it is important to note that the recorded mortalities from Jeddah are concentrated on just a few dates (Figure S10). Unfortunately, our data set does not allow us to assess if the observed mortality rates are the result of shifts in treatment regimes or admission procedures during the sampling window.

We then tested if R203K/G204R SNPs were associated with higher viral copy numbers as indicated by the cycle threshold (Ct) values obtained through quantitative PCRs (see Methods). From the unadjusted regression we found a positive and statistically significant relationship between R203K/G204R SNPs and log_10_(viral copy number), with the mean of log_10_(viral copy number) values increasing by 1.03 units (95% CI 0.67-1.46). The significance was still observed in the adjusted model, although the relationship decreased to 0.57 units (95% CI 0.13-1.01) (Figure 3C). Similarly, the adjusted model showed a significant relationship between D614G SNPs and log_10_(viral copy number), with the mean values increasing by 0.32 units (95% CI 0.13-1.01), consistent with earlier reports ^13,28^. The positive and statistically significant association of R203K/G204R SNPs with higher viral load in critical COVID-19 patients suggests its functional implications during viral infection.

### N mutant protein has high oligomerization potential and RNA binding affinity

The SARS-CoV-2 N protein binds the viral RNA genome and is central to viral replication ^30^. Protein structure predictions have shown that the R203K/G204R mutations result in significant changes in protein structure ^24^, theoretically destabilizing the N structure ^31^, and potentially enhancing the protein’s ability to bind RNA and alter its response to serine phosphorylation events ^32^. The R203K/G204R mutations in the SARS-CoV-2 N protein are within the linkage region (LKR) containing the serine/arginine-rich motif (SR-rich motif) (Figure 4A), known to be involved in the oligomerization of N proteins ^33,34^. Protein cross-linking shows that N mutant protein (with the R203K/G204R mutations) has higher oligomerization potential compared to the control N protein (without the changed amino acids) at low protein concentration (Figure S11A-B).

**Figure 4.**
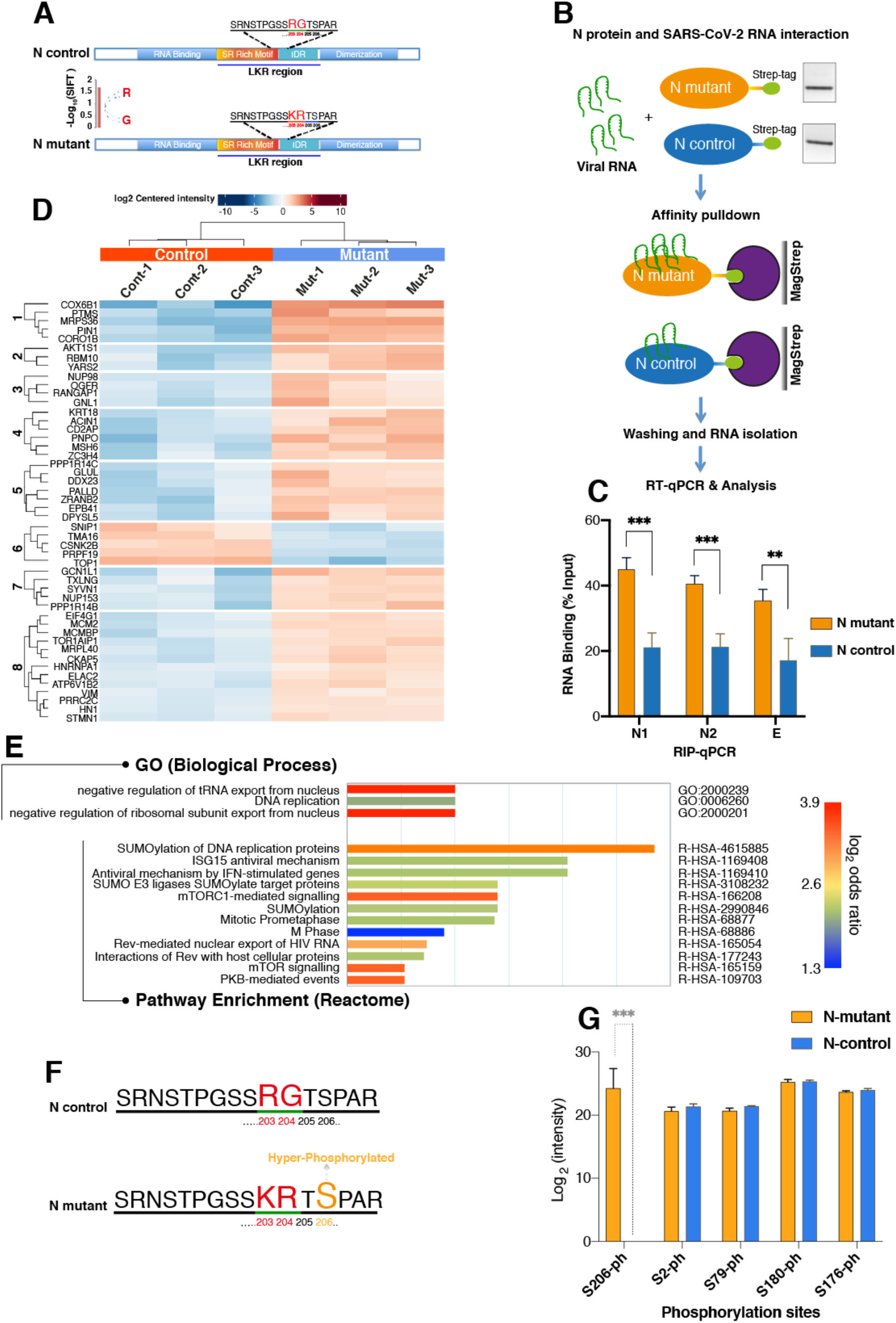
RNA binding and Affinity Purification Mass-Spectrometry (AP-MS) analysis of mutant and control SARS-CoV-2 N protein. A) A schematic diagram showing the SARS-CoV-2 N protein different domains (Upper: control, Lower: mutant) and highlighting the mutation site (R203K and G204R) and the linker region (LKR) containing a serine-arginine rich motif (SR-motif). The bar-plot (lower panel) indicates the SIFT ^29^ predicted deleteriousness score of substitution at position 204 from G to R. B) Sketch of In-vitro RNA immunoprecipitation (RIP) procedure used for analysis of viral RNA interaction with mutant and control N protein (See methods for details). Isolated RNAs were analyzed by RT-qPCR using specific primers for viral N gene (N1 and N2) and E gene. C) Bar chart shows level of viral RNA retrieval (% input) with mutant and control N protein (± SD from 4 experiments, [t-test, p value 0.00016 (***), 0.00019 (***), and 0.003 (**)]). D) Identification of host-interacting partners of mutant and control SARS-CoV-2 N protein by Affinity Mass-Spectrometry. Heatmap showing significantly differentially changed human proteins (3 replicates) interactome in mutant versus control N protein AP-MS analysis. E) Gene Ontology (GO)-enrichment analysis of significantly changed terms between mutant and control proteins in terms of biological process and pathway enrichment. The scale shows p-value adjusted Log2 of odds ration mutant-versus-control. F) Profiling of phosphorylation status of mutant and control N protein by Mass-Spectrometry. Sketch showing part of SR-rich motif of SARS-CoV-2 N protein containing the KR mutation site (R203K and G204R) (Lower). The hyper-phosphorylated serine 206 (as shown in G) in the mutant N protein near the KR mutation site is indicated in orange color. G) Phosphorylation status of mutant and control N protein was analyzed by mass spectrometry (3 biological replicates per affinity condition). Bar-plot shows the Log2 intensities of selected phosphorylated peptides in control and mutant condition. Serine 206 is hyper-phosphorylated in mutant N protein (± SD from 3 experiments, p-value 0.00017 (***) t-test).

Given that the oligomerization of N protein acts as a platform for viral RNA interactions ^35^, we sought to examine the binding affinity of mutant and control N protein with viral RNA isolated from COVID-19 patient swabs. The RNA-binding activity of mutant and control N proteins was examined by pulled-down viral RNA through an *in vitro* RIP assay (Figure 4B), and our data revealed that the mutant N protein enriched significantly higher level of viral RNA compared to control protein (Figure 4C). This indicates a strong binding capability of mutant N proteins with viral RNA, which could potentially impact the essential roles of N protein at various stages of viral life cycle and its interaction with the host.

### The R203K/G204R mutations in the N protein affect its interaction with host proteins

According to the SIFT tool ^29^, a substitution at position 204 from G to R in the N protein is predicted to affect functional properties (Figure 4A). Therefore, we decided to investigate how the two amino acids substitution (R203K and G204R) in the N protein impact its functional interaction with the host that could modulate viral pathogenesis and rewiring of host cell pathways and processes. HEK-293T cells (3 biological replicates, Supplementary Information) were used for affinity-purification followed by mass spectrometry analysis (AP-MS) to identify host proteins associated with the control and mutant N protein (Figure S12). The majority (87%) of non-differentially interacting proteins overlapped with the previously reported^36^ N protein interacting partners (Figure S12D and Table S4). We identified 48 human proteins that displayed significant (adjusted p-value ≤ 0.05, and Log_2_ fold change ≥ 1) differential interactions with the mutant and control N protein (Figure 4D, Figure S12E, Table S5). Among these, 43 proteins showed increased interaction and 5 proteins showed decreased interaction with the N mutant (Figure 4D, Figure S12E). Among the group with increased interaction, we identified many proteins associated with TOR and other signaling pathways (such as AKT1S1 and PIN1), proteins associated with the viral process, viral transcription, and negative regulation of RNA nuclear export (NUP153 and NUP98), and proteins involved in apoptotic and cell death processes (PAWR, ACIN1, and PDCD5) (Figure 4D, Figure S12E). We also identified proteins in the mutant condition that are linked with the immune system processes (PTMS), kinase activity (GCN1), and translation (e.g. MRPS36) (Figure 4D, Figure S12E). In the group with decreased interaction, we identified SNIP1 (NF-kappaB signaling), TMA16 (translation), and CSNK2B (casein kinase II) (Figure 4D, Figure S12E). Gene ontology analysis showed that the most enriched biological processes are associated with negative regulation of tRNA and ribosomal subunit export from the nucleus (Figure 4E). This finding suggests that the mutant virus may more efficiently inhibit and hijack the host translation to facilitate viral replication and pathogenesis. Further, many viruses can manipulate the host sumoylation process to enhance viral survival and pathogenesis^37^. By pathway enrichment analysis of differentially interacting proteins, we identified pathways associated with the sumoylation of host proteins and antiviral mechanisms (Figure 4E).

### Serine 206 (S206) displays hyper-phosphorylation in the mutant N protein

In SARS-CoV, it has been shown that phosphorylation of the N protein is more prevalent during viral transcription and replication ^38^ and inhibition of phosphorylation diminishes viral titer and cytopathogenic effects ^39^. Recent elegant studies elaborated the role of N protein phosphorylation in modulating RNA binding and phase separation in SARS-CoV-2 ^35,40-42^. Thus, phosphorylation of N protein in the LKR region is critical for regulating both viral genome processing (transcription and replication) and nucleocapsid assembly ^35,40^. To further understand the functional relevance of KR mutation in the N protein, we performed phosphoproteomic analysis in control and mutant conditions. We consistently found that the serine 206 (S206) site, which is next to the KR mutation site (Figure 4F), is highly phosphorylated, specifically in the mutant N protein (Figure 4G, Table S6). Notably, the phosphorylation level at serine 2 (S2) and other serine sites (S79, S176, and S180) within the LKR region did not change between mutant and control conditions (Figure 4F).

### The N mutant (R203K/G204R) induces overexpression of interferon related genes in transfected host cells

To understand whether the R203K/G204R mutations in the N gene affect host cell transcriptome, we transfected HEK293T cells with plasmids expressing the full-length N-control and N-mutant protein along with mock-transfection control. The transcriptome profile of N-mutant and N-control transfected cells displays a distinct pattern from the mock-control (Figure S13A). We identified 83 and 67 differentially expressed genes (DEGs) in the N-mutant and N-control transfected cells, respectively, with adjusted *p*-value < 0.05 and log_2_ fold-change ≥ 1 (Figure S13B-C and Table S7). Among the DEGs, numerous interferon, cytokine, and immune-related genes are up-regulated, some of which are shown in Figure 5 (for complete list see Table S8). We found a robust overexpression of interferon-related genes in the N-mutant compared to N-control transfected cells (Figure 5A-B) after adjusting for fold change (Figure S13D). Indeed, strong overexpression of interferon and chemokine related genes (Table S7) were reported in critical COVID-19 patients ^43,44^. Recent reports further indicate a link between increased expression of interferon-related genes and higher viral load in severe COVID-19 patients ^45-47^. Also, we found overexpression of other genes such as ACE2, STAT1 ^44^, and TMPRSS13 ^48^ (Figure 5A and Table S7) that are elevated in critical COVID-19 disease.

**Figure 5.**
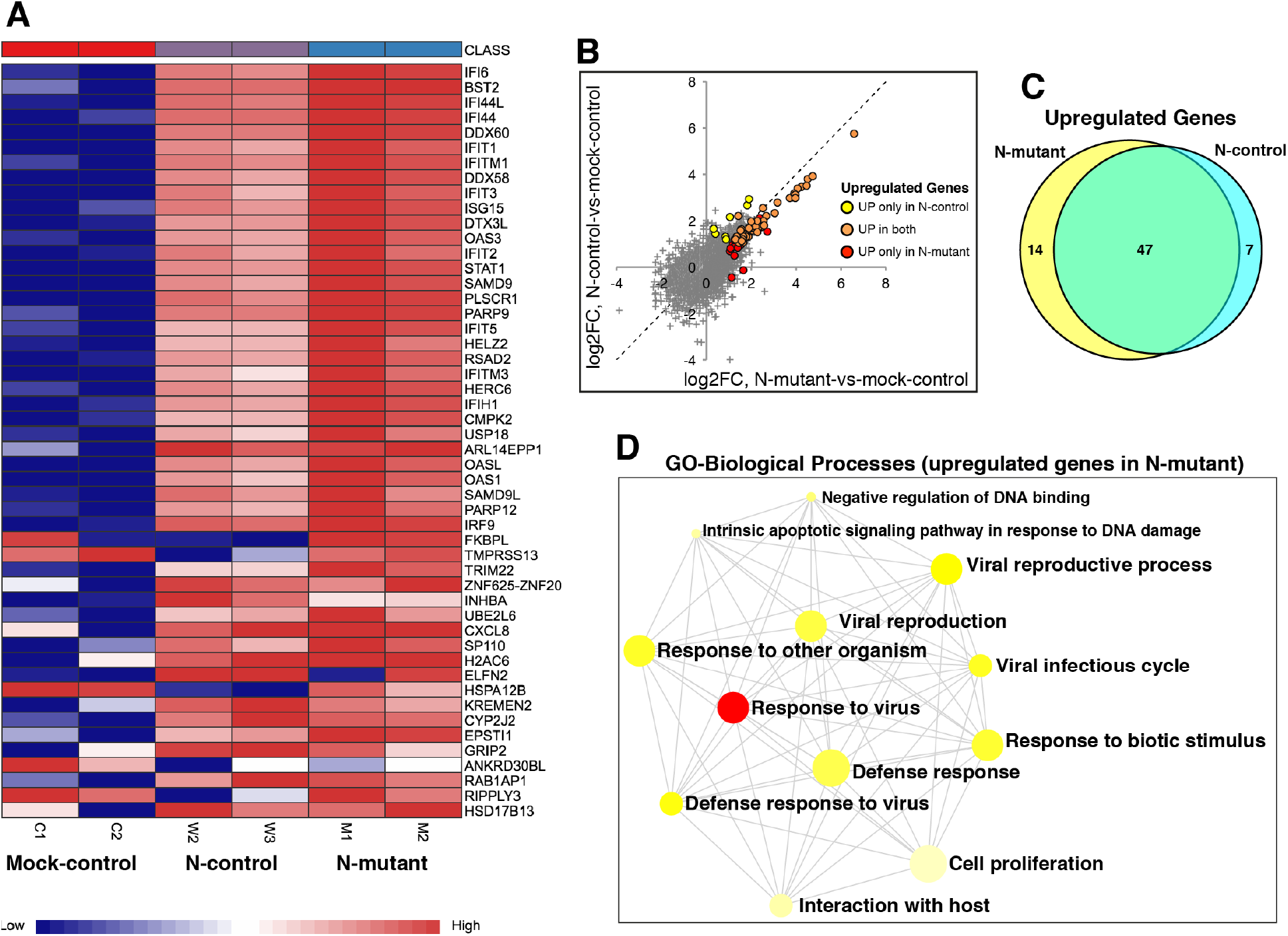
Transcriptional profiling of mutant and control N transfected cells. HEK293T cells were transfected with plasmids expressing the full-length N-control and N-mutant protein along with mock control. 48-hour post-transfection total RNA was isolated and subjected to RNA-sequencing using illumina NovaSeq 6000 platform. A) Heatmap shows normalized expression of top significantly differentially expressed genes in N-mutant and N-control conditions (adj p-value <0.05 and log2 fold-change cutoff ≥1). Genes enriched in interferon and immune related processes are overexpressed in the N-mutant transfected cells. The heatmap was generated by the visualization module in the NetworkAnalyst. B) Plot showing comparison of fold-changes for up-regulated genes in N-mutant and N-control conditions. Differentially expressed genes display higher up-regulation in the N-mutant condition (as orange dots that represent common up-regulated genes are skewed towards the lower half of the diagonal). C) Venn diagram shows the common and unique up-regulated genes in both conditions. D) GO-enrichment analysis of uniquely up-regulated genes in the N-mutant condition. The enriched GO BP (Biological Processes) term is related to interferon response. The enriched terms display an interconnected network with overlapping gene sets (from the list). Each node represents an enriched term and colored by its p-value. The size of each node corresponds to number of linked genes from the list.

Pathway enrichment analysis of the uniquely up-regulated genes in the N-mutant condition (Figure 5C) shows an overrepresentation of biological process pathways associated with response to the virus (Figure 5D). Similarly, all up-regulated genes were related to substantially enriched pathways, such as interferon-related response, cytokine production, and viral reproductive processes (Figure S13E). The enriched GO terms display an interconnected network highlighting the relationships between up-regulated overlapping genes sets in these pathways (Figure 5D and Figure S13E). Taken together, these results suggest that the R203K/G204R mutations in the N protein may enhance its function in provoking a hyper-expression of interferon-related genes that contribute to the cytokine storm in exacerbating COVID-19 pathogenesis.

## Discussion

From 892 samples collected across the country over the course of approximately 6 months we have analyzed the dynamics of transmission and diversity of SARS-CoV-2 in Saudi Arabia. The lineage analysis of assembled genomes highlights the repeated influx of SARS-CoV-2 lineages into the Kingdom. The earliest estimated importation dates point to an entry during the early stages of the pandemic (Figure 2B, Figure 2C), with the first importation likely to have an Asian origin (Figure 2C). From estimates of viral genetic diversity and reproduction rate, we find that decreased diversity and reproduction rate coincides with imposed national curfews and is followed by an observed drop in reported COVID-19 cases (Figure 1D).

Our COVID-19 patient data allowed to us detect three SNPs – underlying the N protein R203K and G204R mutations – significantly associated with higher viral load. It is worth noting that two studies have found higher viral load has in infected patients to be associated with severity and mortality ^49,50^. Among our samples we initially observed an apparent association between the R203K/G204R mutations and mortality, however, the association was no longer statistically significant when correcting for sampling time.

A dated phylogenetic approach suggests that the R203K/G204R mutations arose early in the SARS-CoV-2 pandemic – perhaps even as multiple independent events – and that the mutations entered Saudi Arabia during late January 2020, most likely through Italy.

The N protein of SARS-CoV-2, an abundant viral protein within infected cells, serves multiple functions during viral infection, which besides RNA binding, oligomerization, and genome packaging, playing essential roles in viral transcription, replication, and translation ^30,51^. Also, the N protein can evade immune response and perturbs other host cellular processes such as translation, cell cycle, TGFβ signaling, and induction of apoptosis ^52^ to enhance virus survival. The critical functional regulatory hub within the N protein is a conserved serine-arginine (SR) rich-linker region (LKR), which is involved in RNA and protein binding ^53^, oligomerization ^33,34^, and phospho-regulation ^35,40^.

We show that the mutant N protein containing R203K and G204R changes has higher oligomerization and stronger viral RNA binding ability, suggesting a potential link of these mutations with efficient viral genome packaging. The R203K and G204R mutations are in close proximity to the recently reported RNA-mediated phase separation domain (aa 210–246) ^42^ that is involved in viral RNA packaging through phase separation. This domain was thought to enhance phase-separation also through protein-protein interactions ^42^. Further studies are needed to examine any definite link between KR mutation and phase-separation; however, the differential interaction of host proteins, as shown in our study could affect this process.

Moreover, the functional activities of the N protein at different stages of the viral life cycle are regulated by phosphorylation-dependent physiochemical changes in the LKR region ^40^. Although all individual phosphorylation sites may not be functionally important ^32,54^, the specific enhancement of phosphorylation at serine 206 in the mutant N protein shown in this study hints at its functional significance. The serine 206 can form a phosphorylation-dependent binding site for protein 14-3-3, involved in cell cycle regulatory pathways regulating human and virus protein expression ^55^. Multiple lines of evidence show that N protein phosphorylation is critical for its dynamic localization and function at replication-transcription complexes (RTC), where it promotes viral RNA transcription and translation by recruiting cellular factors ^38-40,56-59^. The enrichment of glycogen synthase kinase 3 A (GSK3A) with the mutant N protein, could specifically phosphorylate serine 206 in the R203K/G204R mutation background. GSK3 was shown to be a key regulator of SARS-CoV replication due to its ability to phosphorylate N protein ^39^. Phosphorylation of serine 206 acts as priming site for initiating a cascade of GSK-3 phosphorylation events ^39,40^. Also, GSK3 inhibition dramatically reduces the production of viral particles and the cytopathic effect in SARS-CoV-infected cells ^39^. Finally, our analysis of the transcriptome in transfected cells suggests that the mutant N protein induce overexpression of interferon-related genes that can aggravate the viral infection by inducing cytokine storm.

As the COVID-19 pandemic is still ongoing, there is a need for novel therapeutic strategies to treat severe infections in patients. Our identified interaction pathways and inhibition of serine 206 phosphorylation could be used as potential targets for therapies.

In conclusion, our results highlight the major influence of the R203K/G204R mutations on the essential properties and phosphorylation status of SARS-CoV-2 N protein that lead to increased host response and efficacy of viral infection.

## Methods

### Sample Collection

As part of the study, nasopharyngeal swab samples were collected in 1ml of TRIzol (Ambion, USA) from 892 COVID-19 patients with various grades of clinical disease manifestations – consisting of severe, mild and asymptomatic symptoms. The anonymized samples were amassed from 8 hospitals and one quarantine hotel located in Madinah, Makkah, Jeddah and Riyadh. Ethical approvals were obtained from the Institutional review board of the Ministry of Health in Makkah region with the numbers H-02-K-076-0420-285 and H-02-K-076-0320-279, as well as the Institutional review board of Dr. Sulaiman Al Habib Hospital number RC20.06.88 for samples from Riyadh and the Eastern regions respectively.

### RNA Isolation

RNA was extracted using the Direct-Zol RNA Miniprep kit (Zymo Research, USA) following the manufacturer’s instructions, along with several optimization steps to improve quality and quantity of RNA from clinical samples. The optimization included extending the TRIzol incubation period, and the addition of chloroform during initial lysis step to obtain the aqueous RNA layer. The quality control of purified RNA was performed using Broad Range Qubit kit (Thermo Fisher, USA) and RNA 6000 Nano LabChip kit (Agilent, USA) respectively. RT-PCR was conducted using the one-step Super Script III with Platinum Taq DNA Polymerase (Thermo Fisher, USA) and TaqPath COVID-19 kit (Applied Biosystems, USA) on the QuantStudio 3 Real-Time PCR instrument (Applied Biosystems, USA) and 7900 HT ABI machine. The primers and probes used were targeting two regions in the nucleocapsid gene (N1 and N2) in the viral genome following the Centre for Disease Control and prevention diagnostic panel, along with primers and probe for human RNase P gene (CDC; fda.gov/media/134922/download). Samples were considered COVID positive once the cycle threshold (Ct) values for both N1 and N2 regions were less than 40. For amplicon seq purposes, the samples chosen were of Ct less than 35 to ensure successful genome assembly in order to upload on GISAID.

### Sequencing and Data analysis

cDNA and amplicon libraries were prepared using the COVID-19 ARTIC-V3 protocol, producing ∼ 400bp amplicons tiling the viral genome using V3 nCoV-2019 primers (Wellcome Sanger Institute, UK; dx.doi.org/10.17504/protocols.io.beuzjex6). Amplicons were then processed for deep, paired-end sequencing with the Novaseq 6000 platform on the SP 2 x 250 bp flow cell type (Illumina, USA).

### Genome assembly, SNP and indel calling

Illumina adapters and low quality sequences were trimmed using Trimmomatic v0.38 ^60^. Reads were mapped to SARS-CoV-2 Wuhan-Hu-1 NCBI reference sequence NC_045512.2 using BWA ^61^. Mapped reads were processed using GATK v 4.1.7 pipeline commands MarkDuplicatesSpark, HaplotypeCaller, VariantFiltration, SelectVariants, BaseRecalibrator, ApplyBQSR, and HaplotypeCaller to identify variants ^62^.

High quality SNPs were filtered using the filter expression:

“QD<2.0 || FS > 60.0 || SOR > 3.0 || MQRankSum < -12.5 || ReadPosRankSum < -8.0” High quality Indels were filtered using the filter expression:

“QD<2.0 || FS > 200.0 || SOR > 10.0 || ReadPosRankSum < -20.0”

Consensus sequences were generated by applying the good quality variants from GATK on the reference sequence using bcftools consensus command ^63^. Regions which are covered by less than 30 reads are masked in the final assembly with ‘N’s.

Consensus assembly sequences were deposited to GISAID (Table S1) ^11^. To retrieve high-confidence SNPs assembled sequences were re-aligned against the Wuhan-Hu-1 reference sequence (NC_045512.2), and only positions in the sample sequences with unambiguous bases in a 7-nucleotide window centred around the SNP position were kept for further analysis.

### Phylogenetic analysis

To generate the phylogeny of Saudi samples with a global context, a total of 308,012 global sequences were downloaded from GISAID on 31 December 2020, filtered and processed using Nextstrain pipeline^12^. Global sequences were grouped by country and sample collection month and 20 sequences per group were randomly sampled which resulted in 10,873 global representative sequences and 952 Saudi sequences. The phylogeny was constructed using IQ-TREE^64^, clades were assigned using Nextclade and internal node dates were inferred and sequences pruned using TreeTime^65^. Nextstrain protocol was followed for the above-mentioned steps. The resulting global phylogenetic tree was reduced to retain the branches that lead to Saudi leaf nodes and visualised using baltic library (https://github.com/evogytis/baltic).

### Phylodynamic analysis

Phylodynamic analyses use the same sequence subset used in the full phylogenetic analysis, extracted from the GISAID SARSCoV-2 database ^11^. Wrapper functions for the importation date estimates and skygrowth model are provided in the sarscov2 R package as ‘compute_timports’ and ‘skygrowth1’ respectively (github.com/emvolz-phylodynamics/sarscov2Rutils).

### Importation date estimates for Nextstrain clades

Sequences corresponding to each Nextstrain ^12^ clade were extracted using the Nextstrain_clade parameter in the GISAID metadata table. A subset of 500 international sequences were select for each clade based on Tamura Nei 93 distance with tn93 (github.com/veg/tn93) and stratified over time^66^. A maximum likelihood phylogeny with an HKY substitution model for each clade was estimated with IQtree^64,67^. Time-scaled phylogenies were estimated from this using treedater with a strict molecular clock constrained between 0.0009 and 0.0015 substitutions per site per year^68^. 15 Variations of each dated phylogeny were produced by collapsing small branches and resolving polytomies. The state of each internal node was reconstructed by maximum parsimony with the phangorn R package^69^. Importation events are estimated at the midpoint of a branch along which a location change is inferred to occur by this method.

### Estimation of donor countries behind importation events

To identify import events that resulted in new introductions into Saudi Arabia, 25,198 sequences were subsampled from 590K global sequences available on GISAID on February 24th 2021. Samples with closer genetic distance to Saudi Samples were preferred. The phylogeny was constructed using IQ-TREE^64^, internal nodes dates and possible country for internal nodes were inferred using TreeTime^65^. Nextstrain protocol was followed for the above-mentioned steps^12^. In house scripts were used to traverse the global phylogenetic tree to identify branches that resulted in transitions into Saudi Arabia from another country.

### Skygrowth model

Sequences from Saudi Arabia available on GISAID on December 31^st^ 2020 were used to construct effective population size and growth rate of SARS-CoV2 in Saudi Arabia over the course of the first wave of the epidemic (March to September 2020). As with the importation date estimates, a maximum likelihood phylogeny was produced, time-scaled and variation introduced by resolving polytomies to give a sample of 15 phylogenies.

We modelled growth rate and effective population size over time on these phylogenies using the R package skygrowth^16^. Skygrowth is a non-parametric Bayesian approach which applies a stochastic process on estimates of growth rate and effective population size. The model included mean-centred, unit variance estimates of travel rates from google mobility data (google.com/covid19/mobility/) as a covariate (transit stations percent change from baseline), 60 timesteps and a tau (precision) value corresponding to a 1% change in growth per week. The growth rate output was converted to an estimate of R over time using an infectious period of 9.5 days^70^.

### Origin of R203K/G204R SNPs

A total of 590K samples submitted to GISAID until February 24 were downloaded and SNPs indentifed by mapping against the Wuhan reference using minimap2^71^. The variants were queried to count the distribution of triplets among various Nextstrain clades (Figure S7). To identify if there are lineages of triplet SNPs in clades other than 20B, a phylogenetic tree was constructed by including all R203K/G204R samples found in other clades outside 20B and its subclades (Figure S4). As it was already evident that 20B and its subclades contains lineages of R203K/G204R samples, subsamples from 20B and its subclades were sufficient to obtain a total of 16,386 samples.

### Statistical analysis

Statistical analyses were performed with the statistical software R version 4.0.3 ^72^ and the R package mgcv version 1.8.33. Model with response (log_10_ copynumber) used 473 observations (samples processed with the TaqPath kit). Models for mortality and severity used 892 observations (all data).

### Plasmid and cloning

The pLVX-EF1alpha-SARS-CoV-2-N-2xStrep-IRES-Puro was a gift from Nevan Krogan (Addgene plasmid # 141391; http://n2t.net/addgene:141391; RRID:Addgene_141391)^36^. The three consecutive SNPs (G28881A, G28882A, G28883C), corresponding to N protein mutation sites R203K and G204R, were introduced by megaprime PCR mutagenesis using the primers listed in Table S8.

### Cell culture and transfection

HEK293T (ATCC; CRL-3216) cells were grown in Dulbecco’s modified Eagle’s medium (DMEM) (4.5 g/l d-glucose and Glutamax, 1 mM sodium pyruvate) (GIBCO) and 10% fetal bovine serum (FBS; GIBCO) with penicillin–streptomycin supplement, according to standard protocols (culture condition 37 °C and 5% CO2). Transfection of ten million cells per 15-cm dish with 2XStrep-tagged N plasmid (20ug/transfection) was performed using lipofectamine-2000 according standard protocol.

### Affinity purification and on-bead digestion

Cell lysis and affinity purification with MagStrep beads (IBA Lifesciences) was manually performed according to the published protocol^36^ with minor modifications. Briefly, after transfection (48 hours) cells were collected with 10mM EDTA in 1xPBS and washed twice with cold PBS (1x). The cell pellets were stored at -80 °C. Cells were lysed in lysis buffer (50 mM Tris-HCl, pH 7.4, 150 mM NaCl, 1 mM EDTA, 0.5% NP40, supplemented with protease and phosphatase inhibitor cocktails) for 30 minutes while rotating at 4 °C and then centrifuge at high speed to collect the supernatant. The cell lysate was incubated with prewashed MagStrep beads (30 µl per reaction) for 3 hours at 4 °C. The beads were then washed four times with wash buffer (50 mM Tris-HCl, pH 7.4, 150 mM NaCl, 1 mM EDTA, 0.05% NP40, supplemented with protease and phosphatase inhibitor cocktails) and then proceed with on-bead digestion. The on-bead digestion was carried out as described before^36^. For affinity confirmation, bound proteins were eluted using buffer BXT (IBA Lifesciences) and after running on SDS-PAGE were subjected to silver staining and western-blot using anti-strep-II antibody (ab76949). To purify clean 2xStrep-tagged N protein (mutant and control), we applied stringent washing and double elution strategy.

### MS analysis using Orbitrap Fusion Lumos

The MS analysis was performed as described previously ^73,74^ with slight modifications. For mass spectrometry analysis an Orbitrap Fusion mass spectrometer (MS) (Lumos, Thermo Fisher Scientific) was used in data-dependent acquisition (DDA) mode. For injection, 0.5 µg peptide mixture was used and desalting was performed for 5 minutes in 0.1% FA in water. The gradient and all other steps were essentially the same as described^73^.

Protein identification analysis from the raw mass spectrometry data was performed using the Maxquant software (version 1.5.3.30)^75^ as described^73^.

For phosphorylated peptides, we used Maxquant label-free quantification (LFQ)^***75***^. The analysis and quantification of phosphorylated peptides was performed according to published protocol^***76***^.

### Analysis of differential interaction

The normalized LFQ data were processed for statistical analysis on the LFQ-Analyst a web-based tool^***77***^ to performed pair-wise comparison between mutant and control N protein AP-MS data. The significant differentially changed proteins between mutant and control conditions were identified. The threshold cut-off of adjusted p-value <= 0.05, and Log fold change >= 1 were used. Among the replicates, outliers were removed based on correlation and PCA analysis. The GO enrichment analysis was performed on the LFQ-Analyst^***77***^.

### BS3 cross-linking

*Bis(sulfosuccinimidyl) suberate (BS3, Thermo Scientific Pierce) was used for* cross-linking of control and mutant N protein to analyse the oligomerization properties. The experiment was performed as reported previously^*78*^.

### RNA-sequencing and differential gene expression analysis

HEK293T cells were transfected with plasmids expressing the full-length N-control and N-mutant protein along with mock control. After 48-hour cells were harvested in Trizol and total RNA was isolated using Zymo-RNA Direct-Zol kit (Zymo, USA) according to the manufacture’s instruction. The concentration of RNA was measured by Qubit (Invitrogen), and RNA integrity was determined by Bioanalyzer 2100 system (Agilent Technologies, CA, USA). The RNA was then subjected to library preparation using Ribozero-plus kit (Illumina). The libraries were sequenced on NovaSeq 6000 platform (Illumina, USA) with 150 bp paired-end reads.

The raw reads from HEK293T RNA-sequencing were processed and trimmed using trimmomatic ^60^ and mapped to annotated ENSEMBL transcripts from the human genome (hg19) ^79,80^ using kallisto^81^. Differential expression analysis was performed after normalization using EdgeR integrated in the NetworkAnalyst ^82^. GO biological process and pathway enrichment analyses on up-regulated genes were performed using NetworkAnalyst ^82^.

## Data Availability

Assembled virus genomes are available at GISAID (Table S1). Reads from RNA-Seq analysis of transfected HEK293T cells have been uploaded to European Nucleotide Archive (https://www.ebi.ac.uk/ena/) under the Study accession number PRJEB44716

## Acknowledgments

We are sincerely grateful to all hospital members for providing samples and collating metadata in such an unprecedented pandemic, along with the MOH and ethical committee, which rendered it permissible. We thank the KAUST Rapid Research Response Team (R3T) under the Vice President – Research (VPR) office in KAUST for generous financial support. We also thank Erik Talley from KAUST Health Safety and Environment (HSE) and Hani Bukhari from KAUST Security for providing timely logistical support for samples transport during COVID-19 Curfew restrictions in the Kingdom.

We extend our thanks and appreciation to GDRS director, PH. Athari Alotaibi (General Director for Research and Studies, MOH) for her vigorous facilitation of the research project, and Mohammad Fawzi (General Directorate of Health Affairs) for his help with the metadata collection. We also deeply thank Dr. Wael Hamzah Motair, Dr. Nader Hamzah Motair, Dr. Hatim Khogeer and the General Directorate of Health Affairs of Makkah Region (GDHAMR), MOH for all their help and assistance to our study.

We gratefully acknowledge all of the authors from the originating laboratories responsible for obtaining the specimens and the submitting laboratories where genetic sequence data were generated and publicly shared via the GISAID Initiative, on which was partially used for additional support for some of the conclusions drawn in this study.

## Grants

KAUST Rapid Research Response Team (R3T) by Vice President – Research (VPR) office in KAUST.

KAUST faculty baseline fund (BAS/1/1020-01-01) to AP.

KACST Grants, Proposal number: 5-20-01-002-0008

MOH COVID-19 project grants number 341

MOH COVID-19 project grants number 754

The deputyship for Research and Innovation, Ministry of Education in Saudi Arabia, project number (436) to AMH.

## IRBs

This project was conducted under the IRB approvals of the MOH (H-02-K-076-0420-285), KAUST (20IBEC14) and at the Dr. Suliman Al-Habib Medical group (HAP-01-R-082) in KSA.

## Author Contributions

A.P. conceived the study and directed the work and acquired funding from KAUST and supplemental funding from King Abdulaziz City for Science and Technology (KACST). A.P., T.M., M.S., S.H., and S.M. designed the research. IRB and ethical approvals from MOH were acquired by A.K., A.H., N.A., A.M., and S.H. to cover the collection from several cities in the Kingdom. S.H. and A.K. acquired funding from the Saudi Ministry of Health (MOH) numbers 754 and 341, utilized in the study. S.H. organized and directed sample collection and metadata collections with aid from F.A., A.S., A.O., S.S., J.T., A.A., N.K., K.K., K.A., and A.D.; S.M. directed the wet lab work involving sample reception, metadata record-keeping, RNA extraction, quality control, and library preparation, with aid from A.S., F.B., R.S., M.S., A.O., L.E., O.D., S.H. and R.N. Illumina sequencing runs, Mass spectrometry and raw data processing were performed by S.M., L.E., S.P., and I.R. respectively. Genome assemblies and submission to GISAID was done by R.N. Phylogenetic and lineage analysis was done by R.N., Q.G., D.J., and E.V. In-depth SNP data analysis was performed by T.M. Statistical analysis done by P.E.M. and E.V. Functional validation of this link was established by M.S.; T.M. wrote the initial draft of the manuscript with input from M.S., S.M., S.H., R.N., and Q.G., followed by edits from A.P. The final version was produced by T.M., M.S. and A.P. after input from all co-authors.

## Data availability

Supplementary Information is available for this paper. Assembled virus genomes are available at GISAID (Table S1). Reads from RNA-Seq analysis of transfected HEK293T cells have been uploaded to European Nucleotide Archive (https://www.ebi.ac.uk/ena/) under the Study accession number PRJEB44716.

Correspondence and requests for materials should be addressed to Arnab Pain.

## Notes

### Competing Interest Statement

The authors have declared no competing interest.

### Summary of Updates

Supplemental files updated

