## Supplementary Information, Tables S2-S8 for "Saudi Arabian SARS-CoV-2 genomes implicate a mutant Nucleocapsid protein in modulating host interactions and increased viral load in COVID-19 patients"

**Daily Cases in Saudi Arabia**

The numbers of Covid-19 cases registered in Saudi Arabian Cities were collected from The Ministry of Health Command Centre for COVID-19 (https://covid19.moh.gov.sa), Saudi Center of Disease Control and Prevention (https://covid19.cdc.gov.sa/), the Saudi Press Agency (SPA) (https://www.spa.gov.sa/search.php?lang=en&search=COVID), The Saudi Ministry of Interior (https://www.moi.gov.sa/), and Algaissi *et al.*[1].

**Polymorphisms**

Duplicate reads were removed from the mapped short Illumina sequence reads using picard tools' MarkDuplicates function (Broad Institute, GitHub repository. http://broadinstitute.github.io/picard/). Apparent mismatches were collected from the output of samtools mpileup[2]. Distal read positions were excluded and only read mapping and base calling qualities of at least 30 were considered. Positions with at a minimum coverage of 1000 and with an overall mismatch rate between 0.3 and 0.7 were considered as potential within-host polymorphic sites.

For each hospital, the number of samples with detected polymorphism at a given SNP site was plotted against the number of samples from the hospital that had this SNP in the assembled genome and the correlation was calculated. Examples of such plots for six different SNPs are shown in Figure S9.

We considered the frequency of a given SNP in the assembled virus genomes from a hospital as a proxy for the prevalence of this virus variant at the hospital. We further consider the probability of seeing polymorphisms of a given SNP due to co-infection as a product of the frequency of viruses with that SNP circulating among patients (the more prevalent a virus strain is in a given hospital environment the more likely it is to co-infect other COVID19 patients). Therefore, if observed polymorphisms were the result of co-infections we would expect to see a positive correlation between SNP polymorphisms and SNP frequencies at different hospitals. In contrast, if observed polymorphisms were the result of inter-sample contaminations, such a correlation is not expected, as contamination would solely rely on the order and workflow by which the samples were handled.

The observed positive correlation between SNP polymorphisms and SNPs in assembled genomes between hospitals (Figure S9) is therefore consistent with the observed polymorphisms being the result of co-infection of patients. And we similarly consider this to be the case for the R203K/G204R SNPs.

### Ct values

Although not associated with severity of infection, significantly lower Ct values have been reported for the D614G SNP [3-5]. In our experimental setup, samples were not only selected for further processing based on the initially obtained Ct values, but sample qPCRs were also run in separate batches, which produced separate standard curves. Additionally, two different laboratory kits were used for the qPCRs (see Methods). Furthermore, viral loads will also be a function of sampling time, with fluctuations over the course of infection and potential recovery. With these caveats in mind, we nevertheless tested for differences in Ct values between samples. This was done using samples processed with the TaqPath kit (see Methods). Hence, to prove that that the R203K/G204R SNPs result in higher viral loads, experimental *in vitro* infection studies with different virus genotypes are needed.

### Table S2. Cities providing samples

| City | Total samples | R203K/G204R frequency |
|---|---|---|
| Madinah | 312 | 0.0321 |
| Makkah | 231 | 0.0303 |
| Jeddah | 262 | 0.3855 |
| Riyadh | 69 | 0.0870 |
| Eastern Region | 18 | 0 |
| **TOTAL** | **892** | **0.1390** |

### Table S3. Detected indels

| position | type | length | ref | allele | sample count |
|---|---|---|---|---|---|
| 103 | del | 2 | CTG | C | 2 |
| 509 | del | 9 | GGTCATGTTA | G | 1 |
| 668 | del | 3 | AGTT | A | 3 |
| 685 | del | 9 | AAAGTCATTT | A | 4 |
| 898 | ins | 262 | TTCATGCACTTTGTCCGAACAACTGGACTTTATTGACACTAAGAGGGGTGTATACTGCTGCCGTGAAC | TTCATGCACTTTGTCCGAACAACTGGACTTTATTGACACTAAGAGGGGTGTATACTGCTGCCGTGAACCACTTTTTCTTTGCATTTACTTTTTTATAGGAACTCCTGTCATCACTCTCTCACACACACACTTAGATGAACCTGATGGCTACCCTCTTGAGTGCATTAAAGACCTTCTAGCACGTGCTGGTAAAGCCTCATGCACTTTGTCCGAACAACTGGACTTTATTGACACTAAGAGGGGTGTATACTGCTGCCGTGAAC | 1 |
| 2463 | del | 4 | TAAAA | TAA | 1 |
| 2628 | del | 3 | TTAT | TT | 1 |
| 2882 | ins | 282 | TGTGTTGTGGCAGATGCTGTC | TGTGTTGTGGCAGATGCTGTCGTGTTGTGGCAGATGCTGTCGTGTTGTGGCAGATGCTGTCGTGTTGTGGCAGATGCTGTCGTGTTGTGGCAGAGGCTGCTCGTGTTGTGGCAGATGCTGTCGTGTTGTGGCAGATGCTGTCGTGTTGTGGCAGATGCTGTAGTGTTGCATCAGAGGCTGCTCGTGTTGTGGCAGATGCTGTCGTGTTGTGGCAGATGCTGTCGTGTTGTGGCAGATGCTGTCGTGTTGTGGCAGATGCTGTCGTGTTGTGGCAGATGCTGTC | 1 |
| 2882 | ins | 141 | TGTGTTGTGGCAGATGCTGTC | TGTGTTGTGGCAGATGCTGTCGTGTTGTGGC | 1 |

| | | | | | |
|---|---|---|---|---|---|
| | | | | AGATGCTGTCGTGTTGTGGCAGATGCTGTCG<br>TGTTGTGGCAGATGCTGTCGTGTTGTGGCAG<br>ATGCTGTCGTGTTGCATCAGAGGCTGCTCGT<br>GTTGTGGCAGATGCTGTC | |
| 7626 | ins | 49 | ATTGTGATACATTCTGTGCTGGTAGT | ATTGTGATACATTCTGTGCTGGTAGTTGTGA<br>TACATTCTGTGCTGGTAGT | 2 |
| 10535 | del | 27 | TACATGCACCATATGGAATTACCAACTG | T | 1 |
| 11074 | ins | 11 | CTTTTTTTT | CTTTTTTTTTTT | 1 |
| 18896 | ins | 309 | TTGTTAAGCGTGTTGACTGGACTATTGAAT<br>ATCCTATAATTGGTGATGAACTGAAGATTA<br>ATGCGGCTTGTAGAAAGGTTCAACA | TTGTTAAGCGTGTTGACTGGACTATTTAATAT<br>CCTATAATTGGTGATGAACTGAAGATTAATG<br>CGGCTTGTAGAAAGGTTCAACATAACATGTT<br>GTGCCAACCACCAGCACTCCTGGGACCTCC<br>ACAGTGCACCTGGCAACCTCTGGGACTCCAT<br>CCTCCCTGCCTGGCCACACAGCCCCTGTCC<br>CTCTCTTGATACCATTCACCCTCAACTTTACC<br>AGATGGGAATGTTAAGCGTGTTGACTGGACT<br>ATTGAATATCCTATAATTGGTGATGAACTGAA<br>GATTAATGCGGCTTGTAGAAAGGTTCAACA | 1 |
| 19517 | del | 3 | TGTA | T | 1 |
| 21066 | del | 5 | TAAAAA | TAA | 1 |
| 21561 | del | 2 | CAA | CA | 2 |
| 21624 | del | 33 | GAACTCAATTACCCCCTGCATACACTAATT<br>CTTT | G | 1 |
| 21740 | del | 45 | TCCAATGTTACTTGGTTCCATGCTATACAT<br>GTCTCTGGGACCAATG | TCCAATG | 1 |
| 21781 | ins | 3 | CAA | CAAA | 1 |
| 21990 | del | 6 | TTTATTA | TTTA | 3 |
| 22048 | del | 3 | TGCG | T | 1 |
| 22288 | del | 6 | TGCTTTA | T | 1 |
| 22353 | del | 7 | CTTATTAT | CTTAT | 1 |

| | | | | | |
|---|---|---|---|---|---|
| 23701 | del | 1 | CA | C | 1 |
| 27263 | del | 29 | CTTTTAAAGTTTCCATTTGGAATCTTGATT | CTT | 1 |
| 27694 | del | 8 | TTTCTTATT | TTT | 1 |
| 27697 | del | 5 | CTTATT | CTT | 4 |
| 28949 | del | 11 | AGATTGAACCAG | A | 1 |
| 29727 | del | 22 | TTTCACCGAGGCCACGCGGAGTA | T | 1 |
| 29755 | del | 1 | GA | G | 2 |
| 29774 | del | 18 | CTAGGGAGAGCTGCCTATA | CTA | 3 |
| 29865 | ins | 5 | AA | AAAACA | 1 |
| 29865 | ins | 7 | AA | AAACAACA | 4 |
| 29865 | ins | 3 | AA | AACA | 1 |
| 29865 | ins | 9 | AA | AACCACAACA | 1 |
| 29865 | ins | 10 | AA | AAGCCACAACA | 1 |
| 29866 | ins | 3 | A | AACC | 1 |
| 29866 | ins | 6 | A | AAGATGC | 1 |
| 29866 | ins | 9 | A | AAGCAGCCTC | 1 |
| 29866 | ins | 3 | A | AATG | 1 |
| 29866 | ins | 8 | A | AGCAGATGC | 1 |

### Table S4. Nucleocapsid N (control and mutant) interacting partners with no significant change in interaction

| Gene Name | Protein IDs | Gene Name | Protein IDs | Gene Name | Protein IDs | Gene Name | Protein IDs | Gene Name | Protein IDs | Gene Name | Protein IDs |
|---|---|---|---|---|---|---|---|---|---|---|---|
| PRPF38B | Q5VTL8 | CSNK2A1 | P68400 | GNB2L1 | P63244 | CARHSP1 | Q9Y2V2 | RAVER1 | A0A087WZ13 | RPS27 | P42677 |
| PRPF38A | Q8NAV1 | CSNK2A2 | P19784 | GNL3 | Q9BVP2 | LUC7L | Q9NQ29 | RBM14 | Q96PK6 | RPS27A | P62979 |
| TMA7 | Q9Y2S6 | CSRP2 | Q16527 | GRWD1 | Q9BQ67 | LUC7L2 | Q9Y383 | RBM17 | Q96I25 | RPS28 | P62857 |
| TBL2 | Q9Y4P3 | PRPF40A | O75400 | GSK3B | P49841 | LUC7L3 | O95232 | RBM28 | Q9NW13 | RPS29 | P62273 |
| ARL6IP4 | Q66PJ3 | FAM32A | Q9Y421 | EMD | P50402 | PFN1 | P07737 | RCC2 | Q9P258 | RPS3 | P23396 |
| ATXN2L | Q8WWM7 | IBA57 | Q5T440 | GTPBP4 | Q9BZE4 | FXR1 | P51114 | RBM39 | Q14498 | RPS3A | P61247 |
| BAG2 | O95816 | PES1 | O00541 | GNL2 | Q13823 | FXR2 | P51116 | RP9 | Q8TA86 | RPS4X | P62701 |
| UQCRC2 | P22695 | RPA3 | P35244 | HADHA | P40939 | PDCD11 | Q14690 | RPL10 | P27635 | RPS5 | P46782 |
| BCLAF1 | Q9NYF8 | SNIP1 | Q8TAD8 | GRN | P28799 | MAT2A | P31153 | RPL10A | P62906 | RPS6 | P62753 |
| FTSJ3 | Q8IY81 | DDX18 | Q9NVP1 | CDKN2A | Q9NXV6 | MATR3 | P43243 | RPL11 | P62913 | RPS7 | P62081 |
| ARGLU1 | Q9NWB6 | DDX21 | Q9NR30 | KTN1 | Q86UP2 | MAZ | P56270 | RPL12 | P30050 | RPS8 | P62241 |
| STAU1 | Q5JW28 | DDX46 | A0A0C4DG89 | PPP1CB | P62140 | ZCCHC17 | Q9NP64 | RPL13 | P26373 | RPS9 | P46781 |
| AIP | O00170 | DDX47 | Q9H0S4 | HLA-C | P10321 | MFAP1 | P55081 | RPL14 | P50914 | RPSA | P08865 |
| BRIX1 | Q8TDN6 | DDX5 | P17844 | HMGB1 | P09429 | MKI67 | P46013 | RPL15 | P61313 | RRP12 | Q5JTH9 |
| NTPCR | Q9BSD7 | DDX56 | Q9NY93 | HMGB2 | P26583 | PRPF19 | Q9UMS4 | RPL18 | Q07020 | RRP15 | Q9Y3B9 |
| BYSL | Q13895 | DEK | P35659 | PHF5A | Q7RTV0 | CWC25 | Q9NXE8 | RPL18A | Q02543 | RRP1B | Q14684 |
| NOP16 | Q9Y3C1 | DHX15 | O43143 | TUBB8 | Q3ZCM7 | MPG | P29372 | RPL19 | P84098 | RRP9 | O43818 |
| PIN1 | Q13526 | DHX30 | Q7L2E3 | HNRNPA0 | Q13151 | EFTUD2 | Q15029 | RPL21 | P46778 | RRS1 | Q15050 |
| CACYBP | Q9HB71 | DHX9 | Q08211 | HNRNPA2B1 | P22626 | PRPF8 | Q6P2Q9 | RPL22 | P35268 | RSL1D1 | O76021 |

| | | | | | | | | | | | |
|---|---|---|---|---|---|---|---|---|---|---|---|
| EIF5B | O60841 | DDX6 | P26196 | HNRNPA3 | P51991-2 | ZMAT2 | Q96NC0 | RPL23 | P62829 | RSRC2 | Q7L4I2 |
| TOP1 | P11387 | DKC1 | O60832 | HNRNPAB | Q99729 | MRPL14 | Q6P1L8 | RPL23A | P62750 | RTCB | Q9Y3I0 |
| CBR1 | P16152 | DDX1 | Q92499 | HNRNPC | G3V4C1 | MRPS12 | O15235 | RPL24 | P83731 | RTTN | Q86VV8 |
| CBX3 | Q13185 | YTHDC2 | Q9H6S0 | HNRNPF | P52597 | MRTO4 | Q9UKD2 | RPL27 | P61353 | RUVBL1 | Q9Y265 |
| CCDC124 | Q96CT7 | ACACA | Q13085 | HNRNPH1 | P31943 | MSN | P26038 | RPL27A | P46776 | RUVBL2 | Q9Y230 |
| HNRNPA1 | P09651 | DUT | P33316 | HNRNPK | P61978 | MTHFD1 | P11586 | RPL28 | P46779 | SAE1 | Q9UBE0 |
| CCT2 | P78371 | HMGA1 | 17096 | HNRNPL | P14866 | MYBBP1A | Q9BQG0 | RPL29 | P47914 | SAFB | Q15424 |
| CCT3 | P49368 | C8orf33 | Q9H7E9 | HNRNPM | P52272 | MYH9 | P35579 | RPL3 | P39023 | SEC61B | P60468 |
| CCT4 | P50991 | LSM14A | Q8ND56 | HNRNPR | O43390 | NCL | P19338 | RPL30 | P62888 | SERBP1 | Q8NC51 |
| CCT5 | P48643 | EBNA1BP2 | Q99848 | HNRNPU | Q00839 | NAT10 | Q9H0A0 | RPL31 | P62899 | SET | Q01105 |
| CCT6A | P40227 | EEF1G | P26641 | HNRNPUL1 | Q9BUJ2 | NHP2 | Q9NX24 | RPL32 | P62910 | SF1 | Q15637 |
| CCT7 | Q99832 | EEF2 | P13639 | HNRNPUL2 | Q1KMD3 | ABCF1 | Q8NE71 | RPL34 | P49207 | SF3B1 | O75533 |
| CCT8 | P50990 | EIF1AX | P47813 | HP1BP3 | Q5SSJ5 | NOC3L | Q8WTT2 | RPL35 | P42766 | SF3B2 | Q13435 |
| ATOX1 | O00244 | DDX54 | Q8TDD1 | SUPT16H | Q9Y5B9 | NONO | Q15233 | RPL35A | P18077 | SF3B3 | Q15393 |
| EIF3G | O75821 | DDX52 | Q9Y2R4 | HSP90AA1 | P07900 | NOP2 | P46087 | RPL36 | Q9Y3U8 | SFPQ | P23246 |
| LTV1 | Q96GA3 | EIF3B | P55884 | ILF2 | Q12905 | NOP56 | O00567 | RPL37A | P61513 | SLC25A3 | Q00325 |
| CFL1 | P23528 | DDX50 | Q9BQ39 | WDR1 | Q9BV38 | NOP58 | Q9Y2X3 | RPL38 | P63173 | SLC25A5 | P05141 |
| TXNDC12 | O95881 | EIF3D | O15371 | HSPA4 | P34932 | NOSIP | Q9Y314 | RPL4 | P36578 | SLC25A6 | P12236 |
| CHERP | Q8IWX8 | DDX24 | Q9GZR7 | HSPA4L | O95757 | NPM1 | P06748 | RPL5 | P46777 | SNRNP27 | Q8WVK2 |
| SNRPD2 | P62316 | EIF4A1 | P60842 | HSPA5 | P11021 | NUMA1 | Q14980 | RPL6 | Q02878 | SNRPA1 | P09661 |
| CHTOP | Q9Y3Y2 | EIF4A3 | P38919 | HSPA6 | P17066 | PA2G4 | Q9UQ80 | RPL7 | P18124 | SNRPD3 | P62318 |

| | | | | | | | | | | | |
|---|---|---|---|---|---|---|---|---|---|---|---|
| CIAPIN1 | Q6FI81 | PABPN1 | Q86U42 | HSPA8 | P11142 | PARK7 | Q99497 | RPL7A | P62424 | SREK1IP<br>1 | Q8N9Q2 |
| PELO | Q9BRX2 | FAHD1 | Q6P587 | HSPA9 | P38646 | PARP1 | P09874 | RPL8 | P62917 | SRP14 | P37108 |
| CSNK2B | P67870 | EIF6 | P56537 | HSPD1 | P10809 | PCBP1 | Q15365 | RPL9 | P32969 | SRP19 | P09132 |
| CNBP | P62633 | ENO1 | P06733 | HSPE1 | P61604 | PCBP2 | Q15366 | RPLP1 | P05386 | SRP68 | Q9UHB9 |
| CNN3 | Q15417 | SOD1 | P00441 | HSPH1 | Q92598 | PCCA | P05165 | RPLP2 | P05387 | SRP9 | P49458 |
| SUGP2 | Q8IX01 | VIM | P08670 | TPR | P12270 | PCMT1 | P22061 | RPS11 | P62280 | SRRM2 | Q9UQ35 |
| MRPS22 | P82650 | ERH | P84090 | PLRG1 | O43660 | PDAP1 | Q13442 | RPS12 | P25398 | SRSF1 | Q07955 |
| CPSF6 | Q16630 | EWSR1 | Q01844 | IGF2BP1 | Q9NZI8 | PHF6 | Q8IWS0 | RPS13 | P62277 | SRSF11 | Q05519 |
| MRPS26 | Q9BYN8 | EXOSC6 | Q5RKV6 | IGF2BP3 | O00425 | PHGDH | O43175 | RPS14 | P62263 | SRSF3 | P84103 |
| TUBB | P07437 | FASN | P49327 | ILF3 | Q12906 | PIN4 | Q9Y237 | RPS15 | P62841 | SRSF4 | Q08170 |
| TUBB4B | P68371 | FAU | P62861 | DNAJC8 | O75937 | PLK1 | P53350 | RPS15A | P62244 | SRSF5 | Q13243 |
| TXN | P10599 | FBL | P22087 | TMA16 | Q96EY4 | PLS3 | P13797 | RPS16 | P62249 | SRSF6 | Q13247 |
| U2AF2 | P26368 | RRP8 | O43159 | RBBP4 | Q09028 | POLR1C | O15160 | RPS17 | P08708 | SRSF7 | Q16629 |
| U2SURP | O15042 | FMR1 | Q06787-7 | POLR2E | P19388 | PPIA | P62937 | RPS18 | P62269 | SRSF9 | Q13242 |
| UBA52 | P62987 | FN3KRP | Q9HA64 | KHSRP | Q92945 | PPIB | P23284 | RPS19 | P39019 | SSB | P05455 |
| UPF1 | Q92900 | FRG1 | Q14331 | PWP1 | Q13610 | PPIL1 | Q9Y3C6 | RPS2 | P15880 | SUB1 | P53999 |
| VCP | P55072 | FUBP1 | Q96AE4 | KPNA2 | P52292 | PPIL4 | Q8WUA<br>2 | RPS20 | P60866 | TAGLN2 | P37802 |
| XRCC5 | P13010 | FUBP3 | Q96I24 | KPNB1 | Q14974 | PRDX1 | Q06830 | RPS21 | P63220 | TCOF1 | Q13428 |
| XRN2 | Q9H0D6 | G3BP1 | Q13283 | LARP1 | Q6PKG0 | PRPF4B | Q13523 | RPS23 | P62266 | TCP1 | P17987 |
| YBX1 | P67809 | G3BP2 | Q9UN86-2 | LMNB1 | P20700 | PTBP1 | P26599 | RPS24 | P62847 | THRAP3 | Q9Y2W<br>1 |
| YBX3 | P16989-2 | GAPDH | P04406 | LRRC47 | Q8N1G4 | RAB11FIP1 | Q6WKZ<br>4 | RPS25 | P62851 | TRA2B | P62995 |
| YWHAB | P31946-2 | GNB1L | Q9BYB4 | LRRC59 | Q96AG4 | RAN | P62826 | RPS26 | P62854 | TRIM28 | Q13263 |
| YWHAE | P62258 | ZC3HAV1 | Q7Z2W4 | ZFR | Q96KR1 | ZNF326 | Q5BKZ1 | ZNF706 | Q9Y5V0 | | |

### Table S5. Proteins displaying significant differential interactions

| Gene Name | Protein IDs | mutant_vs_control_log2 fold change | mutant_vs_control_p.val | mutant_vs_control_significant | |
|---|---|---|---|---|---|
| ACIN1 | Q9UKV3 | 2.68 | 0.0014 | TRUE | Apoptotic chromatin condensation inducer in the nucleus |
| AKT1S1 | Q96B36 | 3.87 | 2.56E-05 | TRUE | Proline-rich AKT1 substrate 1 |
| CD2AP | Q9Y5K6 | 3.08 | 0.000245 | TRUE | CD2-associated protein |
| CKAP5 | Q14008 | 2.18 | 0.0013 | TRUE | Cytoskeleton-associated protein 5 |
| CORO1B | Q9BR76 | 4.76 | 0.000167 | TRUE | Coronin-1B |
| COX6B1 | P14854 | 5.5 | 3.51E-06 | TRUE | Cytochrome c oxidase subunit 6B1 |
| CSNK2B | P67870 | -2.82 | 0.000532 | TRUE | Casein kinase II subunit beta |
| CTNND1 | O60716 | 2.3 | 0.000923 | TRUE | Catenin delta-1 |
| DPYSL5 | Q9BPU6 | 3.8 | 0.000207 | TRUE | Dihydropyrimidinase-related protein 5 |
| ELAC2 | Q9BQ52 | 2.61 | 0.000164 | TRUE | Zinc phosphodiesterase ELAC protein 2 |
| GCN1L1 | Q92616 | 2.76 | 0.00025 | TRUE | Translational activator GCN1 |
| HN1 | Q9UK76 | 1.76 | 0.00123 | TRUE | Hematological and neurological expressed 1 protein |
| KRT18 | P05783 | 2.63 | 0.000766 | TRUE | Keratin, type I cytoskeletal 18 |
| MCMBP | Q9BTE3 | 1.88 | 0.00116 | TRUE | Mini-chromosome maintenance complex-binding protein |
| MRPL40 | Q9NQ50 | 2.92 | 0.0016 | TRUE | 39S ribosomal protein L40, mitochondrial |
| MRPS36 | P82909 | 4.31 | 0.00034 | TRUE | 28S ribosomal protein S36, mitochondrial |
| MSH6 | P52701 | 3.63 | 0.000413 | TRUE | DNA mismatch repair protein Msh6 |
| NUP153 | P49790 | 2.27 | 0.000366 | TRUE | Nuclear pore complex protein Nup153 |
| PALLD | Q8WX93 | 2.94 | 0.000289 | TRUE | Palladin |
| PAWR | Q96IZ0 | 2.58 | 0.00031 | TRUE | PRKC apoptosis WT1 regulator protein |
| PIN1 | Q13526 | 3.71 | 0.00102 | TRUE | Peptidyl-prolyl cis-trans isomerase NIMA-interacting 1 |

| | | | | | |
|---|---|---|---|---|---|
| PNPO | Q9NVS9 | 3.13 | 0.00111 | TRUE | Pyridoxine-5-phosphate oxidase |
| PPP1R14B | Q96C90 | 3.61 | 0.000247 | TRUE | Protein phosphatase 1 regulatory subunit 14B |
| PPP1R14C | Q8TAE6 | 2.74 | 0.000809 | TRUE | Protein phosphatase 1 regulatory subunit 14C |
| PRPF19 | Q9UMS4 | -1.89 | 0.000803 | TRUE | Pre-mRNA-processing factor 19 |
| PRRC2C | E7EPN9 | 1.76 | 0.00119 | TRUE | Protein PRRC2C |
| PTMS | P20962 | 4.09 | 0.000322 | TRUE | Parathymosin |
| RANGAP1 | P46060 | 2.39 | 0.00113 | TRUE | Ran GTPase-activating protein 1 |
| RBM10 | P98175 | 2.41 | 0.00211 | TRUE | RNA-binding protein 10 |
| STMN1 | P16949 | 2.03 | 0.000652 | TRUE | Stathmin |
| TMA16 | Q96EY4 | -3.23 | 0.000435 | TRUE | Translation machinery-associated protein 16 |
| TOP1 | P11387 | -3.46 | 0.000197 | TRUE | DNA topoisomerase 1 |
| TOR1AIP1 | Q5JTV8-3 | 2.46 | 0.000327 | TRUE | Torsin-1A-interacting protein 1 |
| YARS2 | Q9Y2Z4 | 3.17 | 0.000165 | TRUE | Tyrosine--tRNA ligase, mitochondrial;Tyrosine--tRNA ligase |
| ZC3H4 | Q9UPT8 | 3.64 | 0.000483 | TRUE | Zinc finger CCCH domain-containing protein 4 |
| NUP98 | P52948 | 1.49 | 0.00712 | TRUE | Nuclear pore complex protein Nup98-Nup96;Nuclear pore complex protein Nup98;Nuclear pore complex protein Nup96 |
| ATP6V1B2 | P21281 | 1.87 | 0.00453 | TRUE | V-type proton ATPase subunit B, brain isoform |
| ZRANB2 | O95218 | 2.97 | 0.00258 | TRUE | Zinc finger Ran-binding domain-containing protein 2 |
| VIM | P08670 | 1.7 | 0.00279 | TRUE | Vimentin |
| TXLNG | Q9NUQ3 | 2.69 | 0.00396 | TRUE | Gamma-taxilin |
| SYVN1 | Q86TM6 | 1.46 | 0.00976 | TRUE | E3 ubiquitin-protein ligase synoviolin |
| SNIP1 | Q8TAD8 | -2.66 | 0.00396 | TRUE | Smad nuclear-interacting protein 1 |
| OGFR | Q9NZT2 | 1.55 | 0.00719 | TRUE | Opioid growth factor receptor |
| MCM2 | P49736 | 1.52 | 0.00511 | TRUE | DNA replication licensing factor MCM2 |
| DDX23 | Q9BUQ8 | 1.49 | 0.00124 | TRUE | Probable ATP-dependent RNA helicase DDX23 |
| EPB41 | P11171 | 1.62 | 0.00538 | TRUE | Protein 4.1 |

| | | | | | |
|---|---|---|---|---|---|
| HNRNPA1 | P09651 | 1.59 | 0.00319 | TRUE | Heterogeneous nuclear ribonucleoprotein A1;Heterogeneous nuclear ribonucleoprotein A1, N-terminally processed |
| GLUL | P15104 | 2.8 | 0.00715 | TRUE | Glutamine synthetase |
| GNL1 | P36915 | 2.33 | 0.00459 | TRUE | Guanine nucleotide-binding protein-like 1 |

### Table S6. Phosphorylation

| | | | Intensity of phosphorylation | | | | | |
|---|---|---|---|---|---|---|---|---|
| Protein Name | phospho site | Localization prob | N mutant (Rep1) | N mutant (Rep2) | N mutant (Rep3) | N control (Rep1) | N control (Rep2) | N control (Rep3) |
| Nucleo-capsid (N) | **S206** | 0.994527 | 207590000 | 13224000 | 2966400 | 0 | 0 | 0 |
| Nucleo-capsid (N) | **S2** | 1 | 955740 | 1907100 | 2265000 | 1921900 | 2763700 | 3502800 |
| Nucleo-capsid (N) | **S79** | 0.969512 | 1327000 | 1562200 | 2342900 | 2651100 | 2688800 | 2985200 |
| Nucleo-capsid (N) | **S180** | 0.997001 | 28745000 | 41617000 | 50307000 | 35401000 | 43784000 | 47220000 |
| Nucleo-capsid (N) | **S176** | 0.9764 | 15311000 | 11139000 | 12863000 | 13966000 | 18277000 | 18382000 |

| Log2 of Intensities (related to Figure 4G) | | | | | | |
|---|---|---|---|---|---|---|
| | N-mutant | | | N-control | | |
| S206-ph | 27.6291617 | 23.6566553 | 21.5002817 | NA | NA | NA |
| S2-ph | 19.8662587 | 20.8629491 | 21.1110796 | 20.8741018 | 21.3981696 | 21.7400772 |
| S79-ph | 20.3397369 | 20.5751477 | 21.1598639 | 21.3381597 | 21.358531 | 21.5093962 |
| S180-ph | 24.7768077 | 25.3106696 | 25.5842558 | 25.0772868 | 25.3839004 | 25.4928947 |
| S176-ph | 23.8680652 | 23.4091164 | 23.6167238 | 23.7354155 | 24.1235259 | 24.1317904 |

### Table S7. Differentially expressed genes

| logFC (N-mutant) | logFC (N-control) | logCPM | LR | PValue | adj.P.Val | Symbols | Name |
|---|---|---|---|---|---|---|---|
| 6.5909 | 5.7375 | 5.548 | 569.36 | 2.32E-124 | 9.22E-121 | IFI44 | interferon induced protein 44 |
| 4.7401 | 3.9182 | 6.2177 | 499.18 | 4.01E-109 | 9.13E-106 | IFITM1 | interferon induced transmembrane protein 1 |
| 4.5185 | 3.8033 | 6.1309 | 710.47 | 5.28E-155 | 2.81E-151 | IFI44L | interferon induced protein 44 like |
| 4.4642 | 3.5178 | 6.8124 | 439.92 | 2.96E-96 | 5.25E-93 | IFIT3 | interferon induced protein with tetratricopeptide repeats 3 |
| 4.2358 | 3.4523 | 7.2185 | 847.95 | 7.42E-185 | 1.18E-180 | IFI6 | interferon alpha inducible protein 6 |
| 4.0804 | 3.3747 | 6.3073 | 756.23 | 6.11E-165 | 4.87E-161 | BST2 | bone marrow stromal cell antigen 2 |
| 3.9799 | 3.1183 | 4.7279 | 228.02 | 3.07E-50 | 2.33E-47 | RSAD2 | radical S-adenosyl methionine domain containing 2 |
| 3.9792 | 2.9881 | 3.5638 | 123.98 | 1.20E-27 | 6.81E-25 | OAS1 | 2'-5'-oligoadenylate synthetase 1 |
| 3.9121 | 3.1594 | 6.5215 | 347.03 | 4.41E-76 | 5.02E-73 | IFIT2 | interferon induced protein with tetratricopeptide repeats 2 |
| 3.8331 | 3.0151 | 8.0227 | 516.7 | 6.31E-113 | 1.68E-109 | IFIT1 | interferon induced protein with tetratricopeptide repeats 1 |
| 3.7219 | 2.9716 | 6.2972 | 431.3 | 2.21E-94 | 3.52E-91 | ISG15 | ISG15 ubiquitin like modifier |
| 3.1787 | 2.7725 | 1.3727 | 20.032 | 4.47E-05 | 0.0036888 | CXCL10 | C-X-C motif chemokine ligand 10 |
| 3.0411 | 2.3176 | 7.3672 | 408.78 | 1.71E-89 | 2.27E-86 | OAS3 | 2'-5'-oligoadenylate synthetase 3 |
| 2.7304 | 1.526 | 4.3908 | 36.542 | 1.16E-08 | 2.37E-06 | PARP10 | poly(ADP-ribose) polymerase family member 10 |
| 2.6618 | 2.2041 | 6.6264 | 549.98 | 3.74E-120 | 1.19E-116 | DDX60 | DExD/H-box helicase 60 |
| 2.5985 | -1.0584 | 0.74694 | 22.054 | 1.63E-05 | 0.0015893 | RPL37P6 | ribosomal protein L37 pseudogene 6 |
| 2.5804 | 1.7983 | 4.6689 | 221.45 | 8.19E-49 | 5.93E-46 | CMPK2 | cytidine/uridine monophosphate kinase 2 |
| 2.5427 | 1.9862 | 8.1562 | 484.13 | 7.45E-106 | 1.48E-102 | DDX58 | DExD/H-box helicase 58 |
| 2.5298 | 2.5444 | 2.3412 | 24.707 | 4.31E-06 | 0.00051599 | HSD17B13 | hydroxysteroid 17-beta dehydrogenase 13 |
| 2.4544 | 1.7193 | 6.6465 | 243.69 | 1.21E-53 | 1.01E-50 | IFITM3 | interferon induced transmembrane protein 3 |
| 2.3399 | 1.917 | 1.698 | 17.42 | 0.00016493 | 0.011042 | SP140 | SP140 nuclear body protein |
| 2.3349 | 1.7178 | 6.5483 | 235 | 9.34E-52 | 7.44E-49 | HELZ2 | helicase with zinc finger 2 |
| 2.2868 | 1.9946 | 4.0167 | 82.703 | 1.10E-18 | 4.74E-16 | SAMD9L | sterile alpha motif domain containing 9 like |
| 2.2536 | 1.4984 | 3.3321 | 65.422 | 6.22E-15 | 2.11E-12 | EPSTI1 | epithelial stromal interaction 1 |
| 2.2083 | 1.727 | 4.2706 | 121.15 | 4.93E-27 | 2.62E-24 | OASL | 2'-5'-oligoadenylate synthetase like |

| | | | | | | | |
|---|---|---|---|---|---|---|---|
| 2.1724 | 1.6504 | 4.8791 | 188.59 | 1.12E-41 | 7.41E-39 | IFIH1 | interferon induced with helicase C domain 1 |
| 2.1156 | 1.6829 | 6.6982 | 376.9 | 1.44E-82 | 1.76E-79 | STAT1 | signal transducer and activator of transcription 1 |
| 2.04 | 1.581 | 8.7036 | 427.94 | 1.19E-93 | 1.72E-90 | DTX3L | deltex E3 ubiquitin ligase 3L |
| 2.0071 | 1.6722 | 5.909 | 298.29 | 1.69E-65 | 1.68E-62 | PLSCR1 | phospholipid scramblase 1 |
| 2.0001 | 1.9709 | 4.1073 | 47.297 | 5.37E-11 | 1.45E-08 | ZNF625-ZNF20 | ZNF625-ZNF20 readthrough (NMD candidate) |
| 1.905 | 1.6937 | 3.5908 | 65.531 | 5.89E-15 | 2.04E-12 | IRF9 | interferon regulatory factor 9 |
| 1.9005 | 1.6755 | 2.7615 | 22.49 | 1.31E-05 | 0.0013267 | GBP1 | guanylate binding protein 1 |
| 1.8816 | 1.2956 | 9.0956 | 296.13 | 4.96E-65 | 4.65E-62 | IFIT5 | interferon induced protein with tetratricopeptide repeats 5 |
| 1.8438 | 2.6727 | 2.1521 | 23.024 | 1.00E-05 | 0.0010654 | GRIP2 | glutamate receptor interacting protein 2 |
| 1.8249 | 1.3301 | 1.9432 | 14.442 | 0.00073102 | 0.036745 | PLSCR2 | phospholipid scramblase 2 |
| 1.8017 | 1.4185 | 7.6401 | 317.26 | 1.28E-69 | 1.36E-66 | SAMD9 | sterile alpha motif domain containing 9 |
| 1.7724 | 1.3604 | 5.3408 | 187.27 | 2.16E-41 | 1.38E-38 | HERC6 | HECT and RLD domain containing E3 ubiquitin protein ligase family member 6 |
| 1.7705 | 1.3244 | 4.4859 | 71.418 | 3.10E-16 | 1.24E-13 | UBE2L6 | ubiquitin conjugating enzyme E2 L6 |
| 1.7036 | 1.3829 | 6.7467 | 265.36 | 2.38E-58 | 2.11E-55 | PARP9 | poly(ADP-ribose) polymerase family member 9 |
| 1.6596 | 1.0636 | 4.7397 | 119.26 | 1.27E-26 | 6.53E-24 | TRIM22 | tripartite motif containing 22 |
| 1.6537 | 1.2988 | 4.069 | 58.028 | 2.51E-13 | 7.99E-11 | SP110 | SP110 nuclear body protein |
| 1.6507 | -0.1502 | 2.5445 | 37.187 | 8.41E-09 | 1.86E-06 | MYLK4 | myosin light chain kinase family member 4 |
| 1.6461 | 1.1795 | 3.2144 | 35.346 | 2.11E-08 | 4.26E-06 | UBA7 | ubiquitin like modifier activating enzyme 7 |
| 1.6313 | 1.5866 | 4.433 | 89.156 | 4.37E-20 | 2.11E-17 | ARL14EPP1 | ARL14EP pseudogene 1 |
| 1.6292 | 1.2168 | 4.7705 | 106.56 | 7.26E-24 | 3.61E-21 | PARP12 | poly(ADP-ribose) polymerase family member 12 |
| 1.5802 | 1.6322 | 2.9138 | 26.263 | 1.98E-06 | 0.00026104 | RAB1AP1 | RAB1A pseudogene 1 |
| 1.5745 | 1.12 | 3.1564 | 29.581 | 3.77E-07 | 6.07E-05 | IFI27 | interferon alpha inducible protein 27 |
| 1.5516 | 1.6406 | 2.8207 | 27.474 | 1.08E-06 | 0.00015811 | CYP2J2 | cytochrome P450 family 2 subfamily J member 2 |
| 1.5515 | 0.97201 | 3.5967 | 32.437 | 9.05E-08 | 1.70E-05 | SYT5 | synaptotagmin 5 |
| 1.5146 | 1.5784 | 2.4415 | 19.426 | 6.05E-05 | 0.0048674 | REC8 | REC8 meiotic recombination protein |
| 1.4569 | 1.0543 | 7.1461 | 146.05 | 1.93E-32 | 1.14E-29 | USP18 | ubiquitin specific peptidase 18 |
| 1.4401 | 2.2006 | 5.1219 | 27.256 | 1.21E-06 | 0.00016817 | ELFN2 | extracellular leucine rich repeat and fibronectin type III |

| | | | | | | | |
|---|---|---|---|---|---|---|---|
| | | | | | | | domain containing 2 |
| 1.3902 | 0.82668 | 3.3307 | 18.779 | 8.36E-05 | 0.0064027 | ACE2 | angiotensin I converting enzyme 2 |
| 1.36 | 1.295 | 5.3129 | 44.627 | 2.04E-10 | 5.24E-08 | CXCL8 | C-X-C motif chemokine ligand 8 |
| 1.3078 | 1.1937 | 6.9007 | 37.632 | 6.74E-09 | 1.51E-06 | H2AC6 | H2A clustered histone 6 |
| 1.2603 | 0.48038 | 3.8104 | 31.406 | 1.51E-07 | 2.74E-05 | IFI16 | interferon gamma inducible protein 16 |
| 1.2502 | 0.87272 | 8.3571 | 174.23 | 1.47E-38 | 8.98E-36 | PARP14 | poly(ADP-ribose) polymerase family member 14 |
| 1.244 | 1.1707 | 3.3145 | 14.832 | 0.00060168 | 0.032064 | OR51E2 | olfactory receptor family 51 subfamily E member 2 |
| 1.1763 | 0.97392 | 8.696 | 200.82 | 2.47E-44 | 1.71E-41 | EIF2AK2 | eukaryotic translation initiation factor 2 alpha kinase 2 |
| 1.1332 | -0.45046 | 3.552 | 34.224 | 3.70E-08 | 7.28E-06 | FBXL15 | F-box and leucine rich repeat protein 15 |
| 1.0949 | 0.79509 | 4.6993 | 22.026 | 1.65E-05 | 0.0015981 | SLC6A6 | solute carrier family 6 member 6 |
| 1.0592 | 0.68795 | 4.8391 | 60.561 | 7.07E-14 | 2.35E-11 | DDX60L | DExD/H-box 60 like |
| 1.0586 | -3.9904 | 4.6686 | 66.5 | 3.63E-15 | 1.34E-12 | FKBPL | FKBP prolyl isomerase like |
| 1.0167 | 0.61158 | 3.7267 | 14.183 | 0.00083222 | 0.040552 | IFI35 | interferon induced protein 35 |
| 0.8698 | 1.1839 | 3.6252 | 25.709 | 2.61E-06 | 0.00033872 | KREMEN2 | kringle containing transmembrane protein 2 |
| 0.85614 | 1.3047 | 3.7445 | 37.045 | 9.03E-09 | 1.97E-06 | INHBA | inhibin subunit beta A |
| 0.60454 | -1.2691 | 1.9918 | 14.734 | 0.0006317 | 0.03311 | RSPH10B2 | radial spoke head 10 homolog B2 |
| 0.40296 | 1.4471 | 2.4986 | 21.083 | 2.64E-05 | 0.002351 | FPR3 | formyl peptide receptor 3 |
| 0.32437 | 1.6498 | 1.6979 | 14.74 | 0.00062979 | 0.03311 | PDE2A | phosphodiesterase 2A |
| 0.2971 | -1.3841 | 4.5746 | 24.477 | 4.84E-06 | 0.00056699 | CXCR5 | C-X-C motif chemokine receptor 5 |
| 0.0016387 | -1.035 | 3.9289 | 28.453 | 6.63E-07 | 9.96E-05 | RIPPLY3 | ripply transcriptional repressor 3 |
| -0.151 | -1.6464 | 2.5644 | 23.457 | 8.06E-06 | 0.00088573 | TMPRSS13 | transmembrane serine protease 13 |
| -0.29246 | -1.5043 | 2.9939 | 27.77 | 9.33E-07 | 0.00013766 | HSPA12B | heat shock protein family A (Hsp70) member 12B |
| -0.39018 | -3.2416 | 0.8389 | 14.735 | 0.00063135 | 0.03311 | OR2W6P | olfactory receptor family 2 subfamily W member 6 pseudogene |
| -0.9212 | 1.0481 | 2.4626 | 19.105 | 7.10E-05 | 0.005575 | LHX6 | LIM homeobox 6 |
| -1.1054 | -0.18711 | 3.2197 | 16.383 | 0.00027703 | 0.017176 | CLDN2 | claudin 2 |
| -1.1267 | -1.4016 | 9.0029 | 27.408 | 1.12E-06 | 0.00015905 | ANKRD30BL | ankyrin repeat domain 30B like |
| -1.1349 | 0.20722 | 2.518 | 14.474 | 0.00071936 | 0.036273 | LMO1 | LIM domain only 1 |
| -1.1446 | -0.94771 | 3.9669 | 15.525 | 0.00042541 | 0.024037 | SRGAP2D | SLIT-ROBO Rho GTPase activating protein 2D |

| | | | | | | | |
|---|---|---|---|---|---|---|---|
| | | | | | | | (pseudogene) |
| -1.1891 | -0.0015398 | 4.5206 | 26.38 | 1.87E-06 | 0.0002503 | SPECC1L-ADORA2A | SPECC1L-ADORA2A readthrough (NMD candidate) |
| -1.2704 | -1.8716 | 4.1256 | 23.855 | 6.61E-06 | 0.00073607 | FMC1-LUC7L2 | FMC1-LUC7L2 readthrough |
| -1.2848 | -0.88484 | 6.7341 | 88.673 | 5.56E-20 | 2.60E-17 | H3P6 | H3 histone pseudogene 6 |
| -1.3599 | -2.2103 | 1.006 | 13.752 | 0.0010321 | 0.047947 | ADAD2 | adenosine deaminase domain containing 2 |
| -1.3833 | 0.077091 | 2.9318 | 27.335 | 1.16E-06 | 0.00016354 | APOBEC3D | apolipoprotein B mRNA editing enzyme catalytic subunit 3D |
| -1.4217 | -1.0934 | 2.3448 | 18.119 | 0.0001163 | 0.0085002 | CYP26C1 | cytochrome P450 family 26 subfamily C member 1 |
| -1.5358 | -0.17689 | 3.15 | 24.833 | 4.05E-06 | 0.00048905 | TMEM272 | transmembrane protein 272 |
| -1.6351 | 1.1156 | 1.6829 | 16.453 | 0.00026747 | 0.016648 | IRX4 | iroquois homeobox 4 |
| -1.6583 | 0.058962 | 1.7474 | 14.75 | 0.00062658 | 0.03311 | CST7 | cystatin F |
| -1.7152 | 0.27984 | 3.449 | 29.492 | 3.94E-07 | 6.28E-05 | PCDHGC5 | "protocadherin gamma subfamily C, 5" |
| -1.7881 | -1.9206 | 1.4148 | 17.614 | 0.0001497 | 0.01034 | SERBP1P5 | SERPINE1 mRNA binding protein 1 pseudogene 5 |
| -2.0378 | -1.1984 | 3.0617 | 20.799 | 3.04E-05 | 0.0026221 | RPS28P7 | ribosomal protein S28 pseudogene 7 |
| -2.0497 | 0.1407 | 2.3942 | 25.451 | 2.97E-06 | 0.0003732 | MYRIP | myosin VIIA and Rab interacting protein |
| -2.1033 | -0.17562 | 3.7323 | 14.523 | 0.0007022 | 0.035633 | CCBE1 | collagen and calcium binding EGF domains 1 |
| -2.1153 | -0.8227 | 1.6676 | 17.81 | 0.00013569 | 0.0097389 | ANO4 | anoctamin 4 |
| -2.2277 | -1.4625 | 4.3468 | 27.063 | 1.33E-06 | 0.0001825 | RPL36A-HNRNPH2 | RPL36A-HNRNPH2 readthrough |
| -2.2726 | -1.7089 | 1.6856 | 16.46 | 0.00026653 | 0.016648 | EIF3FP3 | eukaryotic translation initiation factor 3 subunit F pseudogene 3 |
| -2.2829 | -1.1793 | 1.7004 | 21.808 | 1.84E-05 | 0.001733 | CLDN23 | claudin 23 |
| -3.8726 | -1.8859 | 1.7013 | 35.035 | 2.47E-08 | 4.92E-06 | FRG2C | FSHD region gene 2 family member C |

### Table S8. Primers

**Primers used for cloning**

| **Primer Name** | **Sequence 5' to 3'** |
|---|---|
| pLVX-N1-F1 | CTATTTCCGGTGAATTCGCCG |
| pLVX-N1-R1 | GGGGCGGGATCCTTACTTTTC |
| pLVX-N1-Mut-F1 | CCAGGGTCCAGTAAACGAACAAGTCCGGCGC |
| pLVX-N1-Mut-R1 | GCGCCGGACTTGTTCGTTTACTGGACCCTGG |

| **2019-nCoV CDC Primers and Probe** | |
|---|---|
| **Name** | **Catalog#** |
| nCOV_N1 Forward Primer Aliquot, 50 nmol | 10006821 |
| nCOV_N1 Reverse Primer Aliquot, 50 nmol | 10006822 |
| nCOV_N1 Probe Aliquot, 25 nmol | 10006823 |
| nCOV_N2 Forward Primer Aliquot, 50 nmol | 10006824 |
| nCOV_N2 Reverse Primer Aliquot, 50 nmol | 10006825 |
| nCOV_N2 Probe Aliquot, 25 nmol | 10006826 |
| E gene E_Sarbeco_F | ACAGGTACGTTAATAGTTAATAGCGT |
| E_Sarbeco_R | ATATTGCAGCAGTACGCACACA |
| E_Sarbeco_P1 | FAM-ACACTAGCCATCCTTACTGCGCTTCG-BBQ |
