## Supplementary Figures S1-S13 for "Saudi Arabian SARS-CoV-2 genomes implicate a mutant Nucleocapsid protein in modulating host interactions and increased viral load in COVID-19 patients"

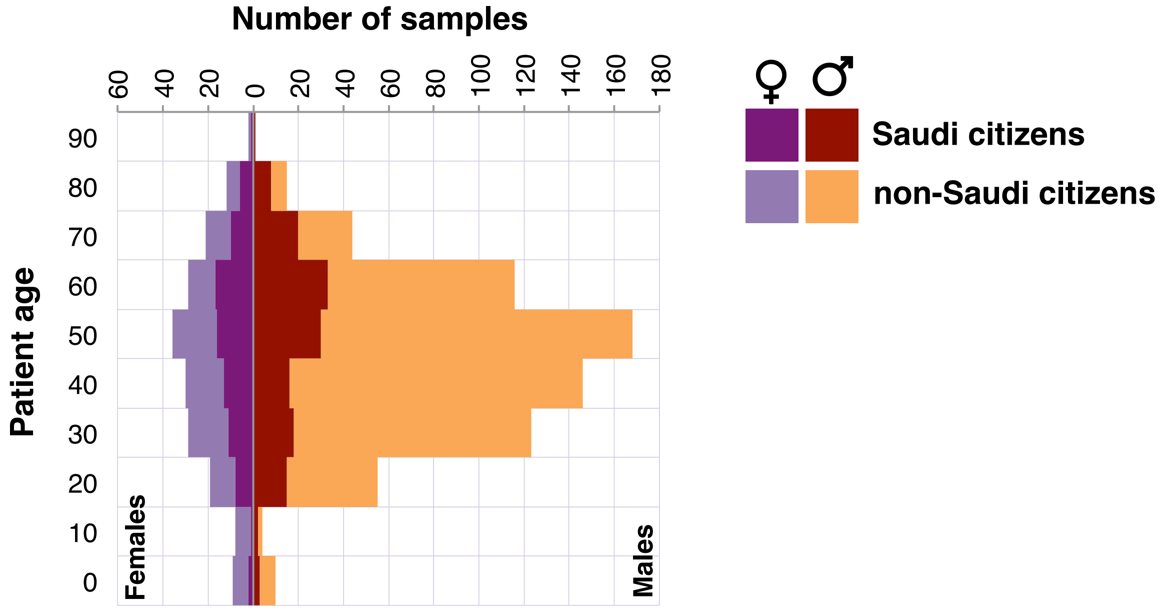


**Figure S1**

Patient age and gender shown for Saudi and non-Saudi citizens.

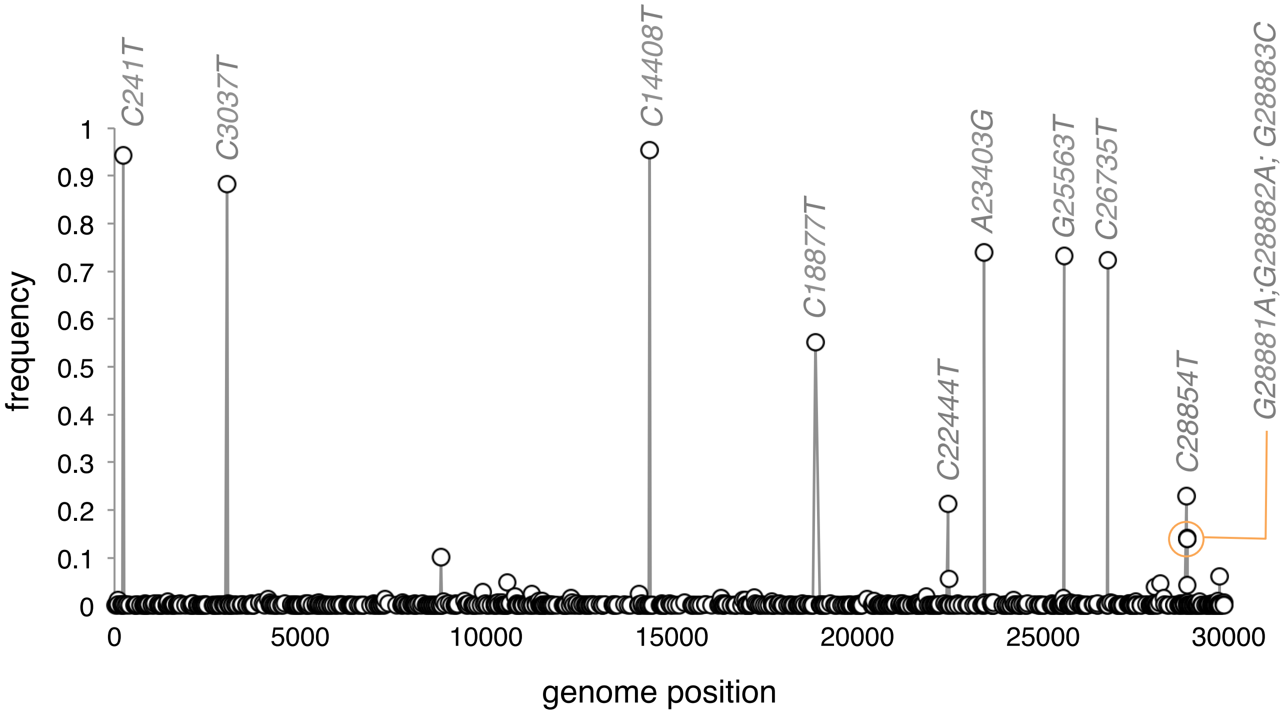


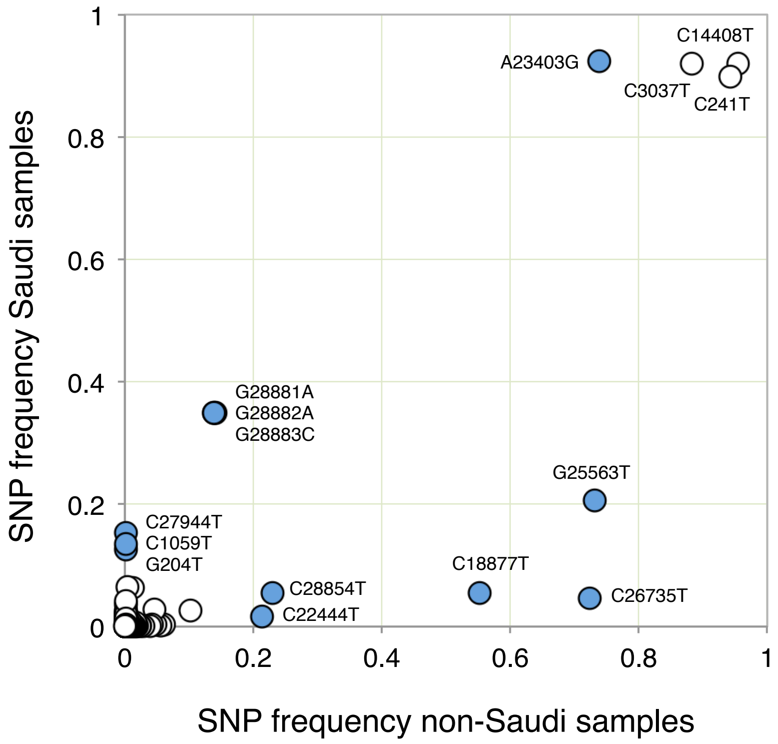


**Figure S2**
Top: The 836 detected SNPs are shown along their positions in the SARS-CoV-2 genome (x-axis) and their frequency in the Saudi samples (y-axis).
High-frequency SNPs are highlighted along with the 3 SNPs underlying the R203K/G204R changes in the N protein (G28881A;G2882A;G28883C).
Bottom: Scatter plot of SNP frequencies in Saudi samples (y-axis) and in global, non-Saudi samples available from GISAID in 2020. SNPs differing by at least 0.1 in absolute values are highlighted in blue.

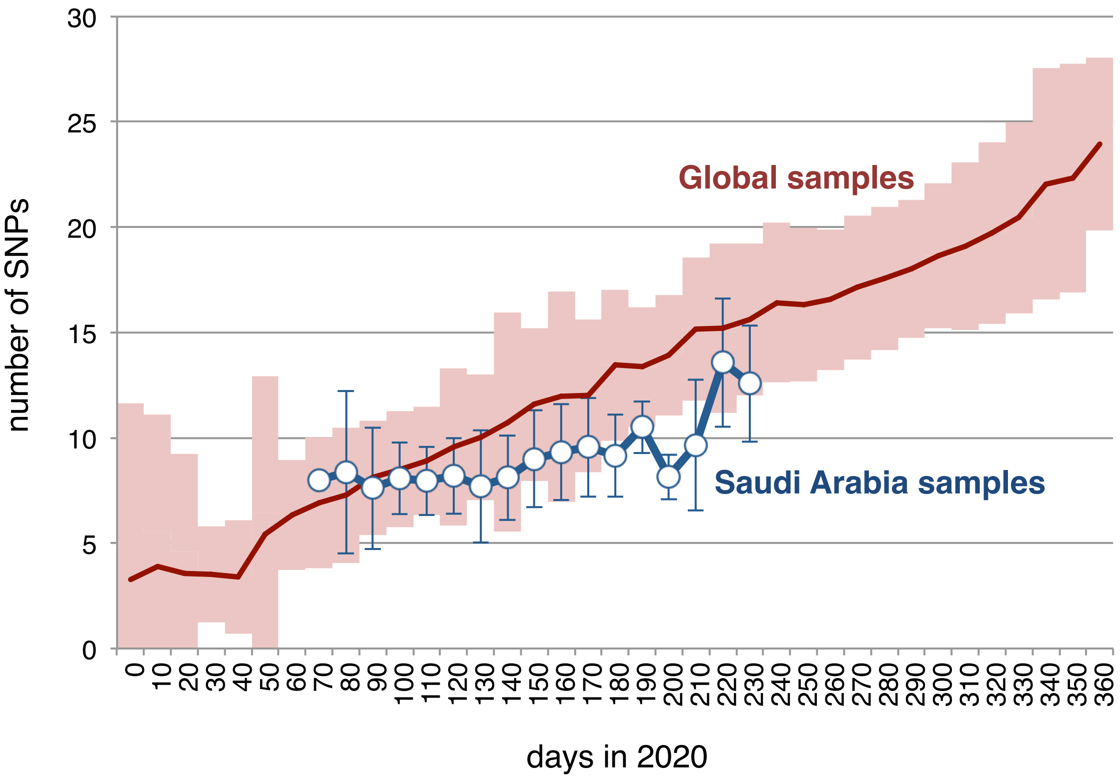


**Figure S3**
Samples were grouped into 10-days periods according to their sampling date. Within each time group, the average number of SNPs (compared to the Wuhan reference) was calculated and shown along with plus/minus one standard deviation.
Global non-Saudi Arabian samples are shown as red line, and Saudi Arabian samples from this study shown as blue line.

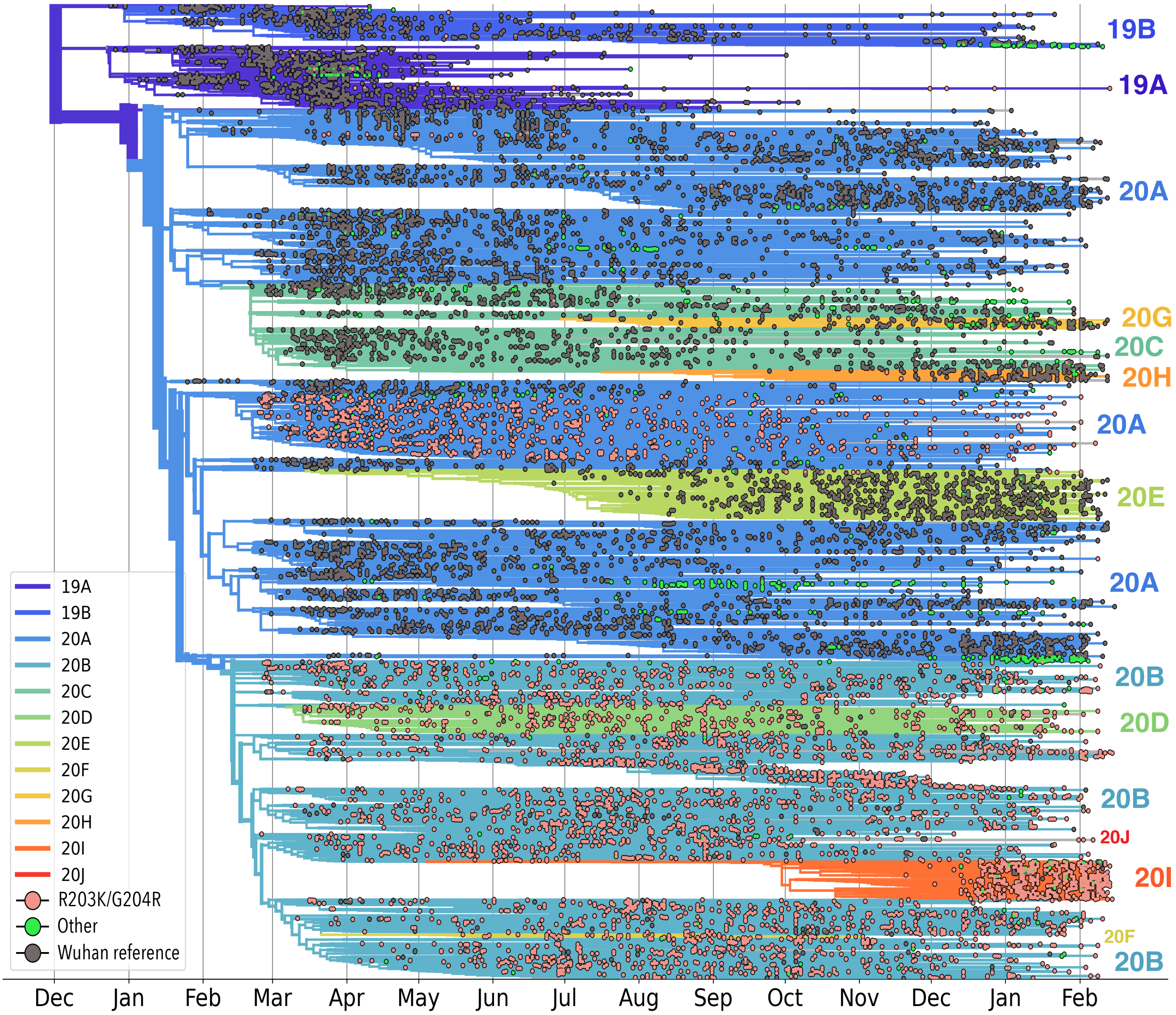


**Figure S4**

ML tree of 16,386 sequences dated with TreeTime showing the distribution of genotypes at genome positions 28,881-28,883 Samples are coloured according to their genotype and branches according to Nextstrain clades. R203K/G204R SNPs were identified from 590K samples submitted to GISAID on February 24, 2021, and are all included in the shown subset.

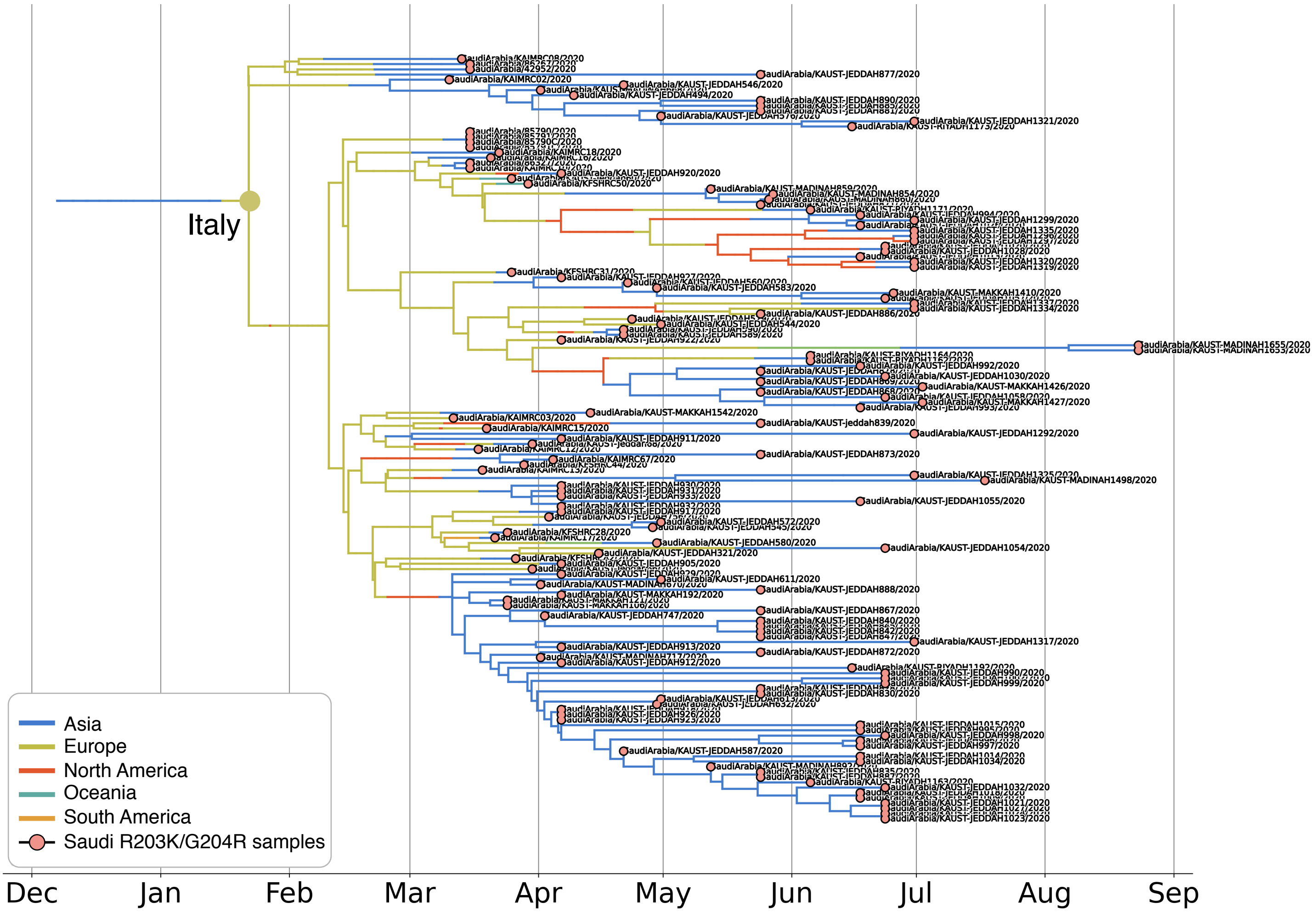


**Figure S5**
A ML tree was constructed for 25,198 sequences subsampled from 590K global sequences available on GISAID on 24 Feb 2021. Samples with closer genetic distance to Saudi samples were preferred. The predicted internal node-dates and possible country for internal nodes were inferred using TreeTime (see Methods). The resulting global phylogenetic tree was reduced to retain branches that lead to Saudi leaf nodes containing the R203K/G204R SNPs.

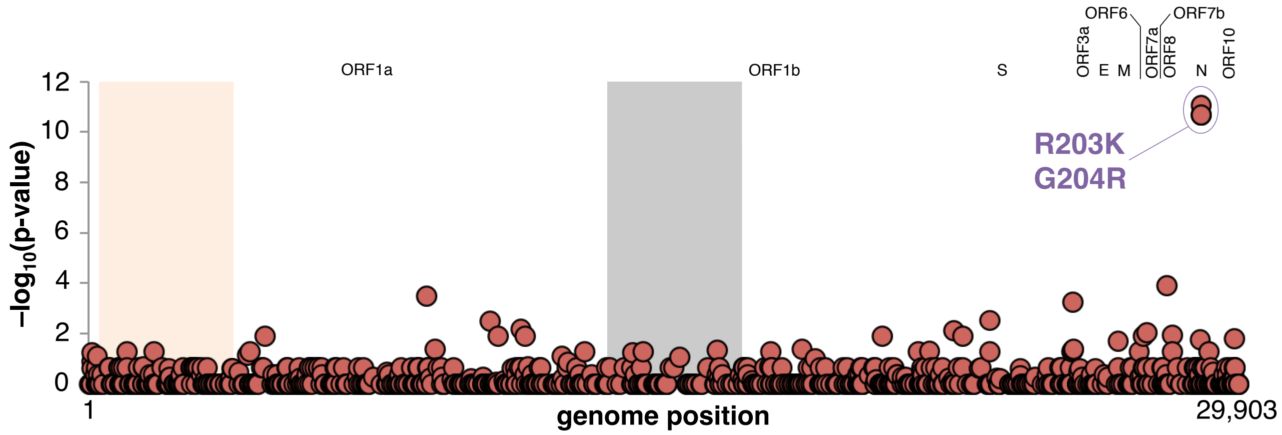


**Figure S6**

Manhattan plot showing the association between SARS-CoV-2 SNPs and recorded mortality in Saudi samples. Negative $\log_{10}$(uncorrected p-values) from Fisher's exact tests are shown as red circles. Gene boundaries are indicated by background colors (listed on top), and the three R203K/G204R SNPs (positions 28,881-28,883) in the N gene are highlighted.

| SNP | Count |
|---|---|
| **GGC** | 23 |
| **GAG** | 47 |
| **GAA** | 29 |
| **AGG** | 563 |
| **AGC** | 27 |
| **AAG** | 26 |
| **AAC** | 232,475 |

| Clade | R203K/G204R counts |
|---|---|
| 19A | 22 |
| 19B | 4 |
| 20A | 523 |
| 20A.EU2 | 30 |
| 20B | 114,487 |
| 20C | 12 |
| 20D | 5,671 |
| 20E (EU1) | 15 |
| 20F | 12,593 |
| 20G | 2 |
| 20H/501Y.V2 | 5 |
| 20I/501Y.V1 | 98,552 |
| 20J/501Y.V3 | 245 |

**Figure S7**
Global samples from GISAID (February 24th 2021).
Left: Counts of partial and full R203K/G204R SNPs at genome position 28,881-28,883, where the Wuhan reference has the GGG genotype.
Right: Counts of R203K/G204R SNPs in different Nextstrain clades.

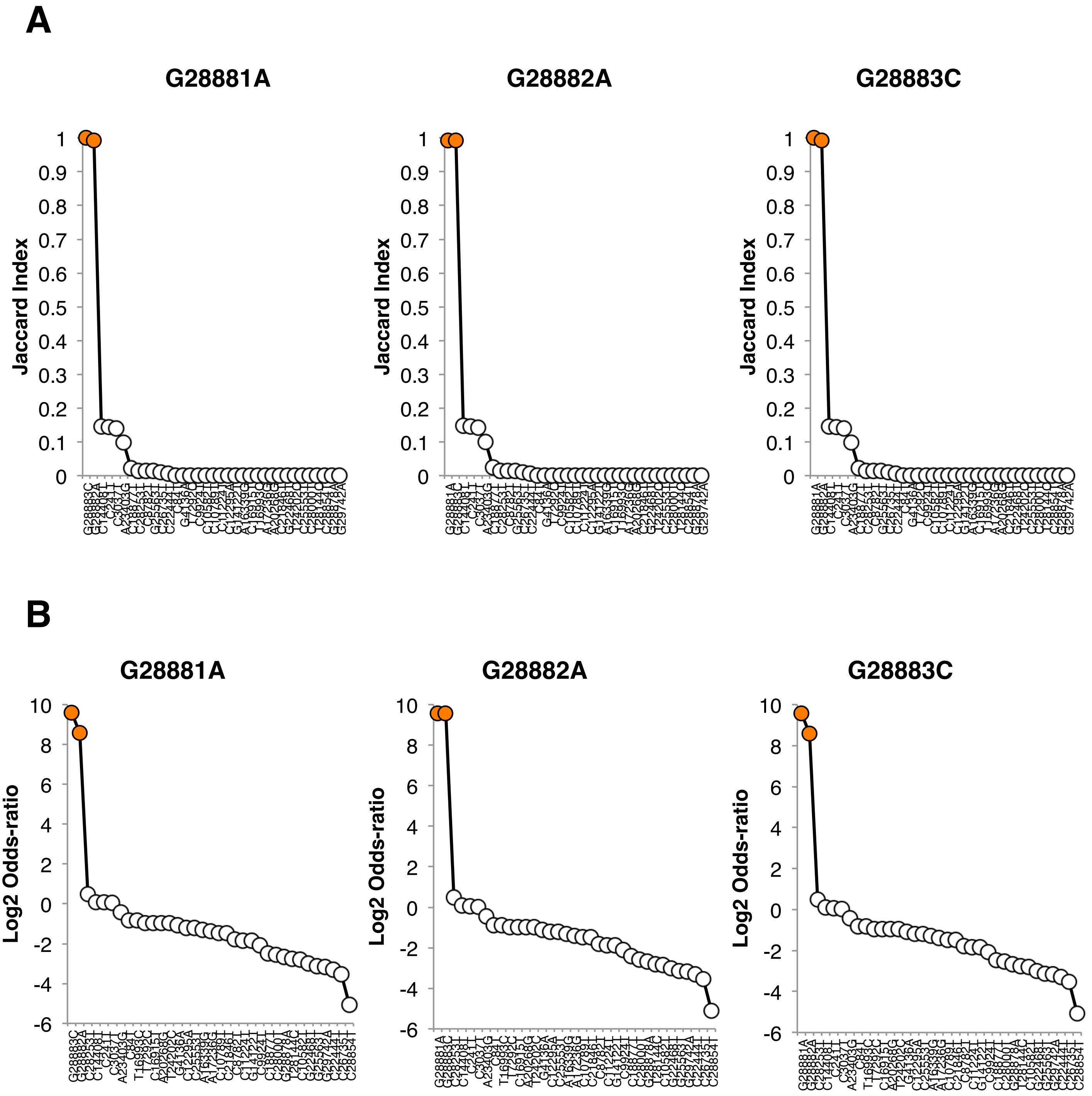


**Figure S8**
Co-occurrences of SNPs in 892 SARS-CoV-2 genomes from Saudi Arabia shown as Jaccard Index (A) and log2 odds-ratio (B).
The co-occurrence between the three SNPs in the R203K and G204R mutations (genomic mutations shown above plots) and all SNPs present in at least 20 samples (x-axes) are shown as circles. Co-occurrences between the three SNPs are highlighted in orange.

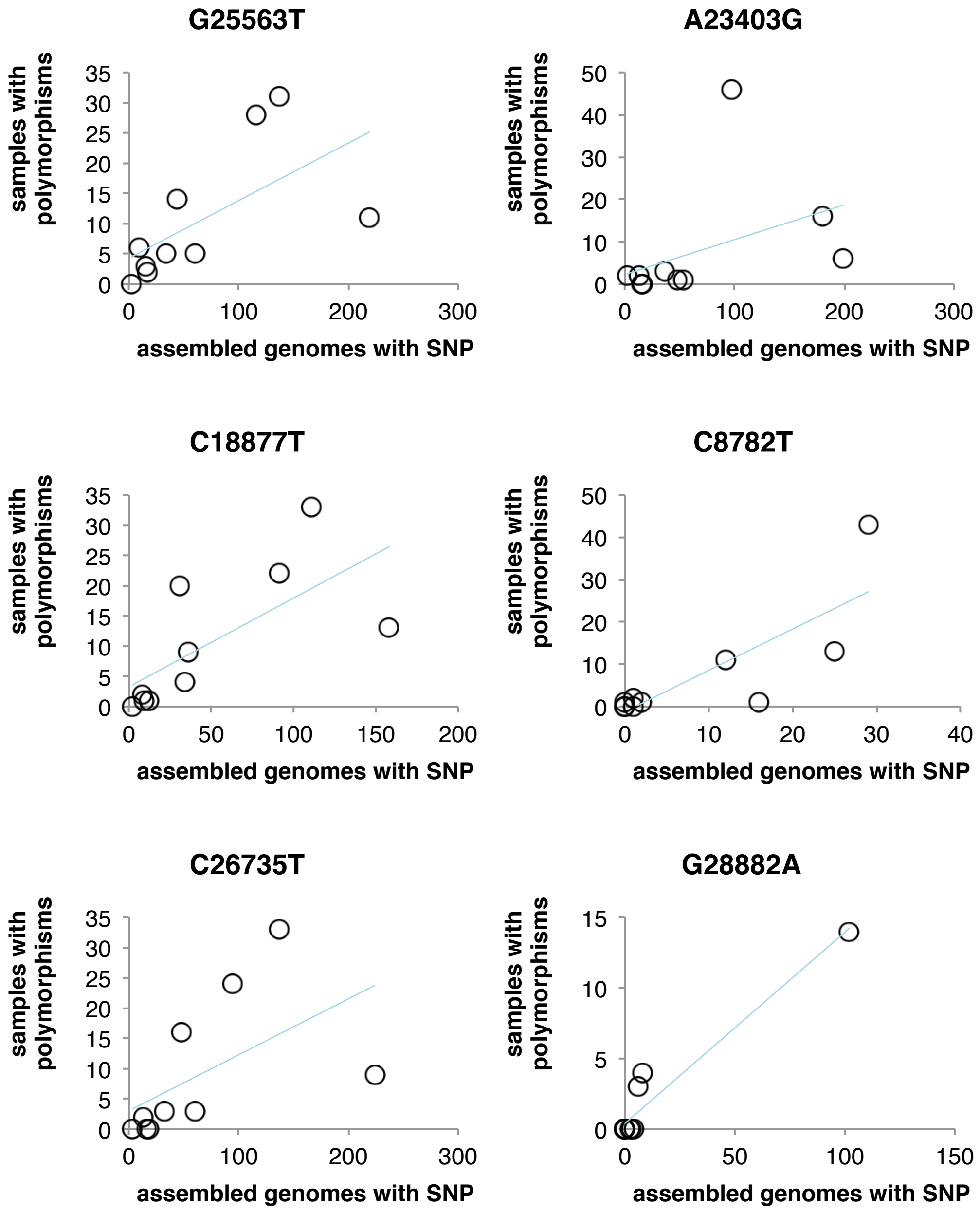


| | Kendall's tau | p-value |
|---|---|---|
| C173T | NA | NA |
| C241T | 0.644 | 0.0183 |
| C3037T | 0.564 | 0.03148 |
| C8782T | 0.700 | 0.008597 |
| G12833T | NA | NA |
| G14122T | 0.722 | 0.01192 |
| C14408T | 0.754 | 0.003397 |
| C17690T | 0.280 | 0.3732 |
| C18877T | 0.674 | 0.007058 |
| A23403G | 0.368 | 0.1475 |
| G25563T | 0.584 | 0.01955 |
| T26448C | NA | NA |
| C26735T | 0.675 | 0.008202 |
| T28144C | 0.658 | 0.01663 |
| C28311T | NA | NA |
| C28854T | 0.644 | 0.01311 |
| G28881A | 0.877 | 0.001608 |
| G28882A | 0.784 | 0.005768 |
| G28883C | 0.784 | 0.005768 |
| G29742A | 0.580 | 0.03878 |

**Figure S9**

For the 20 SNPs showing the highest levels of within-host polymorphisms in SARS-CoV-2 genomes from Saudi Arabia (see Supplementary Information), the number of samples with polymorphisms of this SNP (y-axes) and the number of samples with that SNP in the assembled reference genomes (x-axes) was plotted for each hospital. Each circle therefore represents a hospital. The correlation between these two parameters were then calculated (table, right). Four SNPs had polymorphisms but were not present in any assembled genome, and correlation could not be calculated ('NA' in table).

Plots are shown for the five SNPs with the highest instances of polymorphisms as well as one of the SNPs (G28882A) from the R203K/G204R mutations.

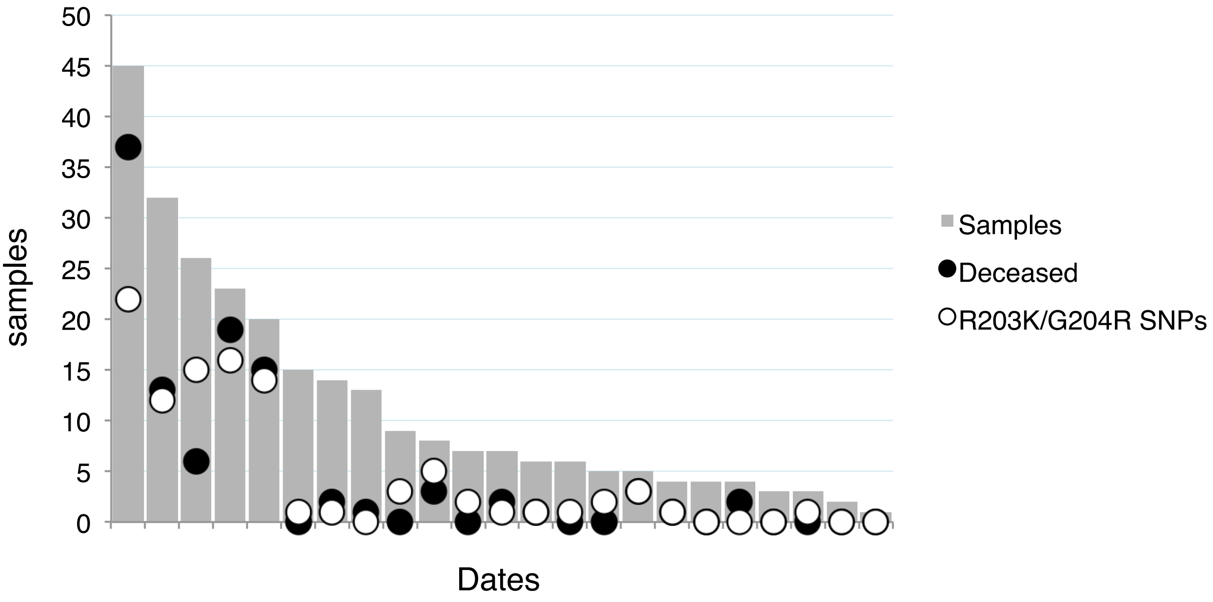


**Figure S10**
Bars show the number of samples from King Abdullah Medical Centre in Jeddah (y-axis) collected on different dates (x-axis). The number of samples with R203K/G204R SNPs are indicated by white circles, and the number of samples from deceased patients as black circles.
Dates are sorted according to their number of samples and are not labeled to ensure anonymity.

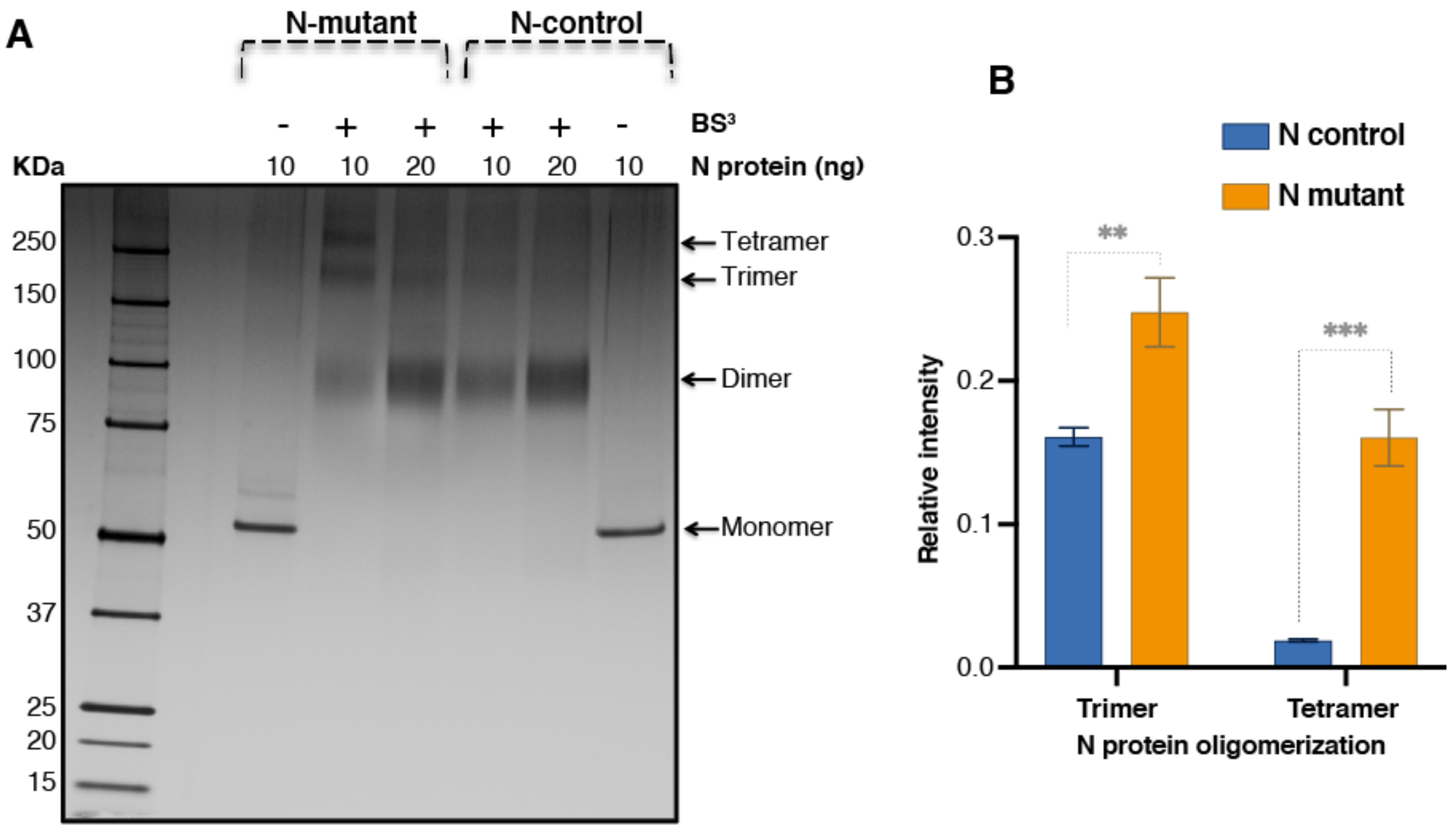


**Figure S11**

Oligomerization analysis of mutant and control N protein. A) $BS^3$ cross-linking (2mM) and SDS-PAGE analysis of the oligomerization forms of mutant and control N proteins. B) Densitometry analysis of bands corresponding to oligomeric forms (trimer and tetramer) was performed. Bar-plot represents the relative intensities from three independent experiments (as shown mean ± SD). (t-test, p value (0.000203***) and (0.00427**).

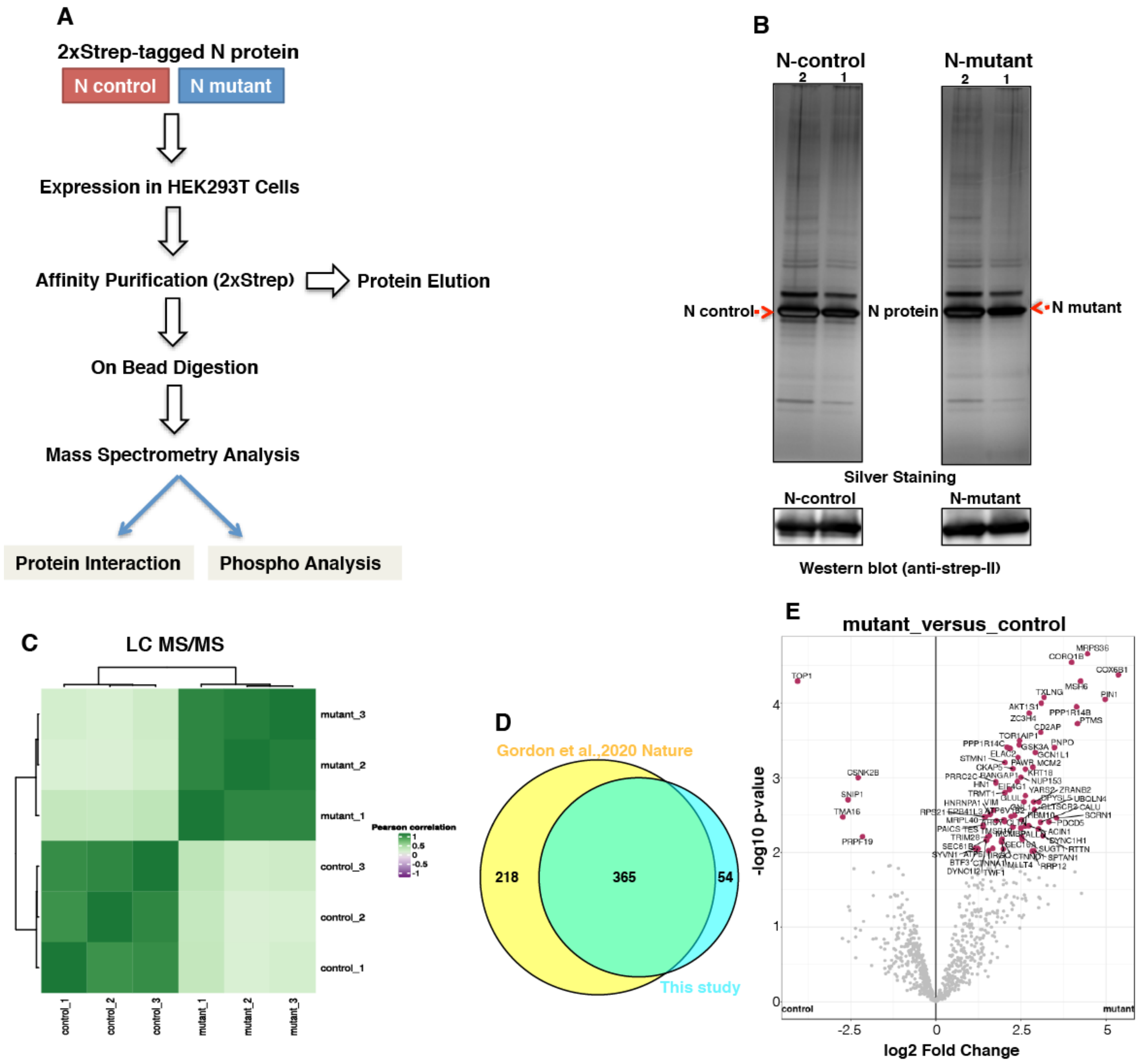


**Figure S12**

Affinity mass spectrometry (AP-MS) analysis of mutant and control SARS-CoV-2 N protein and host protein interaction. A) Sketch showing the workflow of affinity mass spectrometry procedure. HEK-293 cell expressing 2XStrep-tagged control and mutant N protein were used for MagStrep affinity purification. Purified proteins were separated on SDS-PAGE and subjected to silver staining and western blotting for confirmation. After confirmation, interacting proteins were analyzed by mass spectrometry. B) (Upper) Silver staining of control and mutant N protein associated host proteins (1 and 2 show two loading volume). (Lower) Western blot confirmation of N protein (mutant and control) using anti-Strep antibody. C) Correlation matrix of three replicates for control and mutant N protein AP-MS. D) Overlapping of identified N interacting proteins with N-interacting proteins reported in previous study[27] (Gordon et al., 2020 *Nature*). E) Volcano plot displaying the differential interactions of pairwise comparisons (mutant_vs_control) in – Log10 adj. p-values vs. the Log2 protein fold change. Proteins with statistically significant (Adjusted p-value <= 0.05, and Log fold change >= 1) difference between mutant and control AP-MS conditions are highlighted.

### Figure S13

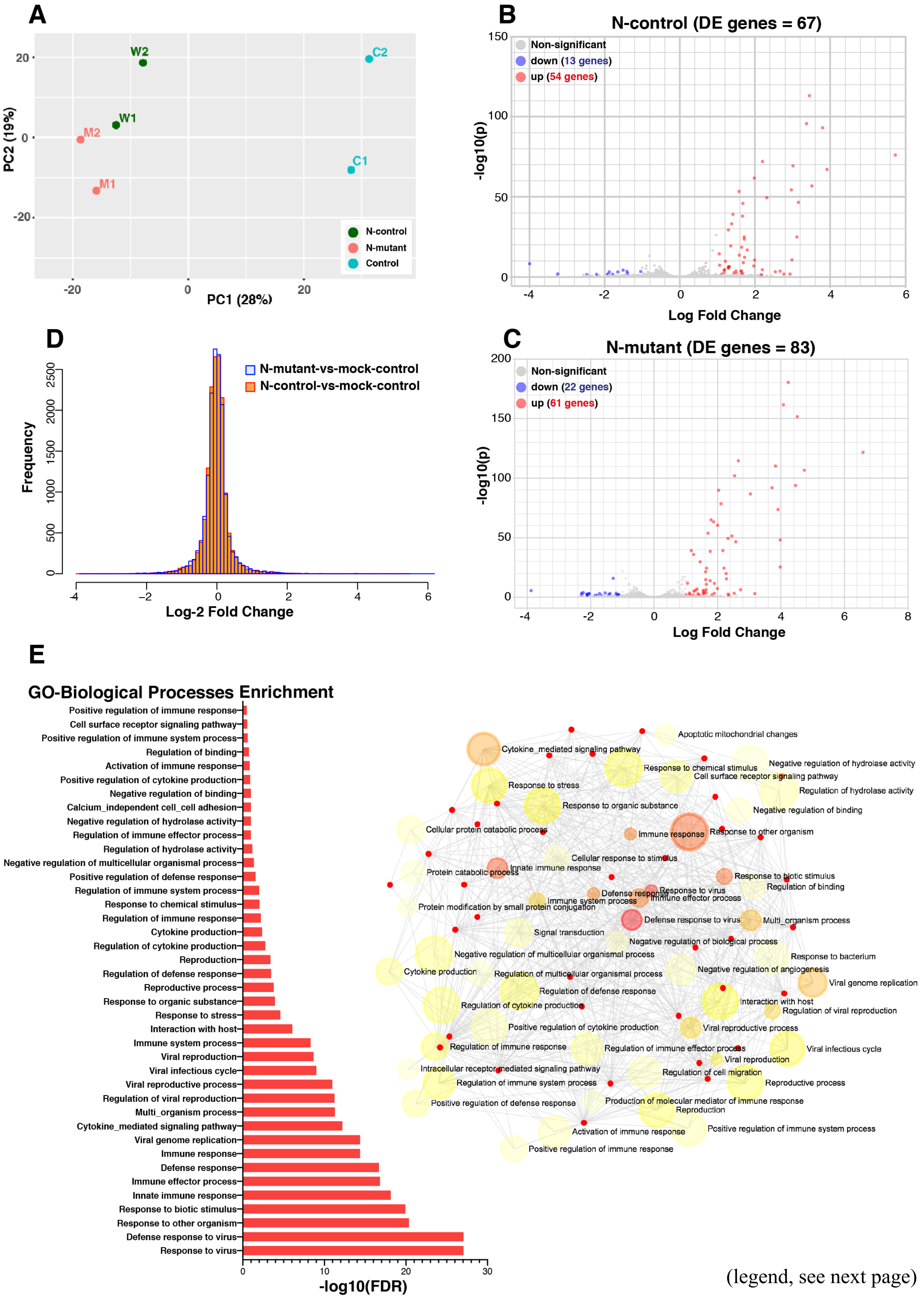


(legend, see next page)

**Figure S13. Transcriptomic analysis of mutant and control N transfected host cells**.
**A**) PCA on transcriptome of HEK293T cells transfected with plasmids expressing the full-length N-control and N-mutant protein along with mock control. **B-C**) Volcano-plot showing differentially expressed (DE) genes based on a filtering criterion of adj p-value $< 0.05$ and fold-change cutoff $\geq 1$) as determined by the method EdgeR in NetworkAnalyst tool. X-axis depicts log2 fold-change of DE genes and Y-axis depicts –log10 P-value. Genes with significant up-regulation are shown in red and dow-regulated are shown in blue. All other non- significant genes are shown in gray. **D**) Plot showing the distribution of log2-fold changes in both N-mutant and N-control conditions. **E**) GO enrichment analysis of all up-regulated genes in the N-mutant condition. The enriched GO BP (Biological Processes) terms are displayed by plotting against the –log10 of the false discovery rate (FDR q value). The enriched terms display an interconnected network with overlapping gene sets (from the list). Each node represents an enriched term and colored by its FDR q value (as shown in the bar-chart). The size of each node corresponds to number of linked genes from the list. The red dots shows up-regulated genes as shown in the Table S7.
