## Supplementary Table S1 for "Saudi Arabian SARS-CoV-2 genomes implicate a mutant Nucleocapsid protein in modulating host interactions and increased viral load in COVID-19 patients"

| sample ID | sample Name | GISAID accession |
|---|---|---|
| 3 | SaudiArabia/KAUST-Makkah03/2020 | EPI_ISL_437459 |
| 4 | SaudiArabia/KAUST-Makkah04/2020 | EPI_ISL_437460 |
| 5 | SaudiArabia/KAUST-Makkah05/2020 | EPI_ISL_437461 |
| 6 | SaudiArabia/KAUST-Makkah06/2020 | EPI_ISL_437462 |
| 9 | SaudiArabia/KAUST-Makkah09/2020 | EPI_ISL_437463 |
| 10 | SaudiArabia/KAUST-Makkah10/2020 | EPI_ISL_437464 |
| 11 | SaudiArabia/KAUST-MAKKAH11/2020 | EPI_ISL_678080 |
| 12 | SaudiArabia/KAUST-Makkah12/2020 | EPI_ISL_437465 |
| 13 | SaudiArabia/KAUST-Makkah13/2020 | EPI_ISL_437466 |
| 18 | SaudiArabia/KAUST-Makkah18/2020 | EPI_ISL_437467 |
| 19 | SaudiArabia/KAUST-Makkah19/2020 | EPI_ISL_437481 |
| 20 | SaudiArabia/KAUST-Makkah20/2020 | EPI_ISL_437468 |
| 22 | SaudiArabia/KAUST-MADINAH22/2020 | EPI_ISL_751237 |
| 23 | SaudiArabia/KAUST-Madinah23/2020 | EPI_ISL_437482 |
| 24 | SaudiArabia/KAUST-Madinah24/2020 | EPI_ISL_437483 |
| 25 | SaudiArabia/KAUST-Madinah25/2020 | EPI_ISL_437469 |
| 26 | SaudiArabia/KAUST-MAKKAH26/2020 | EPI_ISL_678141 |
| 27 | SaudiArabia/KAUST-Madinah27/2020 | EPI_ISL_437484 |
| 30 | SaudiArabia/KAUST-Madinah30/2020 | EPI_ISL_437485 |
| 31 | SaudiArabia/KAUST-Madinah31/2020 | EPI_ISL_437486 |
| 34 | SaudiArabia/KAUST-Madinah34/2020 | EPI_ISL_437470 |
| 38 | SaudiArabia/KAUST-Madinah38/2020 | EPI_ISL_437471 |
| 40 | SaudiArabia/KAUST-MADINAH40/2020 | EPI_ISL_677964 |
| 41 | SaudiArabia/KAUST-Madinah41/2020 | EPI_ISL_437472 |
| 42 | SaudiArabia/KAUST-MADINAH42/2020 | EPI_ISL_677976 |
| 44 | SaudiArabia/KAUST-Madinah44/2020 | EPI_ISL_437473 |
| 45 | SaudiArabia/KAUST-Madinah45/2020 | EPI_ISL_437474 |
| 47 | SaudiArabia/KAUST-Madinah47/2020 | EPI_ISL_437487 |
| 48 | SaudiArabia/KAUST-Madinah48/2020 | EPI_ISL_437488 |
| 49 | SaudiArabia/KAUST-Madinah49/2020 | EPI_ISL_437489 |
| 50 | SaudiArabia/KAUST-Madinah50/2020 | EPI_ISL_437490 |
| 51 | SaudiArabia/KAUST-Madinah51/2020 | EPI_ISL_437491 |
| 53 | SaudiArabia/KAUST-Madinah53/2020 | EPI_ISL_437492 |
| 54 | SaudiArabia/KAUST-Madinah54/2020 | EPI_ISL_437493 |
| 55 | SaudiArabia/KAUST-Madinah55/2020 | EPI_ISL_437494 |
| 57 | SaudiArabia/KAUST-Madinah57/2020 | EPI_ISL_437495 |
| 59 | SaudiArabia/KAUST-Jeddah59/2020 | EPI_ISL_437496 |
| 60 | SaudiArabia/KAUST-Jeddah60/2020 | EPI_ISL_437497 |
| 61 | SaudiArabia/KAUST-Jeddah61/2020 | EPI_ISL_437754 |
| 63 | SaudiArabia/KAUST-JEDDAH63/2020 | EPI_ISL_677940 |
| 64 | SaudiArabia/KAUST-Jeddah64/2020 | EPI_ISL_437475 |
| 66 | SaudiArabia/KAUST-Jeddah66/2020 | EPI_ISL_437755 |
| 68 | SaudiArabia/KAUST-Jeddah68/2020 | EPI_ISL_437756 |
| 70 | SaudiArabia/KAUST-Jeddah70/2020 | EPI_ISL_437757 |

| | | |
|---|---|---|
| 73 | SaudiArabia/KAUST-Jeddah73/2020 | EPI_ISL_437758 |
| 75 | SaudiArabia/KAUST-Jeddah75/2020 | EPI_ISL_437759 |
| 76 | SaudiArabia/KAUST-Jeddah76/2020 | EPI_ISL_437760 |
| 80 | SaudiArabia/KAUST-MAKKAH80/2020 | EPI_ISL_678041 |
| 90 | SaudiArabia/KAUST-Makkah90/2020 | EPI_ISL_437761 |
| 97 | SaudiArabia/KAUST-Makkah97/2020 | EPI_ISL_437762 |
| 104 | SaudiArabia/KAUST-MAKKAH104/2020 | EPI_ISL_678078 |
| 105 | SaudiArabia/KAUST-MAKKAH105/2020 | EPI_ISL_678072 |
| 106 | SaudiArabia/KAUST-MAKKAH106/2020 | EPI_ISL_678079 |
| 107 | SaudiArabia/KAUST-MAKKAH107/2020 | EPI_ISL_678227 |
| 108 | SaudiArabia/KAUST-MAKKAH108/2020 | EPI_ISL_678228 |
| 111 | SaudiArabia/KAUST-MAKKAH111/2020 | EPI_ISL_678042 |
| 112 | SaudiArabia/KAUST-MAKKAH112/2020 | EPI_ISL_678081 |
| 117 | SaudiArabia/KAUST-MAKKAH117/2020 | EPI_ISL_678082 |
| 118 | SaudiArabia/KAUST-MAKKAH118/2020 | EPI_ISL_678076 |
| 121 | SaudiArabia/KAUST-MAKKAH121/2020 | EPI_ISL_678054 |
| 123 | SaudiArabia/KAUST-MAKKAH123/2020 | EPI_ISL_678062 |
| 124 | SaudiArabia/KAUST-Makkah124/2020 | EPI_ISL_437691 |
| 126 | SaudiArabia/KAUST-Makkah126/2020 | EPI_ISL_437692 |
| 138 | SaudiArabia/KAUST-Makkah138/2020 | EPI_ISL_437693 |
| 139 | SaudiArabia/KAUST-Makkah139/2020 | EPI_ISL_437694 |
| 142 | SaudiArabia/KAUST-Makkah142/2020 | EPI_ISL_437695 |
| 143 | SaudiArabia/KAUST-MAKKAH143/2020 | EPI_ISL_678229 |
| 144 | SaudiArabia/KAUST-MAKKAH144/2020 | EPI_ISL_678043 |
| 145 | SaudiArabia/KAUST-MAKKAH145/2020 | EPI_ISL_678063 |
| 146 | SaudiArabia/KAUST-MAKKAH146/2020 | EPI_ISL_678061 |
| 149 | SaudiArabia/KAUST-MAKKAH149/2020 | EPI_ISL_678057 |
| 150 | SaudiArabia/KAUST-MAKKAH150/2020 | EPI_ISL_678096 |
| 151 | SaudiArabia/KAUST-MAKKAH151/2020 | EPI_ISL_678052 |
| 155 | SaudiArabia/KAUST-Makkah155/2020 | EPI_ISL_437476 |
| 156 | SaudiArabia/KAUST-Makkah156/2020 | EPI_ISL_437696 |
| 157 | SaudiArabia/KAUST-MAKKAH157/2020 | EPI_ISL_678116 |
| 158 | SaudiArabia/KAUST-MAKKAH158/2020 | EPI_ISL_678117 |
| 159 | SaudiArabia/KAUST-Makkah159/2020 | EPI_ISL_437477 |
| 162 | SaudiArabia/KAUST-MAKKAH162/2020 | EPI_ISL_678127 |
| 163 | SaudiArabia/KAUST-Makkah163/2020 | EPI_ISL_437697 |
| 165 | SaudiArabia/KAUST-Makkah165/2020 | EPI_ISL_437478 |
| 166 | SaudiArabia/KAUST-Makkah166/2020 | EPI_ISL_437698 |
| 167 | SaudiArabia/KAUST-MAKKAH167/2020 | EPI_ISL_678135 |
| 168 | SaudiArabia/KAUST-Makkah168/2020 | EPI_ISL_437479 |
| 169 | SaudiArabia/KAUST-MAKKAH169/2020 | EPI_ISL_678048 |
| 170 | SaudiArabia/KAUST-MAKKAH170/2020 | EPI_ISL_678039 |
| 171 | SaudiArabia/KAUST-MAKKAH171/2020 | EPI_ISL_678058 |
| 172 | SaudiArabia/KAUST-MAKKAH172/2020 | EPI_ISL_678136 |
| 173 | SaudiArabia/KAUST-Makkah173/2020 | EPI_ISL_437699 |
| 176 | SaudiArabia/KAUST-MAKKAH176/2020 | EPI_ISL_678073 |

| | | |
|---|---|---|
| 177 | SaudiArabia/KAUST-Makkah177/2020 | EPI_ISL_437700 |
| 178 | SaudiArabia/KAUST-Makkah178/2020 | EPI_ISL_437701 |
| 179 | SaudiArabia/KAUST-Makkah179/2020 | EPI_ISL_437702 |
| 180 | SaudiArabia/KAUST-MAKKAH180/2020 | EPI_ISL_678230 |
| 181 | SaudiArabia/KAUST-Makkah181/2020 | EPI_ISL_437703 |
| 182 | SaudiArabia/KAUST-MAKKAH182/2020 | EPI_ISL_678065 |
| 183 | SaudiArabia/KAUST-Makkah183/2020 | EPI_ISL_437704 |
| 184 | SaudiArabia/KAUST-MAKKAH184/2020 | EPI_ISL_678040 |
| 185 | SaudiArabia/KAUST-MAKKAH185/2020 | EPI_ISL_678231 |
| 186 | SaudiArabia/KAUST-MAKKAH186/2020 | EPI_ISL_678232 |
| 188 | SaudiArabia/KAUST-Makkah188/2020 | EPI_ISL_437705 |
| 189 | SaudiArabia/KAUST-Makkah189/2020 | EPI_ISL_437706 |
| 190 | SaudiArabia/KAUST-Makkah190/2020 | EPI_ISL_437707 |
| 191 | SaudiArabia/KAUST-MAKKAH191/2020 | EPI_ISL_678074 |
| 192 | SaudiArabia/KAUST-MAKKAH192/2020 | EPI_ISL_678059 |
| 193 | SaudiArabia/KAUST-Makkah193/2020 | EPI_ISL_437708 |
| 194 | SaudiArabia/KAUST-Makkah194/2020 | EPI_ISL_437709 |
| 195 | SaudiArabia/KAUST-MAKKAH195/2020 | EPI_ISL_678055 |
| 196 | SaudiArabia/KAUST-Makkah196/2020 | EPI_ISL_437710 |
| 197 | SaudiArabia/KAUST-MAKKAH197/2020 | EPI_ISL_678233 |
| 198 | SaudiArabia/KAUST-Makkah198/2020 | EPI_ISL_437711 |
| 199 | SaudiArabia/KAUST-MAKKAH199/2020 | EPI_ISL_678077 |
| 200 | SaudiArabia/KAUST-MAKKAH200/2020 | EPI_ISL_678047 |
| 201 | SaudiArabia/KAUST-MAKKAH201/2020 | EPI_ISL_678137 |
| 202 | SaudiArabia/KAUST-Makkah202/2020 | EPI_ISL_437712 |
| 203 | SaudiArabia/KAUST-Makkah203/2020 | EPI_ISL_437713 |
| 204 | SaudiArabia/KAUST-Makkah204/2020 | EPI_ISL_437714 |
| 205 | SaudiArabia/KAUST-MAKKAH205/2020 | EPI_ISL_678068 |
| 206 | SaudiArabia/KAUST-MAKKAH206/2020 | EPI_ISL_678138 |
| 207 | SaudiArabia/KAUST-MAKKAH207/2020 | EPI_ISL_678071 |
| 208 | SaudiArabia/KAUST-Madinah208/2020 | EPI_ISL_437715 |
| 209 | SaudiArabia/KAUST-MADINAH209/2020 | EPI_ISL_678234 |
| 210 | SaudiArabia/KAUST-MADINAH210/2020 | EPI_ISL_678026 |
| 211 | SaudiArabia/KAUST-Madinah211/2020 | EPI_ISL_437716 |
| 213 | SaudiArabia/KAUST-Madinah213/2020 | EPI_ISL_437717 |
| 215 | SaudiArabia/KAUST-Makkah215/2020 | EPI_ISL_437718 |
| 216 | SaudiArabia/KAUST-MAKKAH216/2020 | EPI_ISL_678069 |
| 217 | SaudiArabia/KAUST-Makkah217/2020 | EPI_ISL_437719 |
| 218 | SaudiArabia/KAUST-Makkah218/2020 | EPI_ISL_437720 |
| 219 | SaudiArabia/KAUST-MAKKAH219/2020 | EPI_ISL_678060 |
| 220 | SaudiArabia/KAUST-Makkah220/2020 | EPI_ISL_437721 |
| 221 | SaudiArabia/KAUST-MAKKAH221/2020 | EPI_ISL_678075 |
| 222 | SaudiArabia/KAUST-MAKKAH222/2020 | EPI_ISL_678139 |
| 223 | SaudiArabia/KAUST-MAKKAH223/2020 | EPI_ISL_678066 |
| 224 | SaudiArabia/KAUST-MAKKAH224/2020 | EPI_ISL_678056 |
| 225 | SaudiArabia/KAUST-Makkah225/2020 | EPI_ISL_437722 |

| | | |
|---|---|---|
| 227 | SaudiArabia/KAUST-Makkah227/2020 | EPI_ISL_437723 |
| 228 | SaudiArabia/KAUST-Makkah228/2020 | EPI_ISL_437724 |
| 229 | SaudiArabia/KAUST-Makkah229/2020 | EPI_ISL_437725 |
| 230 | SaudiArabia/KAUST-Makkah230/2020 | EPI_ISL_437726 |
| 231 | SaudiArabia/KAUST-Makkah231/2020 | EPI_ISL_437727 |
| 232 | SaudiArabia/KAUST-Makkah232/2020 | EPI_ISL_437728 |
| 233 | SaudiArabia/KAUST-Makkah233/2020 | EPI_ISL_437729 |
| 234 | SaudiArabia/KAUST-Makkah234/2020 | EPI_ISL_437730 |
| 235 | SaudiArabia/KAUST-MAKKAH235/2020 | EPI_ISL_678140 |
| 236 | SaudiArabia/KAUST-Makkah236/2020 | EPI_ISL_437731 |
| 237 | SaudiArabia/KAUST-Makkah237/2020 | EPI_ISL_437732 |
| 238 | SaudiArabia/KAUST-Madinah238/2020 | EPI_ISL_437733 |
| 239 | SaudiArabia/KAUST-MADINAH239/2020 | EPI_ISL_678235 |
| 240 | SaudiArabia/KAUST-MADINAH240/2020 | EPI_ISL_677958 |
| 244 | SaudiArabia/KAUST-MADINAH244/2020 | EPI_ISL_677960 |
| 245 | SaudiArabia/KAUST-MADINAH245/2020 | EPI_ISL_677963 |
| 246 | SaudiArabia/KAUST-MADINAH246/2020 | EPI_ISL_677970 |
| 248 | SaudiArabia/KAUST-Madinah248/2020 | EPI_ISL_437734 |
| 249 | SaudiArabia/KAUST-Madinah249/2020 | EPI_ISL_437735 |
| 250 | SaudiArabia/KAUST-MADINAH250/2020 | EPI_ISL_678027 |
| 251 | SaudiArabia/KAUST-MADINAH251/2020 | EPI_ISL_677946 |
| 252 | SaudiArabia/KAUST-Madinah252/2020 | EPI_ISL_437736 |
| 253 | SaudiArabia/KAUST-Madinah253/2020 | EPI_ISL_437737 |
| 254 | SaudiArabia/KAUST-Madinah254/2020 | EPI_ISL_437738 |
| 255 | SaudiArabia/KAUST-Madinah255/2020 | EPI_ISL_437739 |
| 256 | SaudiArabia/KAUST-Madinah256/2020 | EPI_ISL_437740 |
| 257 | SaudiArabia/KAUST-Madinah257/2020 | EPI_ISL_437741 |
| 258 | SaudiArabia/KAUST-Madinah258/2020 | EPI_ISL_437742 |
| 259 | SaudiArabia/KAUST-MADINAH259/2020 | EPI_ISL_677968 |
| 260 | SaudiArabia/KAUST-Madinah260/2020 | EPI_ISL_437743 |
| 261 | SaudiArabia/KAUST-Madinah261/2020 | EPI_ISL_437744 |
| 262 | SaudiArabia/KAUST-Madinah262/2020 | EPI_ISL_437745 |
| 263 | SaudiArabia/KAUST-Madinah263/2020 | EPI_ISL_437746 |
| 264 | SaudiArabia/KAUST-Madinah264/2020 | EPI_ISL_437747 |
| 265 | SaudiArabia/KAUST-Madinah265/2020 | EPI_ISL_437748 |
| 266 | SaudiArabia/KAUST-Madinah266/2020 | EPI_ISL_437749 |
| 267 | SaudiArabia/KAUST-Madinah267/2020 | EPI_ISL_437750 |
| 268 | SaudiArabia/KAUST-MADINAH268/2020 | EPI_ISL_678028 |
| 269 | SaudiArabia/KAUST-Madinah269/2020 | EPI_ISL_437751 |
| 270 | SaudiArabia/KAUST-Madinah270/2020 | EPI_ISL_437752 |
| 272 | SaudiArabia/KAUST-Madinah272/2020 | EPI_ISL_437753 |
| 273 | SaudiArabia/KAUST-MADINAH273/2020 | EPI_ISL_513121 |
| 274 | SaudiArabia/KAUST-MADINAH274/2020 | EPI_ISL_513122 |
| 276 | SaudiArabia/KAUST-MADINAH276/2020 | EPI_ISL_513123 |
| 280 | SaudiArabia/KAUST-MADINAH280/2020 | EPI_ISL_513124 |
| 281 | SaudiArabia/KAUST-MADINAH281/2020 | EPI_ISL_513125 |

| | | |
|---|---|---|
| 282 | SaudiArabia/KAUST-MADINAH282/2020 | EPI_ISL_513126 |
| 284 | SaudiArabia/KAUST-MADINAH284/2020 | EPI_ISL_678029 |
| 290 | SaudiArabia/KAUST-MAKKAH290/2020 | EPI_ISL_513214 |
| 291 | SaudiArabia/KAUST-MAKKAH291/2020 | EPI_ISL_513215 |
| 293 | SaudiArabia/KAUST-MAKKAH293/2020 | EPI_ISL_513216 |
| 294 | SaudiArabia/KAUST-MAKKAH294/2020 | EPI_ISL_513217 |
| 295 | SaudiArabia/KAUST-MAKKAH295/2020 | EPI_ISL_513218 |
| 297 | SaudiArabia/KAUST-MAKKAH297/2020 | EPI_ISL_513219 |
| 299 | SaudiArabia/KAUST-MAKKAH299/2020 | EPI_ISL_513220 |
| 300 | SaudiArabia/KAUST-MAKKAH300/2020 | EPI_ISL_513221 |
| 301 | SaudiArabia/KAUST-MAKKAH301/2020 | EPI_ISL_513222 |
| 302 | SaudiArabia/KAUST-MAKKAH302/2020 | EPI_ISL_513223 |
| 303 | SaudiArabia/KAUST-MAKKAH303/2020 | EPI_ISL_513224 |
| 304 | SaudiArabia/KAUST-MAKKAH304/2020 | EPI_ISL_513225 |
| 309 | SaudiArabia/KAUST-MAKKAH309/2020 | EPI_ISL_513226 |
| 313 | SaudiArabia/KAUST-JEDDAH313/2020 | EPI_ISL_512908 |
| 317 | SaudiArabia/KAUST-JEDDAH317/2020 | EPI_ISL_512909 |
| 321 | SaudiArabia/KAUST-JEDDAH321/2020 | EPI_ISL_512910 |
| 322 | SaudiArabia/KAUST-JEDDAH322/2020 | EPI_ISL_512911 |
| 323 | SaudiArabia/KAUST-JEDDAH323/2020 | EPI_ISL_512912 |
| 324 | SaudiArabia/KAUST-JEDDAH324/2020 | EPI_ISL_512913 |
| 328 | SaudiArabia/KAUST-JEDDAH328/2020 | EPI_ISL_751221 |
| 329 | SaudiArabia/KAUST-JEDDAH329/2020 | EPI_ISL_512914 |
| 330 | SaudiArabia/KAUST-JEDDAH330/2020 | EPI_ISL_512915 |
| 331 | SaudiArabia/KAUST-JEDDAH331/2020 | EPI_ISL_512916 |
| 332 | SaudiArabia/KAUST-JEDDAH332/2020 | EPI_ISL_512917 |
| 333 | SaudiArabia/KAUST-MADINAH333/2020 | EPI_ISL_513127 |
| 335 | SaudiArabia/KAUST-MADINAH335/2020 | EPI_ISL_513128 |
| 342 | SaudiArabia/KAUST-MADINAH342/2020 | EPI_ISL_513129 |
| 343 | SaudiArabia/KAUST-MADINAH343/2020 | EPI_ISL_513130 |
| 344 | SaudiArabia/KAUST-MADINAH344/2020 | EPI_ISL_513131 |
| 346 | SaudiArabia/KAUST-MADINAH346/2020 | EPI_ISL_513132 |
| 347 | SaudiArabia/KAUST-MADINAH347/2020 | EPI_ISL_513133 |
| 354 | SaudiArabia/KAUST-MADINAH354/2020 | EPI_ISL_513134 |
| 355 | SaudiArabia/KAUST-MADINAH355/2020 | EPI_ISL_513135 |
| 357 | SaudiArabia/KAUST-MADINAH357/2020 | EPI_ISL_513136 |
| 360 | SaudiArabia/KAUST-MADINAH360/2020 | EPI_ISL_513137 |
| 361 | SaudiArabia/KAUST-MADINAH361/2020 | EPI_ISL_513138 |
| 362 | SaudiArabia/KAUST-MADINAH362/2020 | EPI_ISL_513139 |
| 363 | SaudiArabia/KAUST-MADINAH363/2020 | EPI_ISL_513140 |
| 364 | SaudiArabia/KAUST-MADINAH364/2020 | EPI_ISL_513141 |
| 368 | SaudiArabia/KAUST-MADINAH368/2020 | EPI_ISL_513142 |
| 378 | SaudiArabia/KAUST-MADINAH378/2020 | EPI_ISL_513143 |
| 380 | SaudiArabia/KAUST-MADINAH380/2020 | EPI_ISL_513144 |
| 384 | SaudiArabia/KAUST-MADINAH384/2020 | EPI_ISL_513145 |
| 387 | SaudiArabia/KAUST-MADINAH387/2020 | EPI_ISL_513146 |

| | | |
|---|---|---|
| 388 | SaudiArabia/KAUST-MADINAH388/2020 | EPI_ISL_751218 |
| 390 | SaudiArabia/KAUST-MADINAH390/2020 | EPI_ISL_636960 |
| 391 | SaudiArabia/KAUST-MADINAH391/2020 | EPI_ISL_513147 |
| 393 | SaudiArabia/KAUST-MADINAH393/2020 | EPI_ISL_513148 |
| 396 | SaudiArabia/KAUST-MADINAH396/2020 | EPI_ISL_513149 |
| 397 | SaudiArabia/KAUST-MADINAH397/2020 | EPI_ISL_513150 |
| 398 | SaudiArabia/KAUST-MADINAH398/2020 | EPI_ISL_513151 |
| 399 | SaudiArabia/KAUST-MADINAH399/2020 | EPI_ISL_513152 |
| 404 | SaudiArabia/KAUST-MADINAH404/2020 | EPI_ISL_513153 |
| 406 | SaudiArabia/KAUST-MADINAH406/2020 | EPI_ISL_513154 |
| 434 | SaudiArabia/KAUST-MADINAH434/2020 | EPI_ISL_513155 |
| 435 | SaudiArabia/KAUST-MADINAH435/2020 | EPI_ISL_513156 |
| 437 | SaudiArabia/KAUST-MADINAH437/2020 | EPI_ISL_513157 |
| 444 | SaudiArabia/KAUST-MADINAH444/2020 | EPI_ISL_513158 |
| 446 | SaudiArabia/KAUST-MADINAH446/2020 | EPI_ISL_513159 |
| 450 | SaudiArabia/KAUST-MADINAH450/2020 | EPI_ISL_513160 |
| 451 | SaudiArabia/KAUST-MADINAH451/2020 | EPI_ISL_678030 |
| 452 | SaudiArabia/KAUST-MADINAH452/2020 | EPI_ISL_513161 |
| 453 | SaudiArabia/KAUST-MADINAH453/2020 | EPI_ISL_678031 |
| 456 | SaudiArabia/KAUST-JEDDAH456/2020 | EPI_ISL_512918 |
| 457 | SaudiArabia/KAUST-JEDDAH457/2020 | EPI_ISL_512919 |
| 458 | SaudiArabia/KAUST-JEDDAH458/2020 | EPI_ISL_751502 |
| 459 | SaudiArabia/KAUST-JEDDAH459/2020 | EPI_ISL_636961 |
| 462 | SaudiArabia/KAUST-JEDDAH462/2020 | EPI_ISL_512920 |
| 466 | SaudiArabia/KAUST-JEDDAH466/2020 | EPI_ISL_512921 |
| 467 | SaudiArabia/KAUST-JEDDAH467/2020 | EPI_ISL_512922 |
| 468 | SaudiArabia/KAUST-JEDDAH468/2020 | EPI_ISL_512923 |
| 469 | SaudiArabia/KAUST-JEDDAH469/2020 | EPI_ISL_512924 |
| 470 | SaudiArabia/KAUST-JEDDAH470/2020 | EPI_ISL_512925 |
| 471 | SaudiArabia/KAUST-JEDDAH471/2020 | EPI_ISL_512926 |
| 473 | SaudiArabia/KAUST-JEDDAH473/2020 | EPI_ISL_512927 |
| 479 | SaudiArabia/KAUST-JEDDAH479/2020 | EPI_ISL_512928 |
| 482 | SaudiArabia/KAUST-JEDDAH482/2020 | EPI_ISL_512929 |
| 483 | SaudiArabia/KAUST-JEDDAH483/2020 | EPI_ISL_512930 |
| 486 | SaudiArabia/KAUST-JEDDAH486/2020 | EPI_ISL_512931 |
| 487 | SaudiArabia/KAUST-JEDDAH487/2020 | EPI_ISL_512932 |
| 488 | SaudiArabia/KAUST-JEDDAH488/2020 | EPI_ISL_512933 |
| 489 | SaudiArabia/KAUST-JEDDAH489/2020 | EPI_ISL_751224 |
| 490 | SaudiArabia/KAUST-JEDDAH490/2020 | EPI_ISL_512934 |
| 492 | SaudiArabia/KAUST-JEDDAH492/2020 | EPI_ISL_512935 |
| 493 | SaudiArabia/KAUST-JEDDAH493/2020 | EPI_ISL_512936 |
| 494 | SaudiArabia/KAUST-JEDDAH494/2020 | EPI_ISL_512937 |
| 496 | SaudiArabia/KAUST-JEDDAH496/2020 | EPI_ISL_512938 |
| 497 | SaudiArabia/KAUST-JEDDAH497/2020 | EPI_ISL_512939 |
| 498 | SaudiArabia/KAUST-JEDDAH498/2020 | EPI_ISL_512940 |
| 500 | SaudiArabia/KAUST-JEDDAH500/2020 | EPI_ISL_677937 |

| | | |
|---|---|---|
| 504 | SaudiArabia/KAUST-JEDDAH504/2020 | EPI_ISL_512941 |
| 505 | SaudiArabia/KAUST-JEDDAH505/2020 | EPI_ISL_512942 |
| 506 | SaudiArabia/KAUST-JEDDAH506/2020 | EPI_ISL_512943 |
| 510 | SaudiArabia/KAUST-MADINAH510/2020 | EPI_ISL_751204 |
| 511 | SaudiArabia/KAUST-MADINAH511/2020 | EPI_ISL_513162 |
| 513 | SaudiArabia/KAUST-MADINAH513/2020 | EPI_ISL_513163 |
| 515 | SaudiArabia/KAUST-MADINAH515/2020 | EPI_ISL_513164 |
| 516 | SaudiArabia/KAUST-MADINAH516/2020 | EPI_ISL_513165 |
| 519 | SaudiArabia/KAUST-MADINAH519/2020 | EPI_ISL_513166 |
| 521 | SaudiArabia/KAUST-MADINAH521/2020 | EPI_ISL_513167 |
| 525 | SaudiArabia/KAUST-JEDDAH525/2020 | EPI_ISL_512944 |
| 528 | SaudiArabia/KAUST-JEDDAH528/2020 | EPI_ISL_512945 |
| 529 | SaudiArabia/KAUST-JEDDAH529/2020 | EPI_ISL_512946 |
| 531 | SaudiArabia/KAUST-JEDDAH531/2020 | EPI_ISL_512947 |
| 533 | SaudiArabia/KAUST-JEDDAH533/2020 | EPI_ISL_677938 |
| 534 | SaudiArabia/KAUST-JEDDAH534/2020 | EPI_ISL_512948 |
| 535 | SaudiArabia/KAUST-JEDDAH535/2020 | EPI_ISL_512949 |
| 536 | SaudiArabia/KAUST-JEDDAH536/2020 | EPI_ISL_512950 |
| 538 | SaudiArabia/KAUST-JEDDAH538/2020 | EPI_ISL_512951 |
| 539 | SaudiArabia/KAUST-JEDDAH539/2020 | EPI_ISL_512952 |
| 542 | SaudiArabia/KAUST-JEDDAH542/2020 | EPI_ISL_512953 |
| 544 | SaudiArabia/KAUST-JEDDAH544/2020 | EPI_ISL_512954 |
| 545 | SaudiArabia/KAUST-JEDDAH545/2020 | EPI_ISL_512955 |
| 546 | SaudiArabia/KAUST-JEDDAH546/2020 | EPI_ISL_512956 |
| 547 | SaudiArabia/KAUST-JEDDAH547/2020 | EPI_ISL_512957 |
| 554 | SaudiArabia/KAUST-JEDDAH554/2020 | EPI_ISL_512958 |
| 558 | SaudiArabia/KAUST-JEDDAH558/2020 | EPI_ISL_512959 |
| 559 | SaudiArabia/KAUST-JEDDAH559/2020 | EPI_ISL_512960 |
| 560 | SaudiArabia/KAUST-JEDDAH560/2020 | EPI_ISL_677939 |
| 561 | SaudiArabia/KAUST-JEDDAH561/2020 | EPI_ISL_512961 |
| 562 | SaudiArabia/KAUST-JEDDAH562/2020 | EPI_ISL_512962 |
| 566 | SaudiArabia/KAUST-JEDDAH566/2020 | EPI_ISL_512963 |
| 569 | SaudiArabia/KAUST-JEDDAH569/2020 | EPI_ISL_512964 |
| 570 | SaudiArabia/KAUST-JEDDAH570/2020 | EPI_ISL_512965 |
| 572 | SaudiArabia/KAUST-JEDDAH572/2020 | EPI_ISL_512966 |
| 573 | SaudiArabia/KAUST-JEDDAH573/2020 | EPI_ISL_512967 |
| 574 | SaudiArabia/KAUST-JEDDAH574/2020 | EPI_ISL_512968 |
| 576 | SaudiArabia/KAUST-JEDDAH576/2020 | EPI_ISL_512969 |
| 577 | SaudiArabia/KAUST-JEDDAH577/2020 | EPI_ISL_512970 |
| 578 | SaudiArabia/KAUST-JEDDAH578/2020 | EPI_ISL_512971 |
| 580 | SaudiArabia/KAUST-JEDDAH580/2020 | EPI_ISL_512972 |
| 581 | SaudiArabia/KAUST-JEDDAH581/2020 | EPI_ISL_512973 |
| 582 | SaudiArabia/KAUST-JEDDAH582/2020 | EPI_ISL_512974 |
| 583 | SaudiArabia/KAUST-JEDDAH583/2020 | EPI_ISL_512975 |
| 586 | SaudiArabia/KAUST-JEDDAH586/2020 | EPI_ISL_512976 |
| 587 | SaudiArabia/KAUST-JEDDAH587/2020 | EPI_ISL_512977 |

| | | |
|---|---|---|
| 588 | SaudiArabia/KAUST-JEDDAH588/2020 | EPI_ISL_512978 |
| 589 | SaudiArabia/KAUST-JEDDAH589/2020 | EPI_ISL_512979 |
| 590 | SaudiArabia/KAUST-JEDDAH590/2020 | EPI_ISL_512980 |
| 591 | SaudiArabia/KAUST-JEDDAH591/2020 | EPI_ISL_512981 |
| 592 | SaudiArabia/KAUST-JEDDAH592/2020 | EPI_ISL_512982 |
| 594 | SaudiArabia/KAUST-JEDDAH594/2020 | EPI_ISL_512983 |
| 595 | SaudiArabia/KAUST-JEDDAH595/2020 | EPI_ISL_512984 |
| 599 | SaudiArabia/KAUST-MADINAH599/2020 | EPI_ISL_513168 |
| 602 | SaudiArabia/KAUST-MADINAH602/2020 | EPI_ISL_678032 |
| 603 | SaudiArabia/KAUST-MADINAH603/2020 | EPI_ISL_513169 |
| 605 | SaudiArabia/KAUST-MADINAH605/2020 | EPI_ISL_513170 |
| 606 | SaudiArabia/KAUST-MADINAH606/2020 | EPI_ISL_513171 |
| 609 | SaudiArabia/KAUST-MADINAH609/2020 | EPI_ISL_513172 |
| 611 | SaudiArabia/KAUST-JEDDAH611/2020 | EPI_ISL_512985 |
| 613 | SaudiArabia/KAUST-JEDDAH613/2020 | EPI_ISL_512986 |
| 615 | SaudiArabia/KAUST-JEDDAH615/2020 | EPI_ISL_512987 |
| 632 | SaudiArabia/KAUST-JEDDAH632/2020 | EPI_ISL_512988 |
| 636 | SaudiArabia/KAUST-MADINAH636/2020 | EPI_ISL_678033 |
| 644 | SaudiArabia/KAUST-MADINAH644/2020 | EPI_ISL_513173 |
| 647 | SaudiArabia/KAUST-MADINAH647/2020 | EPI_ISL_513174 |
| 654 | SaudiArabia/KAUST-MADINAH654/2020 | EPI_ISL_678034 |
| 655 | SaudiArabia/KAUST-MADINAH655/2020 | EPI_ISL_513175 |
| 662 | SaudiArabia/KAUST-MADINAH662/2020 | EPI_ISL_678035 |
| 663 | SaudiArabia/KAUST-MADINAH663/2020 | EPI_ISL_678036 |
| 664 | SaudiArabia/KAUST-MADINAH664/2020 | EPI_ISL_677947 |
| 665 | SaudiArabia/KAUST-MADINAH665/2020 | EPI_ISL_678037 |
| 668 | SaudiArabia/KAUST-MADINAH668/2020 | EPI_ISL_751212 |
| 669 | SaudiArabia/KAUST-MADINAH669/2020 | EPI_ISL_677973 |
| 670 | SaudiArabia/KAUST-MADINAH670/2020 | EPI_ISL_677966 |
| 673 | SaudiArabia/KAUST-MADINAH673/2020 | EPI_ISL_678038 |
| 680 | SaudiArabia/KAUST-JEDDAH680/2020 | EPI_ISL_677941 |
| 682 | SaudiArabia/KAUST-JEDDAH682/2020 | EPI_ISL_677942 |
| 683 | SaudiArabia/KAUST-MADINAH683/2020 | EPI_ISL_677945 |
| 684 | SaudiArabia/KAUST-MADINAH684/2020 | EPI_ISL_513176 |
| 687 | SaudiArabia/KAUST-MADINAH687/2020 | EPI_ISL_513177 |
| 688 | SaudiArabia/KAUST-MADINAH688/2020 | EPI_ISL_513178 |
| 690 | SaudiArabia/KAUST-MADINAH690/2020 | EPI_ISL_677965 |
| 691 | SaudiArabia/KAUST-MADINAH691/2020 | EPI_ISL_513179 |
| 694 | SaudiArabia/KAUST-MADINAH694/2020 | EPI_ISL_513180 |
| 702 | SaudiArabia/KAUST-MADINAH702/2020 | EPI_ISL_513181 |
| 704 | SaudiArabia/KAUST-MADINAH704/2020 | EPI_ISL_513182 |
| 708 | SaudiArabia/KAUST-MADINAH708/2020 | EPI_ISL_677955 |
| 710 | SaudiArabia/KAUST-MADINAH710/2020 | EPI_ISL_513183 |
| 711 | SaudiArabia/KAUST-MADINAH711/2020 | EPI_ISL_513184 |
| 712 | SaudiArabia/KAUST-MADINAH712/2020 | EPI_ISL_513185 |
| 713 | SaudiArabia/KAUST-MADINAH713/2020 | EPI_ISL_513186 |

| | | |
|---|---|---|
| 715 | SaudiArabia/KAUST-MADINAH715/2020 | EPI_ISL_513187 |
| 716 | SaudiArabia/KAUST-MADINAH716/2020 | EPI_ISL_513188 |
| 717 | SaudiArabia/KAUST-MADINAH717/2020 | EPI_ISL_513189 |
| 719 | SaudiArabia/KAUST-JEDDAH719/2020 | EPI_ISL_677943 |
| 720 | SaudiArabia/KAUST-JEDDAH720/2020 | EPI_ISL_512989 |
| 721 | SaudiArabia/KAUST-JEDDAH721/2020 | EPI_ISL_512990 |
| 722 | SaudiArabia/KAUST-JEDDAH722/2020 | EPI_ISL_512991 |
| 723 | SaudiArabia/KAUST-JEDDAH723/2020 | EPI_ISL_512992 |
| 725 | SaudiArabia/KAUST-MAKKAH725/2020 | EPI_ISL_513227 |
| 726 | SaudiArabia/KAUST-MAKKAH726/2020 | EPI_ISL_513228 |
| 727 | SaudiArabia/KAUST-MAKKAH727/2020 | EPI_ISL_513229 |
| 729 | SaudiArabia/KAUST-MAKKAH729/2020 | EPI_ISL_678142 |
| 730 | SaudiArabia/KAUST-MAKKAH730/2020 | EPI_ISL_513230 |
| 733 | SaudiArabia/KAUST-MAKKAH733/2020 | EPI_ISL_751207 |
| 734 | SaudiArabia/KAUST-MAKKAH734/2020 | EPI_ISL_751205 |
| 735 | SaudiArabia/KAUST-MAKKAH735/2020 | EPI_ISL_513231 |
| 737 | SaudiArabia/KAUST-MAKKAH737/2020 | EPI_ISL_513232 |
| 741 | SaudiArabia/KAUST-MAKKAH741/2020 | EPI_ISL_513233 |
| 742 | SaudiArabia/KAUST-JEDDAH742/2020 | EPI_ISL_677944 |
| 743 | SaudiArabia/KAUST-JEDDAH743/2020 | EPI_ISL_512993 |
| 746 | SaudiArabia/KAUST-JEDDAH746/2020 | EPI_ISL_512994 |
| 747 | SaudiArabia/KAUST-JEDDAH747/2020 | EPI_ISL_512995 |
| 748 | SaudiArabia/KAUST-JEDDAH748/2020 | EPI_ISL_512996 |
| 754 | SaudiArabia/KAUST-JEDDAH754/2020 | EPI_ISL_512997 |
| 755 | SaudiArabia/KAUST-JEDDAH755/2020 | EPI_ISL_512998 |
| 756 | SaudiArabia/KAUST-JEDDAH756/2020 | EPI_ISL_512999 |
| 757 | SaudiArabia/KAUST-JEDDAH757/2020 | EPI_ISL_513000 |
| 759 | SaudiArabia/KAUST-JEDDAH759/2020 | EPI_ISL_513001 |
| 760 | SaudiArabia/KAUST-MAKKAH760/2020 | EPI_ISL_513234 |
| 761 | SaudiArabia/KAUST-MAKKAH761/2020 | EPI_ISL_513235 |
| 762 | SaudiArabia/KAUST-MAKKAH762/2020 | EPI_ISL_513236 |
| 766 | SaudiArabia/KAUST-MAKKAH766/2020 | EPI_ISL_513237 |
| 768 | SaudiArabia/KAUST-MAKKAH768/2020 | EPI_ISL_513238 |
| 769 | SaudiArabia/KAUST-MAKKAH769/2020 | EPI_ISL_513239 |
| 770 | SaudiArabia/KAUST-MAKKAH770/2020 | EPI_ISL_513240 |
| 771 | SaudiArabia/KAUST-MAKKAH771/2020 | EPI_ISL_513241 |
| 772 | SaudiArabia/KAUST-MAKKAH772/2020 | EPI_ISL_513242 |
| 773 | SaudiArabia/KAUST-MAKKAH773/2020 | EPI_ISL_513243 |
| 774 | SaudiArabia/KAUST-MAKKAH774/2020 | EPI_ISL_513244 |
| 775 | SaudiArabia/KAUST-MAKKAH775/2020 | EPI_ISL_678143 |
| 776 | SaudiArabia/KAUST-MAKKAH776/2020 | EPI_ISL_513245 |
| 777 | SaudiArabia/KAUST-MAKKAH777/2020 | EPI_ISL_513246 |
| 778 | SaudiArabia/KAUST-MAKKAH778/2020 | EPI_ISL_513247 |
| 779 | SaudiArabia/KAUST-MAKKAH779/2020 | EPI_ISL_513248 |
| 781 | SaudiArabia/KAUST-MAKKAH781/2020 | EPI_ISL_513249 |
| 782 | SaudiArabia/KAUST-MAKKAH782/2020 | EPI_ISL_513250 |

| | | |
|---|---|---|
| 783 | SaudiArabia/KAUST-MAKKAH783/2020 | EPI_ISL_513251 |
| 784 | SaudiArabia/KAUST-MAKKAH784/2020 | EPI_ISL_513252 |
| 785 | SaudiArabia/KAUST-MAKKAH785/2020 | EPI_ISL_513253 |
| 786 | SaudiArabia/KAUST-MAKKAH786/2020 | EPI_ISL_513254 |
| 789 | SaudiArabia/KAUST-MAKKAH789/2020 | EPI_ISL_678044 |
| 794 | SaudiArabia/KAUST-MAKKAH794/2020 | EPI_ISL_513255 |
| 798 | SaudiArabia/KAUST-MAKKAH798/2020 | EPI_ISL_678144 |
| 799 | SaudiArabia/KAUST-MAKKAH799/2020 | EPI_ISL_513256 |
| 802 | SaudiArabia/KAUST-MAKKAH802/2020 | EPI_ISL_513257 |
| 808 | SaudiArabia/KAUST-MAKKAH808/2020 | EPI_ISL_513258 |
| 809 | SaudiArabia/KAUST-MAKKAH809/2020 | EPI_ISL_513259 |
| 810 | SaudiArabia/KAUST-MAKKAH810/2020 | EPI_ISL_513260 |
| 812 | SaudiArabia/KAUST-MAKKAH812/2020 | EPI_ISL_513261 |
| 813 | SaudiArabia/KAUST-MAKKAH813/2020 | EPI_ISL_513262 |
| 814 | SaudiArabia/KAUST-MAKKAH814/2020 | EPI_ISL_513263 |
| 815 | SaudiArabia/KAUST-MAKKAH815/2020 | EPI_ISL_513264 |
| 819 | SaudiArabia/KAUST-MAKKAH819/2020 | EPI_ISL_678145 |
| 825 | SaudiArabia/KAUST-MAKKAH825/2020 | EPI_ISL_678146 |
| 826 | SaudiArabia/KAUST-JEDDAH826/2020 | EPI_ISL_513002 |
| 828 | SaudiArabia/KAUST-JEDDAH828/2020 | EPI_ISL_513003 |
| 830 | SaudiArabia/KAUST-JEDDAH830/2020 | EPI_ISL_513004 |
| 831 | SaudiArabia/KAUST-JEDDAH831/2020 | EPI_ISL_513005 |
| 832 | SaudiArabia/KAUST-JEDDAH832/2020 | EPI_ISL_751208 |
| 833 | SaudiArabia/KAUST-JEDDAH833/2020 | EPI_ISL_513006 |
| 834 | SaudiArabia/KAUST-JEDDAH834/2020 | EPI_ISL_513007 |
| 835 | SaudiArabia/KAUST-JEDDAH835/2020 | EPI_ISL_513008 |
| 836 | SaudiArabia/KAUST-JEDDAH836/2020 | EPI_ISL_513009 |
| 837 | SaudiArabia/KAUST-JEDDAH837/2020 | EPI_ISL_513010 |
| 838 | SaudiArabia/KAUST-JEDDAH838/2020 | EPI_ISL_513011 |
| 839 | SaudiArabia/KAUST-jeddah839/2020 | EPI_ISL_636972 |
| 840 | SaudiArabia/KAUST-JEDDAH840/2020 | EPI_ISL_513012 |
| 841 | SaudiArabia/KAUST-JEDDAH841/2020 | EPI_ISL_513013 |
| 842 | SaudiArabia/KAUST-JEDDAH842/2020 | EPI_ISL_513014 |
| 843 | SaudiArabia/KAUST-JEDDAH843/2020 | EPI_ISL_513015 |
| 844 | SaudiArabia/KAUST-JEDDAH844/2020 | EPI_ISL_513016 |
| 845 | SaudiArabia/KAUST-JEDDAH845/2020 | EPI_ISL_513017 |
| 846 | SaudiArabia/KAUST-JEDDAH846/2020 | EPI_ISL_513018 |
| 847 | SaudiArabia/KAUST-JEDDAH847/2020 | EPI_ISL_513019 |
| 850 | SaudiArabia/KAUST-MADINAH850/2020 | EPI_ISL_513190 |
| 851 | SaudiArabia/KAUST-MADINAH851/2020 | EPI_ISL_513191 |
| 854 | SaudiArabia/KAUST-MADINAH854/2020 | EPI_ISL_513192 |
| 857 | SaudiArabia/KAUST-MADINAH857/2020 | EPI_ISL_513193 |
| 858 | SaudiArabia/KAUST-MADINAH858/2020 | EPI_ISL_513194 |
| 859 | SaudiArabia/KAUST-MADINAH859/2020 | EPI_ISL_513195 |
| 860 | SaudiArabia/KAUST-MADINAH860/2020 | EPI_ISL_513196 |
| 862 | SaudiArabia/KAUST-MADINAH862/2020 | EPI_ISL_513197 |

| | | |
|---|---|---|
| 866 | SaudiArabia/KAUST-JEDDAH866/2020 | EPI_ISL_513020 |
| 867 | SaudiArabia/KAUST-JEDDAH867/2020 | EPI_ISL_513021 |
| 868 | SaudiArabia/KAUST-JEDDAH868/2020 | EPI_ISL_513022 |
| 869 | SaudiArabia/KAUST-JEDDAH869/2020 | EPI_ISL_513023 |
| 870 | SaudiArabia/KAUST-JEDDAH870/2020 | EPI_ISL_513024 |
| 871 | SaudiArabia/KAUST-JEDDAH871/2020 | EPI_ISL_513025 |
| 872 | SaudiArabia/KAUST-JEDDAH872/2020 | EPI_ISL_513026 |
| 873 | SaudiArabia/KAUST-JEDDAH873/2020 | EPI_ISL_513027 |
| 875 | SaudiArabia/KAUST-JEDDAH875/2020 | EPI_ISL_513028 |
| 876 | SaudiArabia/KAUST-JEDDAH876/2020 | EPI_ISL_636962 |
| 877 | SaudiArabia/KAUST-JEDDAH877/2020 | EPI_ISL_513029 |
| 878 | SaudiArabia/KAUST-JEDDAH878/2020 | EPI_ISL_513030 |
| 879 | SaudiArabia/KAUST-JEDDAH879/2020 | EPI_ISL_513031 |
| 880 | SaudiArabia/KAUST-JEDDAH880/2020 | EPI_ISL_636963 |
| 881 | SaudiArabia/KAUST-JEDDAH881/2020 | EPI_ISL_751219 |
| 882 | SaudiArabia/KAUST-JEDDAH882/2020 | EPI_ISL_513032 |
| 883 | SaudiArabia/KAUST-JEDDAH883/2020 | EPI_ISL_513033 |
| 884 | SaudiArabia/KAUST-JEDDAH884/2020 | EPI_ISL_513034 |
| 885 | SaudiArabia/KAUST-JEDDAH885/2020 | EPI_ISL_751220 |
| 886 | SaudiArabia/KAUST-JEDDAH886/2020 | EPI_ISL_513035 |
| 887 | SaudiArabia/KAUST-JEDDAH887/2020 | EPI_ISL_513036 |
| 888 | SaudiArabia/KAUST-JEDDAH888/2020 | EPI_ISL_513037 |
| 889 | SaudiArabia/KAUST-JEDDAH889/2020 | EPI_ISL_513038 |
| 890 | SaudiArabia/KAUST-JEDDAH890/2020 | EPI_ISL_513039 |
| 891 | SaudiArabia/KAUST-JEDDAH891/2020 | EPI_ISL_513040 |
| 892 | SaudiArabia/KAUST-MADINAH892/2020 | EPI_ISL_513198 |
| 893 | SaudiArabia/KAUST-MADINAH893/2020 | EPI_ISL_636964 |
| 896 | SaudiArabia/KAUST-MADINAH896/2020 | EPI_ISL_513199 |
| 898 | SaudiArabia/KAUST-MADINAH898/2020 | EPI_ISL_513200 |
| 900 | SaudiArabia/KAUST-JEDDAH900/2020 | EPI_ISL_513041 |
| 903 | SaudiArabia/KAUST-JEDDAH903/2020 | EPI_ISL_513042 |
| 905 | SaudiArabia/KAUST-JEDDAH905/2020 | EPI_ISL_513043 |
| 911 | SaudiArabia/KAUST-JEDDAH911/2020 | EPI_ISL_513044 |
| 912 | SaudiArabia/KAUST-JEDDAH912/2020 | EPI_ISL_513045 |
| 913 | SaudiArabia/KAUST-JEDDAH913/2020 | EPI_ISL_513046 |
| 916 | SaudiArabia/KAUST-JEDDAH916/2020 | EPI_ISL_513047 |
| 917 | SaudiArabia/KAUST-JEDDAH917/2020 | EPI_ISL_513048 |
| 918 | SaudiArabia/KAUST-JEDDAH918/2020 | EPI_ISL_513049 |
| 919 | SaudiArabia/KAUST-JEDDAH919/2020 | EPI_ISL_513050 |
| 920 | SaudiArabia/KAUST-JEDDAH920/2020 | EPI_ISL_513051 |
| 921 | SaudiArabia/KAUST-JEDDAH921/2020 | EPI_ISL_513052 |
| 922 | SaudiArabia/KAUST-JEDDAH922/2020 | EPI_ISL_513053 |
| 923 | SaudiArabia/KAUST-JEDDAH923/2020 | EPI_ISL_513054 |
| 925 | SaudiArabia/KAUST-JEDDAH925/2020 | EPI_ISL_678236 |
| 926 | SaudiArabia/KAUST-JEDDAH926/2020 | EPI_ISL_513055 |
| 927 | SaudiArabia/KAUST-JEDDAH927/2020 | EPI_ISL_513056 |

| | | |
|---|---|---|
| 929 | SaudiArabia/KAUST-JEDDAH929/2020 | EPI_ISL_513057 |
| 930 | SaudiArabia/KAUST-JEDDAH930/2020 | EPI_ISL_513058 |
| 931 | SaudiArabia/KAUST-JEDDAH931/2020 | EPI_ISL_513059 |
| 932 | SaudiArabia/KAUST-JEDDAH932/2020 | EPI_ISL_513060 |
| 933 | SaudiArabia/KAUST-JEDDAH933/2020 | EPI_ISL_513061 |
| 935 | SaudiArabia/KAUST-JEDDAH935/2020 | EPI_ISL_513062 |
| 936 | SaudiArabia/KAUST-JEDDAH936/2020 | EPI_ISL_513063 |
| 937 | SaudiArabia/KAUST-MADINAH937/2020 | EPI_ISL_513201 |
| 938 | SaudiArabia/KAUST-MADINAH938/2020 | EPI_ISL_513202 |
| 941 | SaudiArabia/KAUST-MADINAH941/2020 | EPI_ISL_513203 |
| 947 | SaudiArabia/KAUST-MADINAH947/2020 | EPI_ISL_513204 |
| 955 | SaudiArabia/KAUST-MADINAH955/2020 | EPI_ISL_513205 |
| 956 | SaudiArabia/KAUST-MADINAH956/2020 | EPI_ISL_513206 |
| 957 | SaudiArabia/KAUST-MADINAH957/2020 | EPI_ISL_513207 |
| 958 | SaudiArabia/KAUST-MADINAH958/2020 | EPI_ISL_513208 |
| 959 | SaudiArabia/KAUST-MADINAH959/2020 | EPI_ISL_513209 |
| 960 | SaudiArabia/KAUST-MADINAH960/2020 | EPI_ISL_513210 |
| 961 | SaudiArabia/KAUST-MADINAH961/2020 | EPI_ISL_513211 |
| 962 | SaudiArabia/KAUST-MADINAH962/2020 | EPI_ISL_751203 |
| 967 | SaudiArabia/KAUST-MADINAH967/2020 | EPI_ISL_513212 |
| 975 | SaudiArabia/KAUST-MADINAH975/2020 | EPI_ISL_513213 |
| 990 | SaudiArabia/KAUST-JEDDAH990/2020 | EPI_ISL_513064 |
| 991 | SaudiArabia/KAUST-JEDDAH991/2020 | EPI_ISL_513065 |
| 992 | SaudiArabia/KAUST-JEDDAH992/2020 | EPI_ISL_513066 |
| 993 | SaudiArabia/KAUST-JEDDAH993/2020 | EPI_ISL_513067 |
| 994 | SaudiArabia/KAUST-JEDDAH994/2020 | EPI_ISL_636965 |
| 995 | SaudiArabia/KAUST-JEDDAH995/2020 | EPI_ISL_513068 |
| 996 | SaudiArabia/KAUST-JEDDAH996/2020 | EPI_ISL_513069 |
| 997 | SaudiArabia/KAUST-JEDDAH997/2020 | EPI_ISL_513070 |
| 998 | SaudiArabia/KAUST-JEDDAH998/2020 | EPI_ISL_513071 |
| 999 | SaudiArabia/KAUST-JEDDAH999/2020 | EPI_ISL_513072 |
| 1000 | SaudiArabia/KAUST-JEDDAH1000/2020 | EPI_ISL_512874 |
| 1001 | SaudiArabia/KAUST-JEDDAH1001/2020 | EPI_ISL_512875 |
| 1002 | SaudiArabia/KAUST-JEDDAH1002/2020 | EPI_ISL_512876 |
| 1003 | SaudiArabia/KAUST-JEDDAH1003/2020 | EPI_ISL_512877 |
| 1007 | SaudiArabia/KAUST-JEDDAH1007/2020 | EPI_ISL_512878 |
| 1008 | SaudiArabia/KAUST-JEDDAH1008/2020 | EPI_ISL_512879 |
| 1009 | SaudiArabia/KAUST-JEDDAH1009/2020 | EPI_ISL_512880 |
| 1010 | SaudiArabia/KAUST-JEDDAH1010/2020 | EPI_ISL_512881 |
| 1011 | SaudiArabia/KAUST-JEDDAH1011/2020 | EPI_ISL_512882 |
| 1012 | SaudiArabia/KAUST-JEDDAH1012/2020 | EPI_ISL_512883 |
| 1013 | SaudiArabia/KAUST-JEDDAH1013/2020 | EPI_ISL_512884 |
| 1014 | SaudiArabia/KAUST-JEDDAH1014/2020 | EPI_ISL_512885 |
| 1015 | SaudiArabia/KAUST-JEDDAH1015/2020 | EPI_ISL_512886 |
| 1016 | SaudiArabia/KAUST-JEDDAH1016/2020 | EPI_ISL_512887 |
| 1017 | SaudiArabia/KAUST-JEDDAH1017/2020 | EPI_ISL_512888 |

| | | |
|---|---|---|
| 1018 | SaudiArabia/KAUST-JEDDAH1018/2020 | EPI_ISL_512889 |
| 1020 | SaudiArabia/KAUST-JEDDAH1020/2020 | EPI_ISL_512890 |
| 1021 | SaudiArabia/KAUST-JEDDAH1021/2020 | EPI_ISL_512891 |
| 1022 | SaudiArabia/KAUST-JEDDAH1022/2020 | EPI_ISL_512892 |
| 1023 | SaudiArabia/KAUST-JEDDAH1023/2020 | EPI_ISL_512893 |
| 1024 | SaudiArabia/KAUST-JEDDAH1024/2020 | EPI_ISL_512894 |
| 1025 | SaudiArabia/KAUST-JEDDAH1025/2020 | EPI_ISL_512895 |
| 1026 | SaudiArabia/KAUST-JEDDAH1026/2020 | EPI_ISL_512896 |
| 1027 | SaudiArabia/KAUST-JEDDAH1027/2020 | EPI_ISL_512897 |
| 1028 | SaudiArabia/KAUST-JEDDAH1028/2020 | EPI_ISL_512898 |
| 1030 | SaudiArabia/KAUST-JEDDAH1030/2020 | EPI_ISL_512899 |
| 1031 | SaudiArabia/KAUST-JEDDAH1031/2020 | EPI_ISL_512900 |
| 1032 | SaudiArabia/KAUST-JEDDAH1032/2020 | EPI_ISL_512901 |
| 1033 | SaudiArabia/KAUST-JEDDAH1033/2020 | EPI_ISL_512902 |
| 1034 | SaudiArabia/KAUST-JEDDAH1034/2020 | EPI_ISL_512903 |
| 1036 | SaudiArabia/KAUST-MADINAH1036/2020 | EPI_ISL_513073 |
| 1038 | SaudiArabia/KAUST-MADINAH1038/2020 | EPI_ISL_513074 |
| 1043 | SaudiArabia/KAUST-MADINAH1043/2020 | EPI_ISL_513075 |
| 1044 | SaudiArabia/KAUST-MADINAH1044/2020 | EPI_ISL_513076 |
| 1046 | SaudiArabia/KAUST-MADINAH1046/2020 | EPI_ISL_513077 |
| 1047 | SaudiArabia/KAUST-MADINAH1047/2020 | EPI_ISL_513078 |
| 1048 | SaudiArabia/KAUST-MADINAH1048/2020 | EPI_ISL_513079 |
| 1049 | SaudiArabia/KAUST-MADINAH1049/2020 | EPI_ISL_513080 |
| 1051 | SaudiArabia/KAUST-MADINAH1051/2020 | EPI_ISL_513081 |
| 1052 | SaudiArabia/KAUST-MADINAH1052/2020 | EPI_ISL_513082 |
| 1053 | SaudiArabia/KAUST-MADINAH1053/2020 | EPI_ISL_513083 |
| 1054 | SaudiArabia/KAUST-JEDDAH1054/2020 | EPI_ISL_512904 |
| 1055 | SaudiArabia/KAUST-JEDDAH1055/2020 | EPI_ISL_512905 |
| 1056 | SaudiArabia/KAUST-JEDDAH1056/2020 | EPI_ISL_636966 |
| 1057 | SaudiArabia/KAUST-JEDDAH1057/2020 | EPI_ISL_512906 |
| 1058 | SaudiArabia/KAUST-JEDDAH1058/2020 | EPI_ISL_512907 |
| 1059 | SaudiArabia/KAUST-MADINAH1059/2020 | EPI_ISL_513084 |
| 1060 | SaudiArabia/KAUST-MADINAH1060/2020 | EPI_ISL_513085 |
| 1061 | SaudiArabia/KAUST-MADINAH1061/2020 | EPI_ISL_513086 |
| 1063 | SaudiArabia/KAUST-MADINAH1063/2020 | EPI_ISL_677951 |
| 1066 | SaudiArabia/KAUST-MADINAH1066/2020 | EPI_ISL_513087 |
| 1070 | SaudiArabia/KAUST-MADINAH1070/2020 | EPI_ISL_513088 |
| 1072 | SaudiArabia/KAUST-MADINAH1072/2020 | EPI_ISL_513089 |
| 1073 | SaudiArabia/KAUST-MADINAH1073/2020 | EPI_ISL_513090 |
| 1074 | SaudiArabia/KAUST-MADINAH1074/2020 | EPI_ISL_513091 |
| 1075 | SaudiArabia/KAUST-MADINAH1075/2020 | EPI_ISL_513092 |
| 1076 | SaudiArabia/KAUST-MADINAH1076/2020 | EPI_ISL_513093 |
| 1077 | SaudiArabia/KAUST-MADINAH1077/2020 | EPI_ISL_513094 |
| 1078 | SaudiArabia/KAUST-MADINAH1078/2020 | EPI_ISL_513095 |
| 1079 | SaudiArabia/KAUST-MADINAH1079/2020 | EPI_ISL_513096 |
| 1080 | SaudiArabia/KAUST-MADINAH1080/2020 | EPI_ISL_636967 |

1081 SaudiArabia/KAUST-MADINAH1081/2020 EPI_ISL_513097
1082 SaudiArabia/KAUST-MADINAH1082/2020 EPI_ISL_636968
1085 SaudiArabia/KAUST-MADINAH1085/2020 EPI_ISL_513098
1086 SaudiArabia/KAUST-MADINAH1086/2020 EPI_ISL_513099
1087 SaudiArabia/KAUST-MADINAH1087/2020 EPI_ISL_513100
1090 SaudiArabia/KAUST-MADINAH1090/2020 EPI_ISL_513101
1093 SaudiArabia/KAUST-MADINAH1093/2020 EPI_ISL_513102
1094 SaudiArabia/KAUST-MADINAH1094/2020 EPI_ISL_513103
1095 SaudiArabia/KAUST-MADINAH1095/2020 EPI_ISL_513104
1096 SaudiArabia/KAUST-MADINAH1096/2020 EPI_ISL_513105
1099 SaudiArabia/KAUST-MADINAH1099/2020 EPI_ISL_513106
1100 SaudiArabia/KAUST-MADINAH1100/2020 EPI_ISL_513107
1101 SaudiArabia/KAUST-MADINAH1101/2020 EPI_ISL_677972
1102 SaudiArabia/KAUST-MADINAH1102/2020 EPI_ISL_513108
1103 SaudiArabia/KAUST-MADINAH1103/2020 EPI_ISL_513109
1106 SaudiArabia/KAUST-MADINAH1106/2020 EPI_ISL_513110
1107 SaudiArabia/KAUST-MADINAH1107/2020 EPI_ISL_513111
1108 SaudiArabia/KAUST-MADINAH1108/2020 EPI_ISL_513112
1109 SaudiArabia/KAUST-MADINAH1109/2020 EPI_ISL_513113
1110 SaudiArabia/KAUST-MADINAH1110/2020 EPI_ISL_513114
1111 SaudiArabia/KAUST-MADINAH1111/2020 EPI_ISL_513115
1112 SaudiArabia/KAUST-MADINAH1112/2020 EPI_ISL_513116
1113 SaudiArabia/KAUST-MADINAH1113/2020 EPI_ISL_513117
1114 SaudiArabia/KAUST-MADINAH1114/2020 EPI_ISL_513118
1115 SaudiArabia/KAUST-MADINAH1115/2020 EPI_ISL_513119
1116 SaudiArabia/KAUST-MADINAH1116/2020 EPI_ISL_513120
1117 SaudiArabia/KAUST-MADINAH1117/2020 EPI_ISL_677977
1118 SaudiArabia/KAUST-MADINAH1118/2020 EPI_ISL_677978
1119 SaudiArabia/KAUST-MADINAH1119/2020 EPI_ISL_677979
1120 SaudiArabia/KAUST-MADINAH1120/2020 EPI_ISL_677980
1121 SaudiArabia/KAUST-MADINAH1121/2020 EPI_ISL_677981
1122 SaudiArabia/KAUST-MADINAH1122/2020 EPI_ISL_677982
1123 SaudiArabia/KAUST-MADINAH1123/2020 EPI_ISL_677983
1124 SaudiArabia/KAUST-MADINAH1124/2020 EPI_ISL_677984
1126 SaudiArabia/KAUST-MADINAH1126/2020 EPI_ISL_677985
1127 SaudiArabia/KAUST-MADINAH1127/2020 EPI_ISL_677986
1128 SaudiArabia/KAUST-MADINAH1128/2020 EPI_ISL_677975
1129 SaudiArabia/KAUST-MADINAH1129/2020 EPI_ISL_677987
1130 SaudiArabia/KAUST-MADINAH1130/2020 EPI_ISL_677988
1131 SaudiArabia/KAUST-MADINAH1131/2020 EPI_ISL_677989
1132 SaudiArabia/KAUST-MADINAH1132/2020 EPI_ISL_678237
1135 SaudiArabia/KAUST-MADINAH1135/2020 EPI_ISL_677967
1136 SaudiArabia/KAUST-MADINAH1136/2020 EPI_ISL_677990
1139 SaudiArabia/KAUST-MADINAH1139/2020 EPI_ISL_677991
1140 SaudiArabia/KAUST-MADINAH1140/2020 EPI_ISL_677992
1141 SaudiArabia/KAUST-MADINAH1141/2020 EPI_ISL_677993

| | | |
|---|---|---|
| 1142 | SaudiArabia/KAUST-MADINAH1142/2020 | EPI_ISL_677994 |
| 1143 | SaudiArabia/KAUST-MADINAH1143/2020 | EPI_ISL_677995 |
| 1144 | SaudiArabia/KAUST-MADINAH1144/2020 | EPI_ISL_677996 |
| 1145 | SaudiArabia/KAUST-MADINAH1145/2020 | EPI_ISL_677997 |
| 1146 | SaudiArabia/KAUST-MADINAH1146/2020 | EPI_ISL_677998 |
| 1147 | SaudiArabia/KAUST-MADINAH1147/2020 | EPI_ISL_677999 |
| 1148 | SaudiArabia/KAUST-MADINAH1148/2020 | EPI_ISL_678000 |
| 1149 | SaudiArabia/KAUST-MADINAH1149/2020 | EPI_ISL_678001 |
| 1150 | SaudiArabia/KAUST-MADINAH1150/2020 | EPI_ISL_678002 |
| 1151 | SaudiArabia/KAUST-MADINAH1151/2020 | EPI_ISL_678003 |
| 1155 | SaudiArabia/KAUST-RIYADH1155/2020 | EPI_ISL_678167 |
| 1157 | SaudiArabia/KAUST-RIYADH1157/2020 | EPI_ISL_678190 |
| 1158 | SaudiArabia/KAUST-RIYADH1158/2020 | EPI_ISL_678187 |
| 1162 | SaudiArabia/KAUST-RIYADH1162/2020 | EPI_ISL_678188 |
| 1163 | SaudiArabia/KAUST-RIYADH1163/2020 | EPI_ISL_678191 |
| 1164 | SaudiArabia/KAUST-RIYADH1164/2020 | EPI_ISL_678181 |
| 1166 | SaudiArabia/KAUST-RIYADH1166/2020 | EPI_ISL_678175 |
| 1167 | SaudiArabia/KAUST-RIYADH1167/2020 | EPI_ISL_678160 |
| 1168 | SaudiArabia/KAUST-RIYADH1168/2020 | EPI_ISL_678169 |
| 1171 | SaudiArabia/KAUST-RIYADH1171/2020 | EPI_ISL_678192 |
| 1172 | SaudiArabia/KAUST-RIYADH1172/2020 | EPI_ISL_678193 |
| 1173 | SaudiArabia/KAUST-RIYADH1173/2020 | EPI_ISL_751222 |
| 1174 | SaudiArabia/KAUST-RIYADH1174/2020 | EPI_ISL_678194 |
| 1175 | SaudiArabia/KAUST-RIYADH1175/2020 | EPI_ISL_678173 |
| 1176 | SaudiArabia/KAUST-RIYADH1176/2020 | EPI_ISL_678195 |
| 1178 | SaudiArabia/KAUST-RIYADH1178/2020 | EPI_ISL_678196 |
| 1179 | SaudiArabia/KAUST-RIYADH1179/2020 | EPI_ISL_678161 |
| 1180 | SaudiArabia/KAUST-RIYADH1180/2020 | EPI_ISL_678197 |
| 1181 | SaudiArabia/KAUST-RIYADH1181/2020 | EPI_ISL_678189 |
| 1187 | SaudiArabia/KAUST-RIYADH1187/2020 | EPI_ISL_678166 |
| 1191 | SaudiArabia/KAUST-RIYADH1191/2020 | EPI_ISL_678198 |
| 1192 | SaudiArabia/KAUST-RIYADH1192/2020 | EPI_ISL_678199 |
| 1193 | SaudiArabia/KAUST-RIYADH1193/2020 | EPI_ISL_678183 |
| 1194 | SaudiArabia/KAUST-RIYADH1194/2020 | EPI_ISL_678200 |
| 1195 | SaudiArabia/KAUST-RIYADH1195/2020 | EPI_ISL_678201 |
| 1196 | SaudiArabia/KAUST-RIYADH1196/2020 | EPI_ISL_678202 |
| 1198 | SaudiArabia/KAUST-RIYADH1198/2020 | EPI_ISL_678163 |
| 1200 | SaudiArabia/KAUST-RIYADH1200/2020 | EPI_ISL_678186 |
| 1205 | SaudiArabia/KAUST-RIYADH1205/2020 | EPI_ISL_678203 |
| 1206 | SaudiArabia/KAUST-RIYADH1206/2020 | EPI_ISL_678176 |
| 1207 | SaudiArabia/KAUST-RIYADH1207/2020 | EPI_ISL_678204 |
| 1208 | SaudiArabia/KAUST-RIYADH1208/2020 | EPI_ISL_678205 |
| 1211 | SaudiArabia/KAUST-RIYADH1211/2020 | EPI_ISL_678168 |
| 1214 | SaudiArabia/KAUST-RIYADH1214/2020 | EPI_ISL_678206 |
| 1215 | SaudiArabia/KAUST-RIYADH1215/2020 | EPI_ISL_678207 |
| 1216 | SaudiArabia/KAUST-RIYADH1216/2020 | EPI_ISL_678208 |

| | | |
|---|---|---|
| 1219 | SaudiArabia/KAUST-RIYADH1219/2020 | EPI_ISL_678209 |
| 1220 | SaudiArabia/KAUST-RIYADH1220/2020 | EPI_ISL_678210 |
| 1222 | SaudiArabia/KAUST-RIYADH1222/2020 | EPI_ISL_678211 |
| 1229 | SaudiArabia/KAUST-RIYADH1229/2020 | EPI_ISL_678238 |
| 1231 | SaudiArabia/KAUST-RIYADH1231/2020 | EPI_ISL_678170 |
| 1233 | SaudiArabia/KAUST-RIYADH1233/2020 | EPI_ISL_678247 |
| 1234 | SaudiArabia/KAUST-RIYADH1234/2020 | EPI_ISL_678174 |
| 1235 | SaudiArabia/KAUST-RIYADH1235/2020 | EPI_ISL_678212 |
| 1239 | SaudiArabia/KAUST-RIYADH1239/2020 | EPI_ISL_678184 |
| 1240 | SaudiArabia/KAUST-RIYADH1240/2020 | EPI_ISL_678239 |
| 1241 | SaudiArabia/KAUST-RIYADH1241/2020 | EPI_ISL_678213 |
| 1243 | SaudiArabia/KAUST-RIYADH1243/2020 | EPI_ISL_678214 |
| 1245 | SaudiArabia/KAUST-RIYADH1245/2020 | EPI_ISL_678240 |
| 1246 | SaudiArabia/KAUST-RIYADH1246/2020 | EPI_ISL_678165 |
| 1247 | SaudiArabia/KAUST-RIYADH1247/2020 | EPI_ISL_678178 |
| 1248 | SaudiArabia/KAUST-RIYADH1248/2020 | EPI_ISL_678171 |
| 1249 | SaudiArabia/KAUST-RIYADH1249/2020 | EPI_ISL_678215 |
| 1250 | SaudiArabia/KAUST-RIYADH1250/2020 | EPI_ISL_678216 |
| 1251 | SaudiArabia/KAUST-RIYADH1251/2020 | EPI_ISL_678217 |
| 1254 | SaudiArabia/KAUST-RIYADH1254/2020 | EPI_ISL_678218 |
| 1255 | SaudiArabia/KAUST-RIYADH1255/2020 | EPI_ISL_678179 |
| 1256 | SaudiArabia/KAUST-RIYADH1256/2020 | EPI_ISL_678182 |
| 1258 | SaudiArabia/KAUST-RIYADH1258/2020 | EPI_ISL_678164 |
| 1259 | SaudiArabia/KAUST-RIYADH1259/2020 | EPI_ISL_678219 |
| 1261 | SaudiArabia/KAUST-RIYADH1261/2020 | EPI_ISL_678220 |
| 1262 | SaudiArabia/KAUST-RIYADH1262/2020 | EPI_ISL_678185 |
| 1263 | SaudiArabia/KAUST-RIYADH1263/2020 | EPI_ISL_678221 |
| 1264 | SaudiArabia/KAUST-RIYADH1264/2020 | EPI_ISL_678222 |
| 1265 | SaudiArabia/KAUST-RIYADH1265/2020 | EPI_ISL_678223 |
| 1267 | SaudiArabia/KAUST-RIYADH1267/2020 | EPI_ISL_678224 |
| 1268 | SaudiArabia/KAUST-RIYADH1268/2020 | EPI_ISL_678225 |
| 1269 | SaudiArabia/KAUST-MADINAH1269/2020 | EPI_ISL_678004 |
| 1271 | SaudiArabia/KAUST-MADINAH1271/2020 | EPI_ISL_678005 |
| 1274 | SaudiArabia/KAUST-MADINAH1274/2020 | EPI_ISL_678006 |
| 1277 | SaudiArabia/KAUST-MADINAH1277/2020 | EPI_ISL_678007 |
| 1279 | SaudiArabia/KAUST-MADINAH1279/2020 | EPI_ISL_677971 |
| 1281 | SaudiArabia/KAUST-MADINAH1281/2020 | EPI_ISL_677950 |
| 1282 | SaudiArabia/KAUST-MADINAH1282/2020 | EPI_ISL_677952 |
| 1283 | SaudiArabia/KAUST-MADINAH1283/2020 | EPI_ISL_678008 |
| 1284 | SaudiArabia/KAUST-MADINAH1284/2020 | EPI_ISL_678009 |
| 1285 | SaudiArabia/KAUST-RIYADH1285/2020 | EPI_ISL_678162 |
| 1286 | SaudiArabia/KAUST-RIYADH1286/2020 | EPI_ISL_678177 |
| 1291 | SaudiArabia/KAUST-JEDDAH1291/2020 | EPI_ISL_677922 |
| 1292 | SaudiArabia/KAUST-JEDDAH1292/2020 | EPI_ISL_677923 |
| 1293 | SaudiArabia/KAUST-JEDDAH1293/2020 | EPI_ISL_678241 |
| 1294 | SaudiArabia/KAUST-JEDDAH1294/2020 | EPI_ISL_677911 |

| | | |
|---|---|---|
| 1295 | SaudiArabia/KAUST-JEDDAH1295/2020 | EPI_ISL_677924 |
| 1296 | SaudiArabia/KAUST-JEDDAH1296/2020 | EPI_ISL_677925 |
| 1297 | SaudiArabia/KAUST-JEDDAH1297/2020 | EPI_ISL_677914 |
| 1299 | SaudiArabia/KAUST-JEDDAH1299/2020 | EPI_ISL_677926 |
| 1301 | SaudiArabia/KAUST-JEDDAH1301/2020 | EPI_ISL_677927 |
| 1312 | SaudiArabia/KAUST-JEDDAH1312/2020 | EPI_ISL_678242 |
| 1316 | SaudiArabia/KAUST-JEDDAH1316/2020 | EPI_ISL_677928 |
| 1317 | SaudiArabia/KAUST-JEDDAH1317/2020 | EPI_ISL_677929 |
| 1318 | SaudiArabia/KAUST-JEDDAH1318/2020 | EPI_ISL_677930 |
| 1319 | SaudiArabia/KAUST-JEDDAH1319/2020 | EPI_ISL_677931 |
| 1320 | SaudiArabia/KAUST-JEDDAH1320/2020 | EPI_ISL_677921 |
| 1321 | SaudiArabia/KAUST-JEDDAH1321/2020 | EPI_ISL_677917 |
| 1323 | SaudiArabia/KAUST-JEDDAH1323/2020 | EPI_ISL_678243 |
| 1324 | SaudiArabia/KAUST-JEDDAH1324/2020 | EPI_ISL_677910 |
| 1325 | SaudiArabia/KAUST-JEDDAH1325/2020 | EPI_ISL_677932 |
| 1326 | SaudiArabia/KAUST-JEDDAH1326/2020 | EPI_ISL_677913 |
| 1329 | SaudiArabia/KAUST-JEDDAH1329/2020 | EPI_ISL_677912 |
| 1330 | SaudiArabia/KAUST-JEDDAH1330/2020 | EPI_ISL_677908 |
| 1331 | SaudiArabia/KAUST-JEDDAH1331/2020 | EPI_ISL_677918 |
| 1332 | SaudiArabia/KAUST-JEDDAH1332/2020 | EPI_ISL_677909 |
| 1333 | SaudiArabia/KAUST-JEDDAH1333/2020 | EPI_ISL_677933 |
| 1334 | SaudiArabia/KAUST-JEDDAH1334/2020 | EPI_ISL_677919 |
| 1335 | SaudiArabia/KAUST-JEDDAH1335/2020 | EPI_ISL_677920 |
| 1337 | SaudiArabia/KAUST-JEDDAH1337/2020 | EPI_ISL_677934 |
| 1338 | SaudiArabia/KAUST-JEDDAH1338/2020 | EPI_ISL_677935 |
| 1339 | SaudiArabia/KAUST-JEDDAH1339/2020 | EPI_ISL_677936 |
| 1340 | SaudiArabia/KAUST-JEDDAH1340/2020 | EPI_ISL_677916 |
| 1341 | SaudiArabia/KAUST-JEDDAH1341/2020 | EPI_ISL_677915 |
| 1344 | SaudiArabia/KAUST-QATIF1344/2020 | EPI_ISL_678152 |
| 1351 | SaudiArabia/KAUST-QATIF1351/2020 | EPI_ISL_678147 |
| 1352 | SaudiArabia/KAUST-QATIF1352/2020 | EPI_ISL_678153 |
| 1353 | SaudiArabia/KAUST-QATIF1353/2020 | EPI_ISL_678154 |
| 1354 | SaudiArabia/KAUST-QATIF1354/2020 | EPI_ISL_678244 |
| 1355 | SaudiArabia/KAUST-QATIF1355/2020 | EPI_ISL_678148 |
| 1362 | SaudiArabia/KAUST-MADINAH1362/2020 | EPI_ISL_678010 |
| 1363 | SaudiArabia/KAUST-MADINAH1363/2020 | EPI_ISL_678011 |
| 1364 | SaudiArabia/KAUST-MADINAH1364/2020 | EPI_ISL_677949 |
| 1366 | SaudiArabia/KAUST-MADINAH1366/2020 | EPI_ISL_678012 |
| 1367 | SaudiArabia/KAUST-MADINAH1367/2020 | EPI_ISL_678013 |
| 1370 | SaudiArabia/KAUST-MADINAH1370/2020 | EPI_ISL_677954 |
| 1371 | SaudiArabia/KAUST-MADINAH1371/2020 | EPI_ISL_677959 |
| 1372 | SaudiArabia/KAUST-MADINAH1372/2020 | EPI_ISL_678014 |
| 1373 | SaudiArabia/KAUST-MADINAH1373/2020 | EPI_ISL_678015 |
| 1374 | SaudiArabia/KAUST-MADINAH1374/2020 | EPI_ISL_678016 |
| 1375 | SaudiArabia/KAUST-MADINAH1375/2020 | EPI_ISL_678017 |
| 1378 | SaudiArabia/KAUST-MADINAH1378/2020 | EPI_ISL_677948 |

| | | |
|---|---|---|
| 1379 | SaudiArabia/KAUST-MADINAH1379/2020 | EPI_ISL_678018 |
| 1380 | SaudiArabia/KAUST-MADINAH1380/2020 | EPI_ISL_677961 |
| 1381 | SaudiArabia/KAUST-MADINAH1381/2020 | EPI_ISL_678019 |
| 1386 | SaudiArabia/KAUST-MAKKAH1386/2020 | EPI_ISL_678064 |
| 1388 | SaudiArabia/KAUST-MAKKAH1388/2020 | EPI_ISL_678083 |
| 1392 | SaudiArabia/KAUST-MAKKAH1392/2020 | EPI_ISL_678084 |
| 1395 | SaudiArabia/KAUST-MAKKAH1395/2020 | EPI_ISL_678085 |
| 1399 | SaudiArabia/KAUST-MAKKAH1399/2020 | EPI_ISL_678086 |
| 1401 | SaudiArabia/KAUST-MAKKAH1401/2020 | EPI_ISL_678087 |
| 1404 | SaudiArabia/KAUST-MAKKAH1404/2020 | EPI_ISL_678088 |
| 1406 | SaudiArabia/KAUST-MAKKAH1406/2020 | EPI_ISL_678089 |
| 1408 | SaudiArabia/KAUST-MAKKAH1408/2020 | EPI_ISL_678090 |
| 1410 | SaudiArabia/KAUST-MAKKAH1410/2020 | EPI_ISL_678091 |
| 1413 | SaudiArabia/KAUST-MAKKAH1413/2020 | EPI_ISL_678070 |
| 1415 | SaudiArabia/KAUST-MAKKAH1415/2020 | EPI_ISL_678092 |
| 1416 | SaudiArabia/KAUST-MAKKAH1416/2020 | EPI_ISL_678049 |
| 1421 | SaudiArabia/KAUST-MAKKAH1421/2020 | EPI_ISL_678093 |
| 1426 | SaudiArabia/KAUST-MAKKAH1426/2020 | EPI_ISL_678094 |
| 1427 | SaudiArabia/KAUST-MAKKAH1427/2020 | EPI_ISL_678095 |
| 1441 | SaudiArabia/KAUST-QATIF1441/2020 | EPI_ISL_678151 |
| 1442 | SaudiArabia/KAUST-RIYADH1442/2020 | EPI_ISL_678226 |
| 1443 | SaudiArabia/KAUST-QATIF1443/2020 | EPI_ISL_678155 |
| 1446 | SaudiArabia/KAUST-RIYADH1446/2020 | EPI_ISL_678180 |
| 1447 | SaudiArabia/KAUST-QATIF1447/2020 | EPI_ISL_678150 |
| 1448 | SaudiArabia/KAUST-QATIF1448/2020 | EPI_ISL_678156 |
| 1451 | SaudiArabia/KAUST-QATIF1451/2020 | EPI_ISL_678149 |
| 1452 | SaudiArabia/KAUST-QATIF1452/2020 | EPI_ISL_678157 |
| 1454 | SaudiArabia/KAUST-RIYADH1454/2020 | EPI_ISL_678172 |
| 1457 | SaudiArabia/KAUST-QATIF1457/2020 | EPI_ISL_678158 |
| 1468 | SaudiArabia/KAUST-QATIF1468/2020 | EPI_ISL_678159 |
| 1472 | SaudiArabia/KAUST-QATIF1472/2020 | EPI_ISL_678245 |
| 1478 | SaudiArabia/KAUST-MADINAH1478/2020 | EPI_ISL_678020 |
| 1479 | SaudiArabia/KAUST-MADINAH1479/2020 | EPI_ISL_677953 |
| 1480 | SaudiArabia/KAUST-MADINAH1480/2020 | EPI_ISL_678021 |
| 1481 | SaudiArabia/KAUST-MADINAH1481/2020 | EPI_ISL_677962 |
| 1482 | SaudiArabia/KAUST-MADINAH1482/2020 | EPI_ISL_677969 |
| 1483 | SaudiArabia/KAUST-MADINAH1483/2020 | EPI_ISL_678022 |
| 1484 | SaudiArabia/KAUST-MADINAH1484/2020 | EPI_ISL_678023 |
| 1487 | SaudiArabia/KAUST-MADINAH1487/2020 | EPI_ISL_678024 |
| 1488 | SaudiArabia/KAUST-MADINAH1488/2020 | EPI_ISL_678025 |
| 1489 | SaudiArabia/KAUST-MADINAH1489/2020 | EPI_ISL_677974 |
| 1490 | SaudiArabia/KAUST-MADINAH1490/2020 | EPI_ISL_677956 |
| 1491 | SaudiArabia/KAUST-MADINAH1491/2020 | EPI_ISL_677957 |
| 1496 | SaudiArabia/KAUST-MADINAH1496/2020 | EPI_ISL_751500 |
| 1497 | SaudiArabia/KAUST-MADINAH1497/2020 | EPI_ISL_751228 |
| 1498 | SaudiArabia/KAUST-MADINAH1498/2020 | EPI_ISL_751214 |

| | | |
|---|---|---|
| 1501 | SaudiArabia/KAUST-MAKKAH1501/2020 | EPI_ISL_751213 |
| 1503 | SaudiArabia/KAUST-MAKKAH1503/2020 | EPI_ISL_678097 |
| 1504 | SaudiArabia/KAUST-MAKKAH1504/2020 | EPI_ISL_678098 |
| 1505 | SaudiArabia/KAUST-MAKKAH1505/2020 | EPI_ISL_751238 |
| 1513 | SaudiArabia/KAUST-MAKKAH1513/2020 | EPI_ISL_678045 |
| 1515 | SaudiArabia/KAUST-MAKKAH1515/2020 | EPI_ISL_678099 |
| 1516 | SaudiArabia/KAUST-MAKKAH1516/2020 | EPI_ISL_678046 |
| 1517 | SaudiArabia/KAUST-MAKKAH1517/2020 | EPI_ISL_678100 |
| 1520 | SaudiArabia/KAUST-MAKKAH1520/2020 | EPI_ISL_678101 |
| 1525 | SaudiArabia/KAUST-MAKKAH1525/2020 | EPI_ISL_678102 |
| 1527 | SaudiArabia/KAUST-MAKKAH1527/2020 | EPI_ISL_678103 |
| 1534 | SaudiArabia/KAUST-MAKKAH1534/2020 | EPI_ISL_751223 |
| 1536 | SaudiArabia/KAUST-MAKKAH1536/2020 | EPI_ISL_678104 |
| 1537 | SaudiArabia/KAUST-MAKKAH1537/2020 | EPI_ISL_678105 |
| 1539 | SaudiArabia/KAUST-MAKKAH1539/2020 | EPI_ISL_678106 |
| 1541 | SaudiArabia/KAUST-MAKKAH1541/2020 | EPI_ISL_678053 |
| 1542 | SaudiArabia/KAUST-MAKKAH1542/2020 | EPI_ISL_678107 |
| 1547 | SaudiArabia/KAUST-MAKKAH1547/2020 | EPI_ISL_678051 |
| 1549 | SaudiArabia/KAUST-MAKKAH1549/2020 | EPI_ISL_678108 |
| 1554 | SaudiArabia/KAUST-MAKKAH1554/2020 | EPI_ISL_678109 |
| 1557 | SaudiArabia/KAUST-MAKKAH1557/2020 | EPI_ISL_678110 |
| 1558 | SaudiArabia/KAUST-MAKKAH1558/2020 | EPI_ISL_678111 |
| 1559 | SaudiArabia/KAUST-MAKKAH1559/2020 | EPI_ISL_678112 |
| 1560 | SaudiArabia/KAUST-MAKKAH1560/2020 | EPI_ISL_678113 |
| 1562 | SaudiArabia/KAUST-MAKKAH1562/2020 | EPI_ISL_678114 |
| 1566 | SaudiArabia/KAUST-MAKKAH1566/2020 | EPI_ISL_678115 |
| 1571 | SaudiArabia/KAUST-MAKKAH1571/2020 | EPI_ISL_751211 |
| 1582 | SaudiArabia/KAUST-MAKKAH1582/2020 | EPI_ISL_751206 |
| 1588 | SaudiArabia/KAUST-MAKKAH1588/2020 | EPI_ISL_678118 |
| 1589 | SaudiArabia/KAUST-MAKKAH1589/2020 | EPI_ISL_751503 |
| 1592 | SaudiArabia/KAUST-MAKKAH1592/2020 | EPI_ISL_678119 |
| 1595 | SaudiArabia/KAUST-MAKKAH1595/2020 | EPI_ISL_678120 |
| 1596 | SaudiArabia/KAUST-MAKKAH1596/2020 | EPI_ISL_678121 |
| 1602 | SaudiArabia/KAUST-MAKKAH1602/2020 | EPI_ISL_678122 |
| 1604 | SaudiArabia/KAUST-MAKKAH1604/2020 | EPI_ISL_678123 |
| 1605 | SaudiArabia/KAUST-MAKKAH1605/2020 | EPI_ISL_678124 |
| 1607 | SaudiArabia/KAUST-MAKKAH1607/2020 | EPI_ISL_678050 |
| 1610 | SaudiArabia/KAUST-MAKKAH1610/2020 | EPI_ISL_678246 |
| 1611 | SaudiArabia/KAUST-MAKKAH1611/2020 | EPI_ISL_678125 |
| 1612 | SaudiArabia/KAUST-MAKKAH1612/2020 | EPI_ISL_678067 |
| 1614 | SaudiArabia/KAUST-MAKKAH1614/2020 | EPI_ISL_751202 |
| 1617 | SaudiArabia/KAUST-MAKKAH1617/2020 | EPI_ISL_678126 |
| 1621 | SaudiArabia/KAUST-MAKKAH1621/2020 | EPI_ISL_678128 |
| 1623 | SaudiArabia/KAUST-MAKKAH1623/2020 | EPI_ISL_678129 |
| 1626 | SaudiArabia/KAUST-MAKKAH1626/2020 | EPI_ISL_678130 |
| 1630 | SaudiArabia/KAUST-MAKKAH1630/2020 | EPI_ISL_678131 |

| | | |
|---|---|---|
| 1631 | SaudiArabia/KAUST-MAKKAH1631/2020 | EPI_ISL_678132 |
| 1632 | SaudiArabia/KAUST-MAKKAH1632/2020 | EPI_ISL_678133 |
| 1635 | SaudiArabia/KAUST-MAKKAH1635/2020 | EPI_ISL_678134 |
| 1638 | SaudiArabia/KAUST-MADINAH1638/2020 | EPI_ISL_751210 |
| 1648 | SaudiArabia/KAUST-MADINAH1648/2020 | EPI_ISL_751226 |
| 1651 | SaudiArabia/KAUST-MADINAH1651/2020 | EPI_ISL_751234 |
| 1653 | SaudiArabia/KAUST-MADINAH1653/2020 | EPI_ISL_751236 |
| 1655 | SaudiArabia/KAUST-MADINAH1655/2020 | EPI_ISL_751235 |
| 1657 | SaudiArabia/KAUST-MADINAH1657/2020 | EPI_ISL_751209 |
| 1659 | SaudiArabia/KAUST-MADINAH1659/2020 | EPI_ISL_751216 |
| 1661 | SaudiArabia/KAUST-MADINAH1661/2020 | EPI_ISL_751230 |
| 1667 | SaudiArabia/KAUST-MADINAH1667/2020 | EPI_ISL_751233 |
| 1669 | SaudiArabia/KAUST-MADINAH1669/2020 | EPI_ISL_751225 |
| 1670 | SaudiArabia/KAUST-MADINAH1670/2020 | EPI_ISL_751229 |
| 1671 | SaudiArabia/KAUST-MADINAH1671/2020 | EPI_ISL_751217 |
| 1674 | SaudiArabia/KAUST-MADINAH1674/2020 | EPI_ISL_751232 |
| 1677 | SaudiArabia/KAUST-MADINAH1677/2020 | EPI_ISL_751227 |
| 1688 | SaudiArabia/KAUST-MADINAH1688/2020 | EPI_ISL_751501 |
| 1689 | SaudiArabia/KAUST-MADINAH1689/2020 | EPI_ISL_751231 |
| 1690 | SaudiArabia/KAUST-MADINAH1690/2020 | EPI_ISL_751215 |
